# Use of Point-of-care Haemoglobin Tests to Diagnose Childhood Anaemia in Low-and Middle-Income Countries: A Systematic Review

**DOI:** 10.1101/2023.06.01.23290834

**Authors:** Rebecca Brehm, Annabelle South, Elizabeth C George

## Abstract

**Objectives:** Anaemia is a major cause of mortality and transfusion in children in Low- and Middle-Income Countries (LMICs), however current diagnostics are slow, costly, and frequently unavailable. Point-of-care haemoglobin tests (POC(Hb)Ts) could improve patient outcomes and use of resources by providing rapid and affordable results. We systematically reviewed the literature to investigate what, where and how POC(Hb)Ts are being used by health facilities in LMICs to diagnose childhood anaemia, and to explore challenges to their use.

**Methods:** We searched a total of nine databases and trial registries up to 10^th^ June 2022 using the concepts: anaemia, POC(Hb)T, LMIC and clinical setting. Adults ≥21 years and literature published >15 years ago were excluded. A single reviewer conducted screening, data extraction and quality assessment (of diagnostic studies) using QUADAS-2. Outcomes including POC(Hb)T used, location, setting, challenges and diagnostic accuracy were synthesised.

**Results:** Of 626 records screened, 41 studies were included. Evidence is available on the use of 15 POC(Hb)Ts in hospitals (n=28, 68%), health centres (n=9, 22%) and clinics/units (n=10, 24%) across 16 LMICs. HemoCue (HemoCue AB, Ängelholm, Sweden) was the most used test (n=31, 76%). Key challenges reported were overestimation of haemoglobin concentration, clinically unacceptable limits of agreement, errors/difficulty in sampling, environmental factors, cost, inter-observer variability, and supply of consumables. Five POC(Hb)Ts (33%) could not detect haemoglobin levels below 4g/dl. Diagnostic accuracy varied, with sensitivity and specificity to detect anaemia ranging from 24.2-92.2% and 70-96.7%, respectively.

**Conclusions:** POC(Hb)Ts have been successfully utilised in health facilities in LMICs to diagnose childhood anaemia. However, limited evidence is available, and challenges exist that must be addressed before wider implementation. Further research is required to confirm accuracy, clinical benefits, and cost-effectiveness.

## Introduction

Anaemia is a major global health problem, affecting over 1.8 billion people worldwide.^1, 2^ The condition is characterised by reduced blood haemoglobin (Hb), resulting in increased morbidity and mortality. Definitions vary by age, with anaemia and severe anaemia classified as Hb<11g/dl and Hb<7g/dl in children aged 6-59 months.^2^ Prevalence and years lived with disability are highest in Low-and Middle-Income Countries (LMICs), particularly Sub-Saharan Africa and South Asia where nutrient deficiencies, infectious diseases and haemoglobinopathies are common.^1, 3^ Children under five years of age are most vulnerable, with an estimated prevalence of 56.5% in LMICs.^4^

Severe anaemia is life-threatening and accounts for many hospital admissions in Sub-Saharan Africa. A large randomised controlled trial (RCT) investigating fluid bolus on mortality in hospitalised African children with severe infection, found 33% of presented children had Hb level <5g/dl and this resulted in increased mortality (FEAST).^5, 6^ Severe anaemia often requires emergency blood transfusion to restore Hb levels. However, this requires efficient diagnosis and availability of donated blood for effective treatment. This poses significant challenges in LMICs, where laboratory analysis is often lengthy, and stock-outs of blood are frequent.^7^ Delays in transfusions are common.^8^ Results from FEAST show that 52% of severely anaemic children died when not transfused within eight hours, with 90% of deaths occurring within 2.5 hours.^5, 6^ Therefore, prompt transfusion is critical to save lives.

Haematology analysers are the routine diagnostic method used to diagnose anaemia. However, equipment is expensive, requires electricity, trained personnel, and regular supply of reagents. This leads to them being often unavailable in LMICs, resulting in inaccurate diagnosis by clinical assessment and inappropriate use of transfusion.^9–11^ Point-of-care haemoglobin tests (POC(Hb)T) have been developed to help address these issues. These tests should be Affordable, Sensitive, Specific, User-friendly, Rapid and Robust, Equipment-free and Deliverable, according to World Health Organisation (WHO) ASSURED criteria.^12^ They are less invasive and provide immediate results (Hb level) at the site of care.^13^

POC(Hb)Ts have shown clinical benefit in African children with uncomplicated severe anaemia (TRACT) and valuable use in a range of settings.^14–22^ The TRACT trial found no significant difference in mortality between immediate and triggered transfusion (by new signs of severity or Hb<4g/dl) after 28 days (Hazard Ratio 0.54; 95% CI: 0.22-1.36; p=0.19).^22^ These results led to development of a treatment management algorithm to improve clinical practice.^23^ However, this approach requires regular use of POC(Hb)Ts every eight hours (first 24 hours), and at 48 hours. This trial provides strong evidence that if POC(Hb)Ts were adopted for routine use, transfusion requirements would decline by 60% and save valuable resources. Although, several POC(Hb)Ts have been developed and evaluated in recent years, few, if any, have been widely utilised by hospitals in LMICs for routine care. It is unclear to what extent POC(Hb)Ts have been employed to diagnose anaemia in underserved populations. Understanding where and what POC(Hb)Ts are currently used by health facilities and the barriers to their use, will help guide work to improve their availability and allow safe implementation.

The aim of this study was to conduct a systematic review to explore and summarise available evidence on POC(Hb)T use in children in LMICs. Using data from published literature and trial registries, we address the following questions: what, where and how are POC(Hb)Ts being used by health facilities in LMICs to diagnose childhood anaemia, and are there challenges to their use? To the best of our knowledge, our systematic review was the first to address these questions and therefore provides invaluable evidence for policymakers.

## Methods

### Literature search

We conducted a systematic review, reported in line with the Preferred Reporting Items for Systematic Reviews and Meta-Analyses (PRISMA) 2020 guidelines.^24^ We aimed to identify all published and unpublished literature using POC(Hb)Ts in children in LMICs. We identified published literature by searching six bibliographic databases: MEDLINE, EMBASE and Global Health via OVID, Web of Science, LILACS, and Cochrane Central Register of Controlled Trials. Clinical trial registries (WHO International Clinical Trials Registry Platform and ClinicalTrials.gov) were searched for unpublished trials. ProQuest Dissertations and Theses were searched for additional grey literature. Entire platforms were searched up to 10^th^ June 2022 and English language filters applied. Relevant journals, articles and authors were manually searched to identify missing literature; this included searching reference lists of included studies.

Search terms were based on four key concepts: anaemia, POC(Hb)T, LMIC and clinical setting. LMIC filters were provided by Cochrane Collaboration and updated according to World Banks Classification 2022.^25, 26^ Full details of the search strategy are outlined in Table S1.

### Selection criteria

We included all Clinical Trials, RCTs and observational studies using POC(Hb)Ts to diagnose anaemia in children attending health facilities. We restricted our review to children aged 0-20 years, and to literature published/registered within the last 15 years. Reviews, at-home POC Hb testing, testing from non-blood samples, high-income countries, and non-English or non-full text publications were excluded. We also excluded studies not performing POC(Hb)Ts immediately at the site of care (laboratory or delayed sample analysis) to provide representative evidence of their intended use. There were no restrictions based on child presentation or characteristics to ensure generalisability in the paediatric population.

### Data extraction

Results were exported to Endnote 20 and Rayyan systematic review management software. Duplicates were removed and further checked manually. The primary reviewer (RB) double-screened titles and abstracts for relevance using pre-specified inclusion/exclusion criteria. Potentially eligible studies were further screened by full-text assessment. Any uncertainties on study eligibility were discussed and resolved by consensus with co-authors (ECG, AS).

We extracted the following information: study characteristics, location, setting, POC(Hb)T(s) used, sample, prevalence of mild/moderate/severe/overall anaemia or mean Hb concentration, diagnostic accuracy and any challenges to test use reported by study authors. All data was collected using three piloted data extraction tools (study characteristics, challenges, and diagnostic accuracy) created in Microsoft Word (Table S2, S3 and Table 3).

**Table 3.**
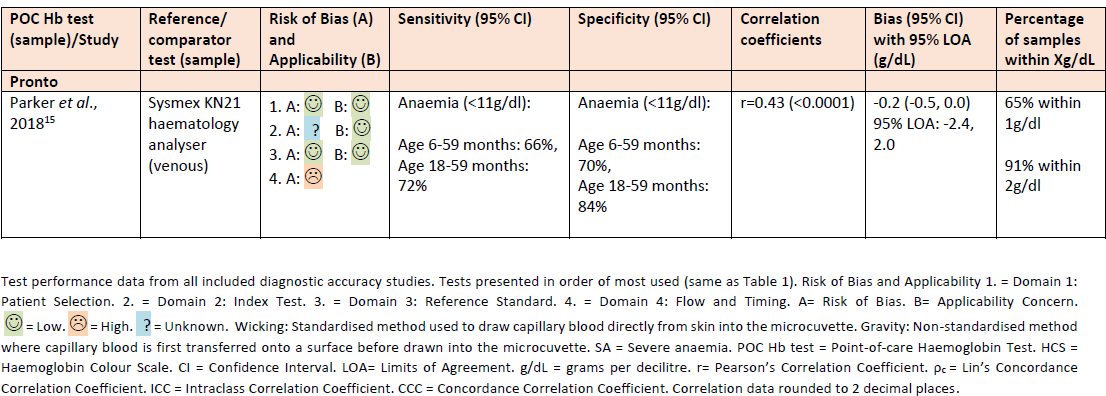
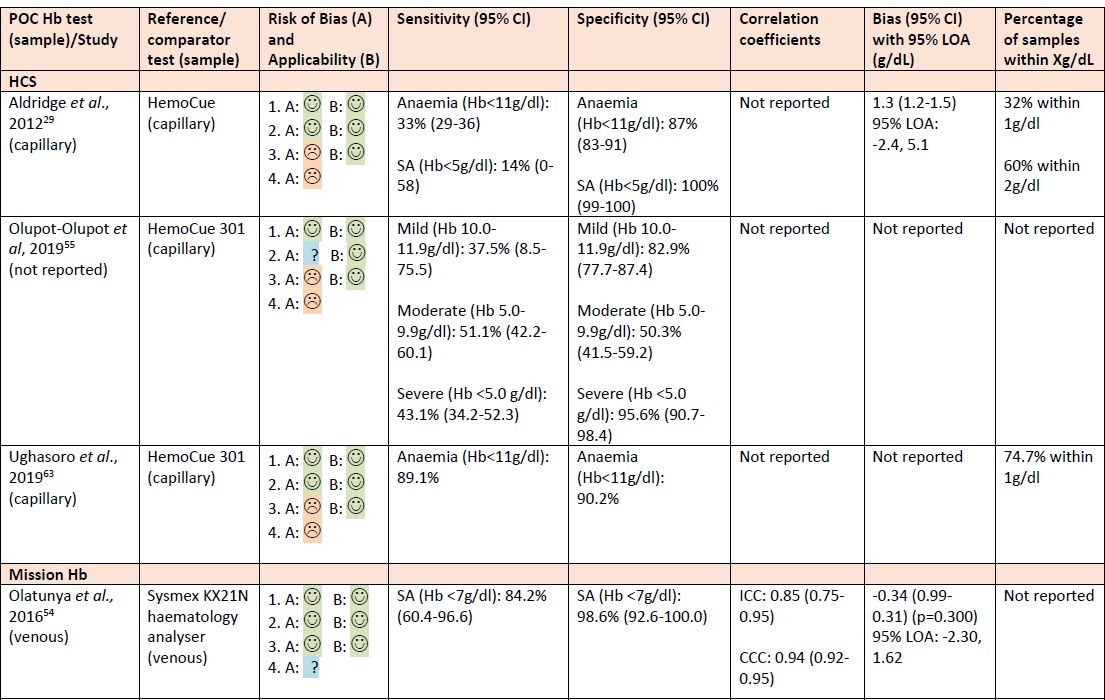
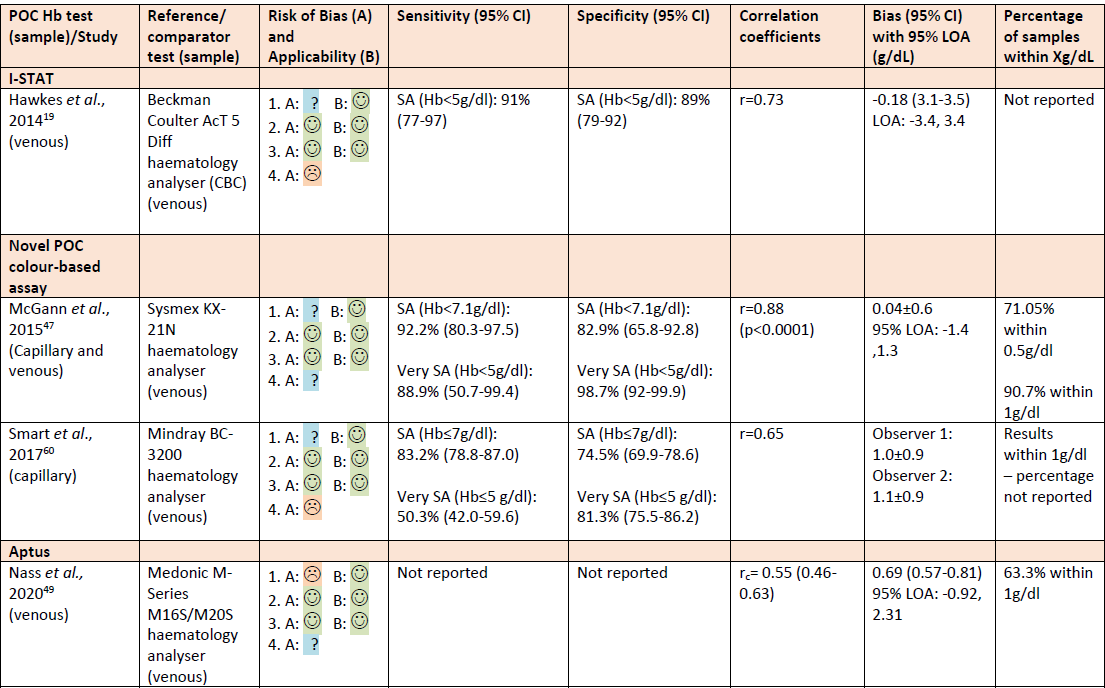
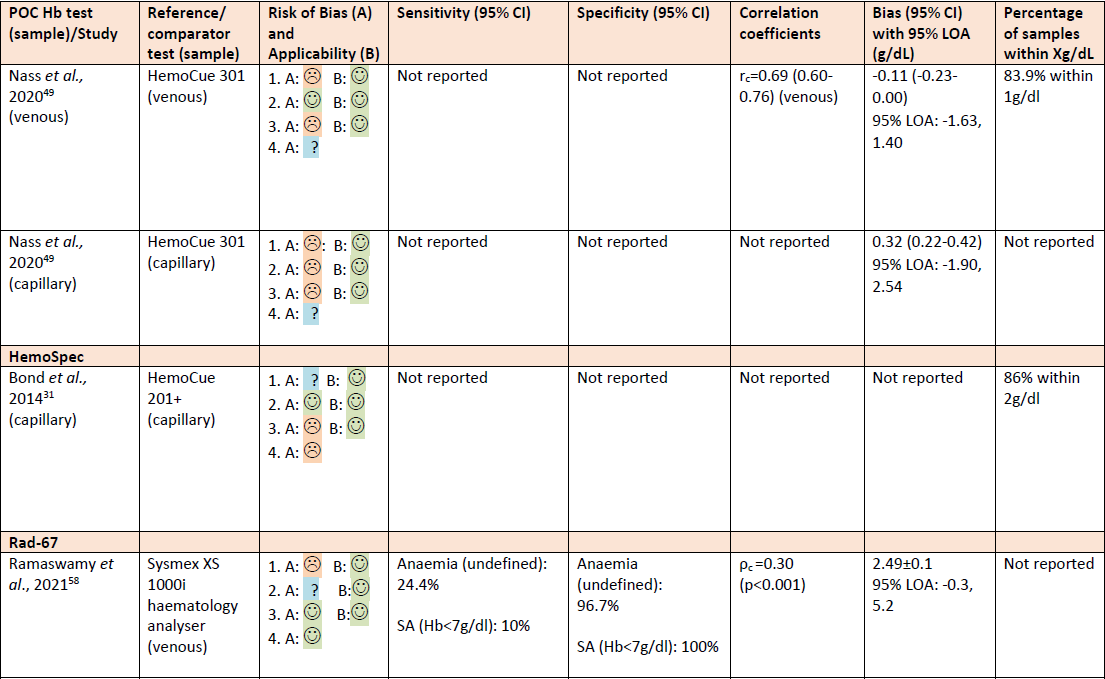
Diagnostic Accuracy of Included POC Hb Tests.

### Quality assessment

We assessed quality of included diagnostic studies to evaluate the reliability and validity of findings on POC(Hb)T performance. The primary reviewer (RB) assessed risk of bias (RoB) and applicability concerns using an adapted QUADAS-2 tool (Table S4).^27^ Due to limited evidence on HemoCue (HemoCue AB, Ängelholm, Sweden) device accuracy in this population and setting, studies using HemoCue as reference standard were judged high RoB for domain three. “Unclear” judgement was only made when insufficient evidence was reported.

### Data analysis

Extracted data was synthesised from all included studies and summarised in groups to answer the review question: study characteristics, POC(Hb)Ts, location/setting and challenges. Median and interquartile-range (IQR) were calculated for test sensitivity and specificity using Microsoft Excel. Due to differences in cut-offs used to define anaemia and severe anaemia across included studies and age groups, we summarised data according to how it was reported in papers (severe or overall anaemia) rather than using WHO definitions. For diagnostic studies, test sensitivity and specificity were therefore summarised by including cut-offs of Hb<5g/dl and Hb<7g/dl for diagnosis of severe anaemia. Meta-analysis was not conducted due to insufficient data available for individual POC(Hb)Ts.

## Results

### Study characteristics

742 records were identified from bibliographic databases and 24 from trial registries, presented in Figure 1. 179 duplicates were removed, resulting in title and abstract screening of 587 records. 439 records were excluded, leaving 148 records for full-text assessment. 39 records were also identified from reviewing related articles and citation lists. A total of 45 records from 41 studies met inclusion criteria and were included in our review. ^3, 5, 6, 13, 15, 19, 22, 28-65^ Reasons for exclusions are outlined in Figure 1.

**Figure 1.**
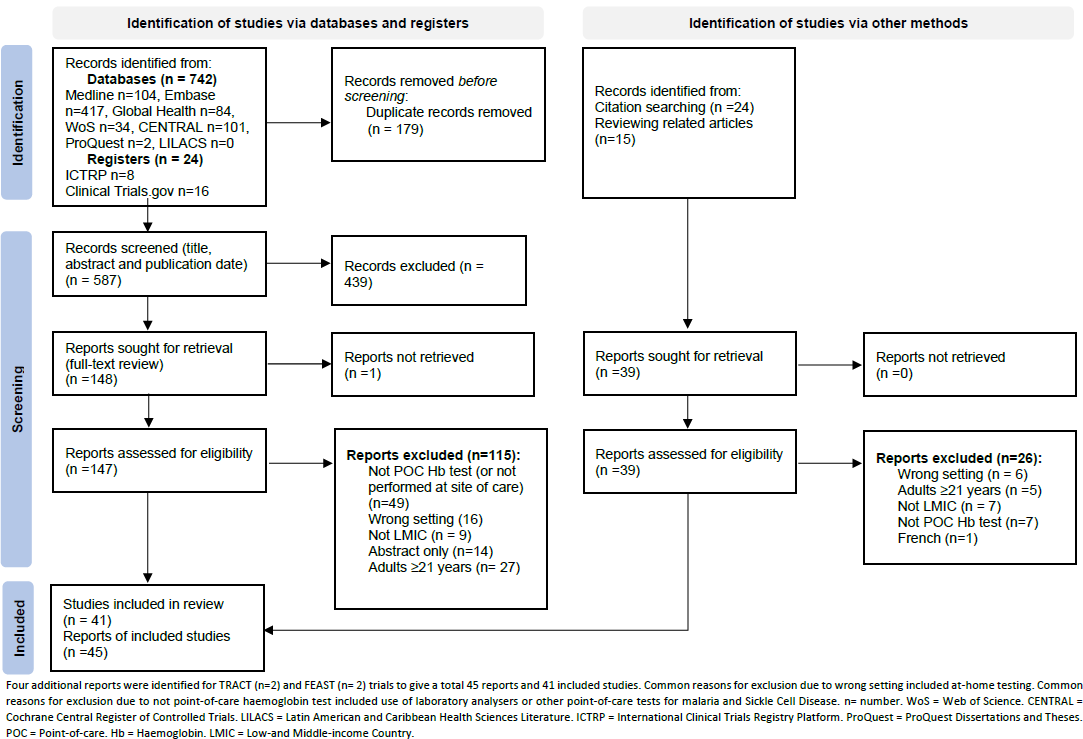
PRISMA 2020 Flow Diagram of Study Selection.

Of 41 included studies, 28 (68%) were observational (cross-sectional n=10, case-control n=3, cohort n=1, diagnostic n=14). 12 studies were RCTs (29%) and one study did not report study design.^48^ POC(Hb)Ts were implemented in 25 studies (61%) and assessed in 16 (39%). Of studies that assessed POC(Hb)Ts, 14 were diagnostic, one was retrospective and assessed user experiences of POC(Hb)T and one assessed use as part of an electronic algorithm.^13, 15, 19, 29, 31, 32, 42, 47, 49, 50, 54, 55, 58, 60, 62, 63^ Sample size ranged from 67-3,983 children undergoing POC Hb testing. Age ranged from 0-20 years with only six studies including older children (>15 years) (15%).^32, 35, 50, 51, 54, 60^ One study did not report specific age of included children.^47^ Four studies included adults and children.^32, 35, 50, 51^ Overall anaemia prevalence ranged from 11.9-100% (n=16). Prevalence of mild, moderate, and severe anaemia ranged from 2.7-52.2% (n=9), 2.5-60.2% (n=10) and 0.8-52.1% (n=14). Mean Hb concentration measured by POC(Hb)Ts ranged from 3.6-12.8g/dl (n=24). Full details of study characteristics are shown in Table S2.

### POC(Hb)Ts

A total of 15 different POC(Hb)Ts were used, presented in Table 1. 13 (87%) were invasive and required whole blood samples, of which 11 were electronic devices. Two were non-invasive electronic devices (13%). All devices were portable, calibrated and used either changeable or rechargeable batteries. All results were available within two minutes.

**Table 1.** Description and Summary of POC Hb Tests Used.

| POC Hb test | Principle method | Consumables | Hb detection range (g/dL) | Operating conditions (temperature; humidity) | Total studies (n) | Reported Study Setting |  |  |
| --- | --- | --- | --- | --- | --- | --- | --- | --- |
|  |  |  |  |  |  | Hospital (n) | Health Centre (n) | Clinic/ basic facilities (n) |
| **Total studies (n)** |  |  |  |  | **41\*** | **28\*** | **9\*** | **10\*** |
| **Invasive** |  |  |  |  |  |  |  |  |
| **HemoCue – Exact model not reported**<br>(HemoCue AB, Ängelholm, Sweden) |  |  |  |  | 12 (29%)<sup>3, 29, 30, 33, 34, 37, 40, 41, 43, 48, 59, 64</sup> | 7<sup>3, 33, 34, 37, 41, 48, 64</sup> | 1<sup>40</sup> | 5<sup>29, 30, 43, 59, 64</sup> |
| **HemoCue 301**<br>(HemoCue AB, Ängelholm, Sweden) | Absorbance at Hb/HbO<sub>2</sub> isosbestic point; transmittance measured at 506nm and 880nm | Reagent-free microcuvettes | 0-25.6 | 10-40°C; ≤90% | 11 (27%)<sup>5, 22, 28, 36, 38, 49, 55, 58, 61-63</sup> | 7<sup>5, 22, 28, 36, 38, 55, 58</sup> | 3<sup>36, 62, 63</sup> | 2<sup>49, 61</sup> |
| **HemoCue 201+**<br>(HemoCue AB, Ängelholm, Sweden) | Modified azidemethaemoglobin reaction; absorbance measured at 570nm and 880nm | Reagent-containing microcuvettes | 0-25.6 | 15-30°C; ≤90% | 4 (10%)<sup>15, 39, 42, 52</sup> | 3<sup>15, 42, 52</sup> | 2<sup>39, 42</sup> | 1<sup>39</sup> |
| **HCS (COPACK, Oststeinbek, Germany)** | Absorbance of blood on chromatography paper and colour compared to a scale of six shades of red representing Hb levels at 2g/dl intervals | Filter paper test strips | 4-14<br>(Scale at 4,6,8,10,12, and 14) | Not known | 4 (10%)<sup>29, 55, 63, 65</sup> | 1<sup>55</sup> | 1<sup>63</sup> | 2<sup>29, 65</sup> |

**Table 1. Description and Summary of POC Hb Tests Used.**
| POC Hb test | Principle method | Consumables | Hb detection range (g/dL) | Operating conditions (temperature; humidity) | Total studies (n) | Reported Study Setting |  |  |
| --- | --- | --- | --- | --- | --- | --- | --- | --- |
|  |  |  |  |  |  | Hospital (n) | Health Centre (n) | Clinic/ basic facilities (n) |
| **Mission Hb** (ACON Laboratories, Inc., San Diego, USA) | Modified azidemethaemoglobin reaction measured by reflectance photometry | Reagent-containing test strips, micropipette | 4.5-25.6 | 10-40°C; ≤90% | 3 (7%)<sup>32, 53, 54</sup> | 3 | 0 | 0 |
| **URIT-12** (URIT Medical Electronics, Guangxi, China) | Modified azidemethaemoglobin reaction measured by reflectance photometry | Reagent-containing test strips | 4-24 | 15-30°C; ≤80% | 3 (7%)<sup>32, 51, 57</sup> | 2<sup>32, 51</sup> | 1<sup>51</sup> | 1<sup>57</sup> |
| **I-STAT** (Abbott, Abbott Park, IL) | Measures conductivity and corrects for electrolyte concentration to estimate HCT. Hb then calculated using the formula: Hb (g/dl) = HCT (% packed cell volume) x0.34 | Cartridges (EC8+ and CHEM8+) | 5.1-25.5 | 16-30°C; ≤90% | 2 (5%)<sup>19, 37</sup> | 2 | 0 | 0 |
| **HemoCue 201** (HemoCue AB, Ängelholm, Sweden) | Modified azidemethaemoglobin reaction; absorbance measured at 570nm and 880nm | Reagent-containing microcuvettes | 0-25.6 | 18-30°C; ≤90% | 2 (5%)<sup>35, 65</sup> | 0 | 1<sup>35</sup> | 1<sup>65</sup> |
| **Novel colour-based assay** | Reaction between Hb, hydrogen peroxide and 3,3',5,5'-TMB that produces a colour change according to Hb concentration: | Reagent tubes and 10µl capillary tube | 2.5-9.1 | Not known | 2 (5%)<sup>47, 60</sup> | 2 | 0 | 0 |

**Table 1. Description and Summary of POC Hb Tests Used.**
| POC Hb test | Principle method | Consumables | Hb detection range (g/dL) | Operating conditions (temperature; humidity) | Total studies (n) | Reported Study Setting |  |  |
| --- | --- | --- | --- | --- | --- | --- | --- | --- |
|  |  |  |  |  |  | Hospital (n) | Health Centre (n) | Clinic/ basic facilities (n) |
|  | blue (Hb<3g/dl), blue/green (Hb 3-5g/dl), yellow/orange (Hb 5-7g/dl), orange/red (Hb 7-9g/dl) to red (Hb >9g/dl)<sup>47, 60</sup> |  |  |  |  |  |  |  |
| **HemoCue 801** (HemoCue AB, Ängelholm, Sweden) | Absorbance of whole blood at Hb/HbO<sub>2</sub> isosbestic point; transmittance measured at 506nm and 880nm | Reagent-free microcuvettes | 1-25.6 | 10-40°C; ≤90% (25°C), ≤75% (40°C) | 1 (2%)<sup>13</sup> | 1 | 0 | 0 |
| **HemoCue B-haemoglobin** (HemoCue AB, Ängelholm, Sweden) | Modified azidemethaemoglobin reaction; absorbance measured at 570nm and 880nm | Reagent-containing microcuvettes | 0-25.6 | 15-30°C | 1 (2%)<sup>50</sup> | 1 | 0 | 0 |
| **Aptus** (Entia, London, UK) | Centrifugation and photometry at 515nm, 660nm and 940nm to estimate HCT level. Hb calculated using MCHC x HCT | Microcuvettes | 5-25 | 5-45°C; ≤90% | 1 (2%)<sup>49</sup> | 0 | 0 | 1 |
| **HemoSpec** | Modified azidemethaemoglobin reaction; absorbance measured at 532nm and 650nm | Reagent-containing chromatography paper | Not known | Not known | 1 (2%)<sup>31</sup> | 1 | 0 | 0 |

**Table 1. Description and Summary of POC Hb Tests Used.**
| POC Hb test | Principle method | Consumables | Hb detection range (g/dL) | Operating conditions (temperature; humidity) | Total studies (n) | Reported Study Setting |  |  |
| --- | --- | --- | --- | --- | --- | --- | --- | --- |
|  |  |  |  |  |  | Hospital (n) | Health Centre (n) | Clinic/ basic facilities (n) |
| **HemoControl** (EKF Diagnostics, Cardiff, UK) | Modified azidemethaemoglobin reaction; absorbance measured at 570nm and 880nm | Reagent-containing microcuvettes | 0-25.6 | 15-40°C; <90% | 1 (2%)<sup>56</sup> | 1 | 1 | 1 |
| **Non-invasive** |  |  |  |  | **2** | **2** | **0** | **0** |
| **Rad-67™ Pulse CO-Oximeter® and rainbow® DCI®-mini-Sensor** (Masimo Corporation, Irvine, USA) | Visible and infrared lights (500-1400nm) signalled through capillary bed and sensor detects changes in light absorption. Hb then calculated using a multi-wavelength calibration equation | None | 8-17 | 0-35°C; 10-95% | 1 (2%)<sup>58</sup> | 1 | 0 | 0 |
| **Pronto® device with DCI-mini™ sensors** (Masimo Corporation, Irvine, USA) | Visible and infrared lights (500- 1300nm) signalled through capillary bed and sensor detects changes in light absorption. Hb then calculated using a multi-wavelength calibration equation | None | 8-17 | 5-40°C; 5-95% | 1 (2%)<sup>15</sup> | 1 | 0 | 0 |

Tests presented in order of most used (number of studies). \* Six studies were conducted across several settings and nine studies used more than one test and so total number in columns does not equal total number of studies or 100%. HCT = Haematocrit. MCHC = Mean Corpuscular Haemoglobin Concentration. n= Number of studies. Hb = Haemoglobin. HbO2 = Oxyhaemoglobin. IL = Illinois. UK = United Kingdom. USA = United States of America. nm = Nanometre. µl = Microlitre. g/dl = Grams per decilitre. °C = Degrees Celsius.

HemoCue devices were the most used test (n=31, 76%) and of papers reporting the specific model used was HemoCue301 (n=11, 27%). Six studies used colour-based tests (Haemoglobin Colour Scale (HCS) (COPACK, Germany) n=4, novel-assay n=2).^29, 47, 55, 60, 63, 65^ Nine studies used multiple tests.^15, 29, 32, 37, 49, 55, 58, 63, 65^ Of studies using invasive tests (n=41), most used capillary sampling (n=19). Eight studies used venous sampling and five studies used both capillary and venous samples. Nine studies did not report the sample type used.^3, 22, 30, 34, 36, 42, 48, 53, 65^ All invasive tests, excluding I-STAT (Abbott Park, Illinois) required a sample volume of 15μl or less (Table S5).

### Location and Setting

Most studies were conducted in Sub-Saharan Africa (n=39, 95%), across 14 African countries, presented in Figure 2. Two trials included more than one Sub-Saharan country.^5, 22^ One study was conducted in Asia (India) and one in South America (Brazil).^58, 59^

28 studies were reported as conducted in hospitals (68%). Nine studies were reported as conducted in health centres (22%).^35, 36, 39, 40, 42, 51, 56, 62, 63^ Eight studies were reported as conducted in health clinics (20%). ^29, 30, 39, 43, 49, 57, 61, 64^ Two studies were reported as conducted in basic health facilities (5%).^59, 65^ Six studies were multifacility and were reported as conducted in hospitals, health centres and/or clinics.^36, 39, 42, 51, 56, 64^ One study included a dispensary.^56^ Health facilities were reported as rural in seven studies.^3, 30, 43, 48, 49, 62, 63^

**Figure 2.**
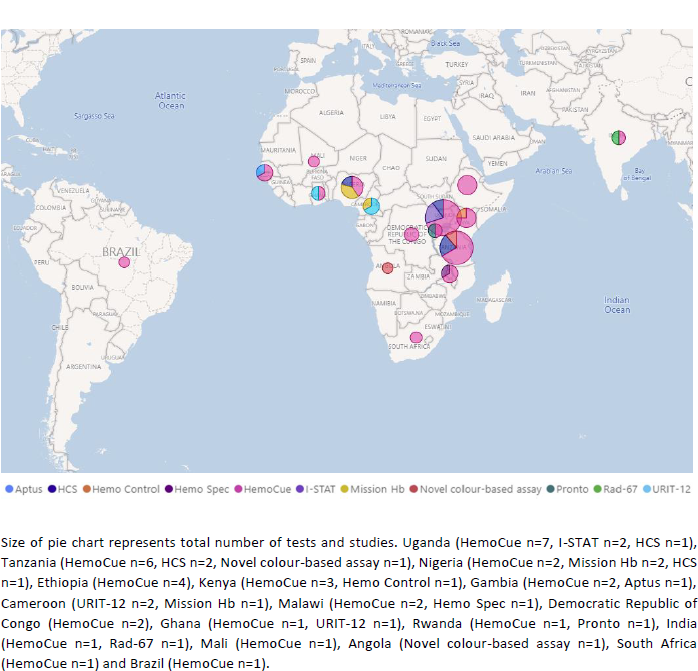
Map Showing where POC Hb Tests have been used by Health Facilities in Children in LMICs.

Use of POC(Hb)Ts varied across included studies: to assess diagnostic accuracy (n=14, 34%), assess anaemia prevalence or associated factors/infections (n=13, 32%), assess Hb levels during intervention follow-up (n=6, 15%), assist triage of sick children (n=3, 7%), guide and monitor transfusion in children with severe anaemia (n=3, 7%), assess trial eligibility (n=1, 2%) and assess effect of anaemia on school performance (n=1, 2%).

### Challenges

13 studies (32%) reported challenges to POC(Hb)T use. ^13, 15, 19, 29, 31, 32, 47, 49, 50, 54, 55, 58, 60^ Overestimation of Hb concentration (n=5) and errors in sampling and environmental factors (n=5) were the most frequently reported challenges. Other challenges included cost of device, consumables and training, difficulty in obtaining measurement, inter-observer variability, supply and stability of consumables, maintenance, and device failure, shown in Table 2. Diagnostic accuracy data were available for 10 POC(Hb)Ts in 11 of 14 diagnostic studies, shown in Table 3.^15, 19, 29, 31, 47, 49, 54, 55, 58, 60, 63^ Accuracy data was not extracted in three studies due to pooled data with adults and children (n=2) and use of a non Hb-measuring tool (n=1).^32, 50, 62^ Five studies reported accuracy results were clinically unacceptable, of which one included adults and children (Table 2).^15, 29, 31, 32, 49^ Test performance varied across individual studies. Sensitivity to detect anaemia and severe anaemia ranged from 24.4-92.2% (median 74%, IQR:32.8, n=4) and 10-92.2% (median 83.7%, IQR:54.4, n=7), respectively. This variability decreased (64-92.2% (n=2) and 84.2-91% (n=3)) when limited to invasive devices.^15, 19, 54, 58^ Specificity to detect anaemia and severe anaemia ranged from 70-96.7% (median 84.7%, IQR:6.1 n=4) and 74.5-100% (median 93.2%, IQR:16.5, n=7), respectively.

**Table 2.**
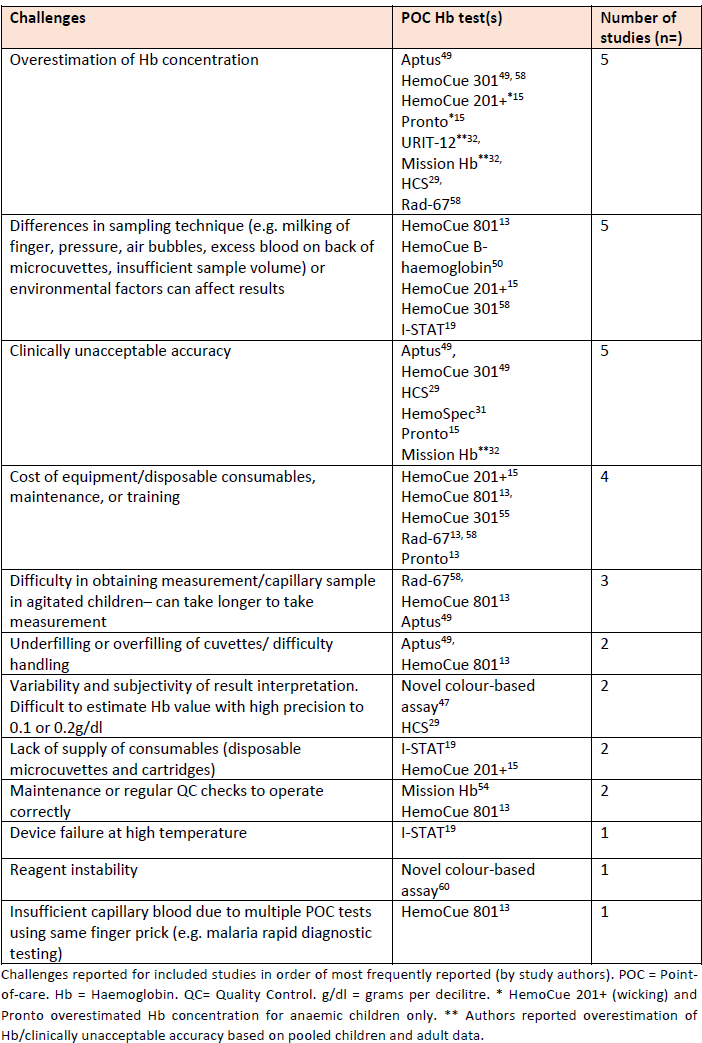
Summary of key Challenges Reported.

| Challenges | POC Hb test(s) | Number of studies (n=) |
| --- | --- | --- |
| Overestimation of Hb concentration | Aptus<sup>49</sup><br>HemoCue 301<sup>49, 58</sup><br>HemoCue 201+<sup>*15</sup><br>Pronto<sup>*15</sup><br>URIT-12<sup>**32,</sup><br>Mission Hb<sup>**32,</sup><br>HCS<sup>29,</sup><br>Rad-67<sup>58</sup> | 5 |
| Differences in sampling technique (e.g. milking of finger, pressure, air bubbles, excess blood on back of microcuvettes, insufficient sample volume) or environmental factors can affect results | HemoCue 801<sup>13</sup><br>HemoCue B-haemoglobin<sup>50</sup><br>HemoCue 201+<sup>15</sup><br>HemoCue 301<sup>58</sup><br>I-STAT<sup>19</sup> | 5 |
| Clinically unacceptable accuracy | Aptus<sup>49,</sup><br>HemoCue 301<sup>49</sup><br>HCS<sup>29</sup><br>HemoSpec<sup>31</sup><br>Pronto<sup>15</sup><br>Mission Hb<sup>**32</sup> | 5 |
| Cost of equipment/disposable consumables, maintenance, or training | HemoCue 201+<sup>15</sup><br>HemoCue 801<sup>13,</sup><br>HemoCue 301<sup>55</sup><br>Rad-67<sup>13, 58</sup><br>Pronto<sup>13</sup> | 4 |
| Difficulty in obtaining measurement/capillary sample in agitated children– can take longer to take measurement | Rad-67<sup>58,</sup><br>HemoCue 801<sup>13</sup><br>Aptus<sup>49</sup> | 3 |
| Underfilling or overfilling of cuvettes/ difficulty handling | Aptus<sup>49,</sup><br>HemoCue 801<sup>13</sup> | 2 |
| Variability and subjectivity of result interpretation. Difficult to estimate Hb value with high precision to 0.1 or 0.2g/dl | Novel colour-based assay<sup>47</sup><br>HCS<sup>29</sup> | 2 |
| Lack of supply of consumables (disposable microcuvettes and cartridges) | I-STAT<sup>19</sup><br>HemoCue 201+<sup>15</sup> | 2 |
| Maintenance or regular QC checks to operate correctly | Mission Hb<sup>54</sup><br>HemoCue 801<sup>13</sup> | 2 |
| Device failure at high temperature | I-STAT<sup>19</sup> | 1 |
| Reagent instability | Novel colour-based assay<sup>60</sup> | 1 |
| Insufficient capillary blood due to multiple POC tests using same finger prick (e.g. malaria rapid diagnostic testing) | HemoCue 801<sup>13</sup> | 1 |

Challenges reported for included studies in order of most frequently reported (by study authors). POC = Point-of-care. Hb = Haemoglobin. QC= Quality Control. g/dl = grams per decilitre. \* HemoCue 201+ (wicking) and Pronto overestimated Hb concentration for anaemic children only. \*\* Authors reported overestimation of Hb/clinically unacceptable accuracy based on pooled children and adult data.

Mean difference/bias between POC(Hb)Ts and reference ranged from −0.34 to 2.49g/dl (n=8), with lower and upper limits of agreement (LOA) ranging from −0.17 to −3.4g/dl and 1.3 to 5.2g/dl, respectively (n=7). Five studies reported percentage of Hb values within 1g/dl of reference standard.^15, 29, 47, 49, 63^

### Quality assessment

We detected high RoB for at least one domain in nine of 11 diagnostic studies (Table 3).^15, 19, 29, 31, 49, 55, 58, 60, 63^ No studies showed concerns for applicability. Incomplete reporting resulted in unclear judgement for one or more domains in nine studies.^15, 19, 31, 47, 49, 54, 55, 58, 60^ Flow and Timing domain showed the greatest proportion of high RoB due to some enrolled patients missing from final analyses.^15, 19, 29, 31, 55, 60, 63^ Five studies were judged high RoB for domain three due to HemoCue as comparator/reference method.^29, 31, 49, 55, 63^ The proportion of studies with low, high, or unclear RoB and reasons for each judgement are presented in Figure S1 and Table S6.

## Discussion

We systematically reviewed the literature to explore POC(Hb)T use in children in LMICs. Using data from published literature and trial registries, we found evidence on the use of 15 different POC(Hb)Ts by health facilities across 16 LMICs in the last 15 years to diagnose childhood anaemia; from 41 studies. 39 studies (95%) were conducted in Africa, indicating little evidence is available outside this region. We found that, to date, HemoCue is the most widely utilised test in this population and setting. Our results represent a relatively small proportion of commercially available and assessed POC(Hb)Ts, suggesting limited evidence is available in children.^17, 66-70^ Imperfect diagnostic accuracy, sampling errors, environmental factors and costs were key challenges reported for POC(Hb)T use.

Five POC(Hb)Ts (33%) could not detect Hb levels below 4g/dl. This is important because many children with severe anaemia in LMICs have Hb<4g/dl.^3, 71^ Three studies in our review reported mean Hb concentration by POC(Hb)T <4g/dl and five additional studies reported Hb values below this level.^3, 5, 33, 37, 44, 47, 50, 55^ The WHO guidelines for the management of children with severe anaemia and the novel transfusion algorithm developed based on results from TRACT use a cut-off of Hb<4g/dl to determine which children require transfusion in the absence of other severity signs.^23, 72^ Therefore, Mission Hb (ACON Laboratories, USA), I-STAT, Aptus (Entia, UK), Rad-67 (Masimo, USA) and Pronto (Masimo, USA) are not currently suitable for identifying severe anaemia in this population. It is worth noting however, that I-STAT, Rad-67 and Pronto measure additional parameters besides Hb and haematocrit, increasing their utility. URIT-12 (URIT, China) and HCS have a Hb cut-off of 4g/dl and so accuracy detection at this level must be further investigated.

We identified several challenges to POC(Hb)T use that must be addressed before wider implementation. No POC(Hb)T showed excellent diagnostic accuracy across all measurements and therefore may not meet all ASSURED criteria.^12^ Five studies reported overestimation of Hb concentration in eight POC(Hb)Ts (Table 2).^15, 29, 32, 49, 58^ This is vital since it could result in misclassification of severity of anaemia and therefore prevent truly eligible children from receiving a lifesaving transfusion or appropriate treatment. In contrast, underestimation of Hb levels by POC(Hb)Ts has been reported in children, causing unnecessary use of scarce resources and exposure of children to transfusion-related risks.^73, 74^ A previous review of HemoCue, found underestimation of Hb most frequently reported, with few studies reporting overestimation of Hb concentration.^75^ These conflicting findings could be explained by variations in child Hb level.^15, 76-78^

Similarly, wide LOA means estimated Hb values could span all categories of anaemia. Our results show seven diagnostic studies and eight POC(Hb)Ts exceeded the clinically acceptable accuracy of upper and lower LOA within 1g/dl (Table 3).^15, 19, 29, 47, 49, 54, 58^ This suggests within-subject variation is too large to provide a clinically meaningful diagnosis. These results are in line with a previous systematic review including adults and children in mixed settings.^79^ However, a different review has shown clinically acceptable LOA.^80^ Clinicians should be aware of these LOA when basing clinical decision solely on these Hb estimations. We also found substantial variation in test sensitivity to detect anaemia and severe anaemia across included studies, with lowest values reported for HCS and Rad-67.^29, 58^ This is contrary to a previous review that identified lowest sensitivity to detect anaemia for HemoCue301 (22.6%) and highest sensitivity for HCS (99.3%) out of 6 POC(Hb)Ts.^81^ Our median values for test sensitivity and specificity to detect anaemia were moderate, suggesting some children would receive a false negative result.

Variation in test performance across studies in our review and with previous literature has several possible explanations. Firstly, nine studies in our review were judged high RoB for at least one domain. ^49, 58 15, 19, 29, 31, 55, 60, 63^ Differences in anaemia prevalence, geographical factors (temperature, altitude, and humidity), reference test, transportation and storage of consumables, sampling technique and training could also explain discrepancy.^15, 82, 83^ Three studies did not use thresholds for severe anaemia defined by WHO for the assessment of sensitivity and specificity (Hb<5g/dl), limiting evidence synthesis and contributing to disparity.^19, 29, 55^ Lastly, two included studies used different sources of sample for the reference method and POC(Hb)T, and five included diagnostic studies used venous samples for POC(Hb)T.^15, 19, 47, 49, 54, 58, 60^ This could explain variation due to known differences in capillary and venous blood.^84^

We highlight that these real-life factors pose a challenge to POC(Hb)T use. There is a need for standardised training protocols to reduce errors in sampling technique and interpretation of colour-based tests. Competency of staff and therefore performance should increase as these tests become routine practice. Our findings also suggest five POC(Hb)Ts used may not be suitable for use in some LMICs due to possible device failure at high temperature (>30°C) (Table 1).^15, 19, 32, 35, 39, 42, 50-52, 57^

Moreover, we identified analyser costs and lack of supply of consumables as challenges to POC(Hb)T use. Although upfront costs are relatively high, particularly for non-invasive and HemoCue devices, it is the recurrent, per test costs that pose an obstacle to sustained use in LMICs and could explain the lack of supply of microcuvettes and cartridges.^85^ Total cost of POC tests must be weighed against their benefits such as earlier diagnosis, reduced morbidity and mortality, improved patient satisfaction and decreased cost of unnecessary referrals, and additional testing.^86^ Use of POC(Hb)Ts with electronic decision support algorithms could enhance their cost-effectiveness in triage of sick children. However, only one included study adopted this approach and therefore this warrants further research.^42^ The novel colour-based assay offers a significant cost advantage at 0.26USD per test, however further evidence on its use is required. Other affordable technologies, such as smartphone-based colorimetry are at early stages of development for identifying severe anaemia in LMICs.^87^

Key strengths of our review were wide inclusion criteria and adherence to systematic review methods. This allowed a novel and comprehensive synthesis of POC(Hb)T use in this population and setting. Ongoing trials were included and therefore reduced publication bias. Furthermore, we assessed RoB and applicability of included diagnostic studies to evaluate the reliability and validity of findings on POC(Hb)T performance.

Our review is not without limitations. Firstly, a single reviewer (RB) screened results, adapted QUADAS-2 tool and judged study inclusion eligibility and RoB and applicability concerns, introducing bias. However, records were screened twice to minimise risk of missing relevant articles and any uncertainties were resolved by consensus with co-authors (ECG, AS). Secondly, we searched a restricted number of databases and trial registries; however this risk was minimised by searching the largest and most renowned databases, specific to the subject area. Other methodological limitations include exclusion of reviews, non-full-text or non-English articles and studies published over 15 years ago. We also note, only 13 studies (32%) reported challenges to POC(Hb)T use and therefore our review may be subject to reporting bias if not all challenges were reported. Lastly, we only assessed data from published literature and trial registries. Clinical trials/studies may not represent the total use of POC(Hb)Ts and may be biased by trial funding and supply of tests and consumables.

### Further research

We found that diagnostic accuracy data for this population and setting was available for only 10 POC(Hb)Ts. This finding is critical, since POC(Hb)Ts must be validated in the population and setting of their intended use before wider adoption. Further high-quality research on test accuracy, particularly using capillary samples is therefore warranted to assess performance in various field settings and address discrepancy between studies. There is currently insufficient data to conduct meta-analysis on individual POC(Hb)Ts in this population and setting. Further research should therefore ensure data and results can be combined with previous studies for meta-analysis.

Our findings also show very few studies used POC(Hb)Ts in guiding and monitoring transfusion.^5, 22, 37^ Therefore, further research is essential to evaluate use with the novel transfusion algorithm and impact on patient-centred outcomes, time to transfusion and usage of blood supply. Further research is also required to understand the clinical and resource implications of under/overestimating Hb levels, especially near the cut-off for severe anaemia. The challenges identified in our review also stress the need for further development of some POC(Hb)Ts and standardised training procedures. For example, by expanding detection ranges to include lower Hb levels and also to enable higher operating temperatures.

## Conclusions

In conclusion, 15 POC(Hb)Ts have been successfully utilised in all-levels of health facilities across 16 LMICs to diagnose childhood anaemia of various aetiologies. However, several challenges to their use exist and must be addressed before wider implementation. We found HemoCue301, HemoCue801 and HemoControl (EKF Diagnostics, UK) offer the most suitable Hb detection ranges and operating temperatures (<40°C) for use in this setting. However, we identified no evidence on diagnostic accuracy for HemoCue801 and HemoControl. We therefore recommend HemoCue301 as the best available POC(Hb)T to diagnose childhood anaemia in LMICs, based on available evidence. However, imperfect diagnostic accuracy is a drawback and must be weighed against benefits in costs, safety, convenience, and improved clinical outcome. Further research is essential to confirm these benefits and diagnostic accuracy in field settings. Routine use of POC(Hb)Ts may significantly reduce child mortality in LMICs, where laboratory analysers are often unavailable and anaemia prevalence is high.

## Data Availability

All data produced in the present work are contained in the manuscript.

## References

1. Safiri S, Kolahi AA, Noori M, Nejadghaderi SA, Karamzad N, Bragazzi NL, et al. Burden of anemia and its underlying causes in 204 countries and territories, 1990-2019: results from the Global Burden of Disease Study 2019. J Hematol Oncol. 2021;14(1):185-.

2. World Health Organisation (WHO). Haemoglobin concentrations for the diagnosis of anaemia and assessment of severity. Geneva: World Health Organization; 2011.

3. Calis JC, Phiri KS, Faragher EB, Brabin BJ, Bates I, Cuevas LE, et al. Severe anemia in Malawian children. Malawi Med J. 2016;28(3):99–107.

4. Sun J, Wu H, Zhao M, Magnussen CG, Xi B. Prevalence and changes of anemia among young children and women in 47 low- and middle-income countries, 2000-2018. EClinicalMedicine. 2021;41:101136.

5. Maitland K, Kiguli S, Opoka RO, Engoru C, Olupot-Olupot P, Akech SO, et al. Mortality after fluid bolus in African children with severe infection. N Engl J Med. 2011;364(26):2483–95.

6. Kiguli S, Maitland K, George EC, Olupot-Olupot P, Opoka RO, Engoru C, et al. Anaemia and blood transfusion in African children presenting to hospital with severe febrile illness. BMC Med. 2015;13:21-.

7. Barnes LS, Stanley J, Bloch EM, Pagano MB, Ipe TS, Eichbaum Q, et al. Status of hospital-based blood transfusion services in low-income and middle-income countries: a cross-sectional international survey. BMJ Open. 2022;12(2):e055017.

8. Shari CR, Sawe HR, Murray BL, Mwafongo VG, Mfinanga JA, Runyon MS. Emergency blood transfusion practices among anaemic children presenting to an urban emergency department of a tertiary hospital in Tanzania. BMC Hematol. 2017;17:19.

9. Opoka RO, Ssemata AS, Oyang W, Nambuya H, John CC, Tumwine JK, et al. High rate of inappropriate blood transfusions in the management of children with severe anemia in Ugandan hospitals. BMC Health Serv Res. 2018;18(1):566.

10. Uyoga S, George E, Bates I, Olupot-Olupot P, Chimalizeni Y, Molyneux E, et al. Point-of-care haemoglobin testing in African hospitals: a neglected essential diagnostic test. British Journal of Haematology. 2021;193.

11. Nabwera HM, Fegan G, Shavadia J, Denje D, Mandaliya K, Bates I, et al. Pediatric blood transfusion practices at a regional referral hospital in Kenya. Transfusion. 2016;56(11):2732–8.

12. Peeling RW, Holmes KK, Mabey D, Ronald A. Rapid tests for sexually transmitted infections (STIs): the way forward. Sex Transm Infect. 2006;82 Suppl 5(Suppl 5):v1-6.

13. Tack B, Vita D, Mansosa I, Mbaki TN, Wasolua N, Luyindula A, et al. Field Experiences with Handheld Diagnostic Devices to Triage Children under Five Presenting with Severe Febrile Illness in a District Hospital in DR Congo. Diagnostics (Basel). 2022;12(3).

14. Yadav K, Kant S, Ramaswamy G, Ahamed F, Vohra K. Digital Hemoglobinometers as Point-of-Care Testing Devices for Hemoglobin Estimation: A Validation Study from India. Indian journal of community medicine : official publication of Indian Association of Preventive & Social Medicine. 2020;45(4):506–10.

15. Parker M, Han Z, Abu-Haydar E, Matsiko E, Iyakaremye D, Tuyisenge L, et al. An evaluation of hemoglobin measurement tools and their accuracy and reliability when screening for child anemia in Rwanda: A randomized study. PloS one. 2018;13(1):e0187663.

16. Chutipongtanate A, Yasaeng C, Virankabutra T, Chutipongtanate S. Systematic comparison of four point-of-care methods versus the reference laboratory measurement of hemoglobin in the surgical ICU setting: a cross-sectional method comparison study. BMC Anesthesiol. 2020;20(1):92-.

17. Neogi SB, Negandhi H, Kar R, Bhattacharya M, Sen R, Varma N, et al. Diagnostic accuracy of haemoglobin colour strip (HCS-HLL), a digital haemoglobinometer (TrueHb) and a non-invasive device (TouchHb) for screening patients with anaemia. J Clin Pathol. 2016;69(2):164–70.

18. Kuusipalo H, Maleta K, Briend A, Manary M, Ashorn P. Growth and change in blood haemoglobin concentration among underweight Malawian infants receiving fortified spreads for 12 weeks: a preliminary trial. J Pediatr Gastroenterol Nutr. 2006;43(4):525–32.

19. Hawkes M, Conroy AL, Opoka RO, Namasopo S, Liles WC, John CC, et al. Performance of point-of-care diagnostics for glucose, lactate, and hemoglobin in the management of severe malaria in a resource-constrained hospital in Uganda. The American journal of tropical medicine and hygiene. 2014;90(4):605–8.

20. Molla A, Egata G, Mesfin F, Arega M, Getacher L. Prevalence of Anemia and Associated Factors among Infants and Young Children Aged 6-23 Months in Debre Berhan Town, North Shewa, Ethiopia. J Nutr Metab. 2020;2956129-.

21. Leepile TT, Mokomo K, Bolaane MMM, Jones AD, Takada A, Black JL, et al. Anemia Prevalence and Anthropometric Status of Indigenous Women and Young Children in Rural Botswana: The San People. Nutrients. 2021;13(4).

22. Maitland K, Kiguli S, Olupot-Olupot P, Engoru C, Mallewa M, Saramago Goncalves P, et al. Immediate Transfusion in African Children with Uncomplicated Severe Anemia. N Engl J Med. 2019;381(5):407–19.

23. Maitland K, Kiguli S, Olupot-Olupot P, Opoka RO, Chimalizeni Y, Alaroker F, et al. Transfusion management of severe anaemia in African children: a consensus algorithm. Br J Haematol. 2021;193(6):1247–59.

24. Page MJ, McKenzie JE, Bossuyt PM, Boutron I, Hoffmann TC, Mulrow CD, et al. The PRISMA 2020 statement: an updated guideline for reporting systematic reviews. BMJ. 2021;372:n71.

25. World Bank Country and Lending Groups 2022. Available from: https://datahelpdesk.worldbank.org/knowledgebase/articles/906519-world-bank-country-and-lending-groups. [Accessed 20 April 2022].

26. Cochrane Effective Practice and Organisation of Care (EPOC) LMIC Filters. 2020 [Available from: https://epoc.cochrane.org/lmic-filters. [Accessed 20 April 2022].

27. Whiting PF, Rutjes AW, Westwood ME, Mallett S, Deeks JJ, Reitsma JB, et al. QUADAS-2: a revised tool for the quality assessment of diagnostic accuracy studies. Ann Intern Med. 2011;155(8):529–36.

28. Alamneh YM, Akalu TY, Shiferaw AA, Atnaf A. Magnitude of anemia and associated factors among children aged 6-59 months at Debre Markos referral hospital, Northwest Ethiopia: a hospital-based cross-sectional study. Italian journal of pediatrics. 2021;47(1):172.

29. Aldridge C, Foster HME, Albonico M, Ame SM, Montresor A. Evaluation of the diagnostic accuracy of the Haemoglobin Colour Scale to detect anaemia in young children attending primary healthcare clinics in Zanzibar. Tropical Medicine and International Health. 2012;17(4):423–9.

30. Bojang KA, Akor F, Conteh L, Webb E, Bittaye O, Conway DJ, et al. Two strategies for the delivery of IPTc in an area of seasonal malaria transmission in the Gambia: A randomised controlled trial. PLoS Medicine. 2011;8(2):e1000409.

31. Bond M, Mvula J, Molyneux E, Richards-Kortum R. Design and Performance of a Low-Cost, Handheld Reader for Diagnosing Anemia in Blantyre, Malawi. Health Innov Point Care Conf. 2014;2014:267–70.

32. Choukem S-P, Sih C, Ntumsi AT, Dimala CA, Mboue-Djieka Y, Ngouadjeu EDT, et al. Evaluation of the accuracy of two point-of-care haemoglobin meters used in sub-Saharan African population: a cross-sectional study. BMC cardiovascular disorders. 2020;20(1):111.

33. Cusick SE, Opoka RO, Ssemata AS, Georgieff MK, John CC. Comparison of iron status 28 d after provision of antimalarial treatment with iron therapy compared with antimalarial treatment alone in Ugandan children with severe malaria. American Journal of Clinical Nutrition. 2016;103(3):919–25.

34. Cusick SE, Opoka RO, Abrams SA, John CC, Georgieff MK, Mupere E. Delaying Iron therapy until 28 days after antimalarial treatment is associated with greater Iron incorporation and equivalent hematologic recovery after 56 days in children: A randomized controlled trial. Journal of Nutrition. 2016;146(9):1769–74.

35. Degarege A, Animut A, Medhin G, Legesse M, Erko B. The association between multiple intestinal helminth infections and blood group, anaemia and nutritional status in human populations from Dore Bafeno, southern Ethiopia. Journal of Helminthology. 2014;88(2):152–9.

36. de Wit M, Funk AL, Moussally K, Nkuba DA, Siddiqui R, Bil K, et al. In vivo efficacy of artesunate-amodiaquine and artemether-lumefantrine for the treatment of uncomplicated falciparum malaria: an open-randomized, non-inferiority clinical trial in South Kivu, Democratic Republic of Congo. Malar J. 2016;15(1):455.

37. Dhabangi A, Ainomugisha B, Cserti-Gazdewich CM, Ddungu H, Kyeyune D, Musisi E, et al. Tissue oxygenation by transfusion in severe anemia with lactic acidosis (total): A prospective, randomized, non-inferiority trial of blood storage duration. Blood. 2015;126(23):769.

38. Gujo AB, Kare AP. Prevalence of Intestinal Parasite Infection and its Association with Anemia among Children Aged 6 to 59 Months in Sidama National Regional State, Southern Ethiopia. Clinical medicine insights Pediatrics. 2021;15:11795565211029259.

39. Heckman J, Samie A, Bessong P, Ntsieni M, Hamandi H, Kohler M, et al. Anaemia among clinically well under-fives attending a community health centre in Venda, Limpopo Province. South African medical journal = Suid-Afrikaanse tydskrif vir geneeskunde. 2010;100(7):445–8.

40. Kamugisha ML, Msangeni H, Beale E, Malecele EK, Akida J, Ishengoma DR, et al. Paracheck PfR compared with microscopy for diagnosis of Plasmodium falciparum malaria among children in Tanga City, north-eastern Tanzania. Tanzania Journal of Health Research. 2008;10(1):14–9.

41. Kebede D, Getaneh F, Endalamaw K, Belay T, Fenta A. Prevalence of anemia and its associated factors among under-five age children in Shanan gibe hospital, Southwest Ethiopia. BMC pediatrics. 2021;21(1):542.

42. Keitel K, Kagoro F, Samaka J, Masimba J, Said Z, Temba H, et al. A novel electronic algorithm using host biomarker point-of-care tests for the management of febrile illnesses in Tanzanian children (e-POCT): A randomized, controlled non-inferiority trial. PLoS Med. 2017;14(10):e1002411.

43. Maiga H, Barger B, Sagara I, Guindo A, Traore OB, Tekete M, et al. Impact of three-year intermittent preventive treatment using artemisinin-based combination therapies on malaria morbidity in malian schoolchildren. Tropical Medicine and Infectious Disease. 2020;5(3):148.

44. Maitland K, Olupot-Olupot P, Kiguli S, Chagaluka G, Alaroker F, Opoka R, et al. Transfusion Volume for Children with Severe Anemia in Africa. The New England journal of medicine. 2019;381:420–31.

45. George E, Uyoga S, M’Baya B, Byabazair D, Kiguli S, Olupot-Olupot P, et al. Whole blood versus red cell concentrates for children with severe anaemia: a secondary analysis of the Transfusion and Treatment of African Children (TRACT) trial. The Lancet Global Health. 2022;10:e360–e8.

46. Maitland K, George EC, Evans JA, Kiguli S, Olupot-Olupot P, Akech SO, et al. Exploring mechanisms of excess mortality with early fluid resuscitation: insights from the FEAST trial. BMC Med. 2013;11:68.

47. McGann PT, Tyburski EA, de Oliveira V, Santos B, Ware RE, Lam WA. An accurate and inexpensive color-based assay for detecting severe anemia in a limited-resource setting. Am J Hematol. 2015;90(12):1122–7.

48. Mtove G, Amos B, von Seidlein L, Hendriksen I, Mwambuli A, Kimera J, et al. Invasive salmonellosis among children admitted to a rural Tanzanian hospital and a comparison with previous studies. PloS one. 2010;5(2):e9244.

49. Nass SA, Hossain I, Sanyang C, Baldeh B, Pereira DIA. Hemoglobin point-of-care testing in rural Gambia: Comparing accuracy of HemoCue and Aptus with an automated hematology analyzer. PLoS One. 2020;15(10):e0239931.

50. Nkrumah B, Nguah SB, Sarpong N, Dekker D, Idriss A, May J, et al. Hemoglobin estimation by the HemoCue® portable hemoglobin photometer in a resource poor setting. BMC Clin Pathol. 2011;11:5.

51. Ntonifor HN, Chewa JS, Oumar M, Mbouobda HD. Intestinal helminths as predictors of some malaria clinical outcomes and il-1beta levels in outpatients attending two public hospitals in bamenda, north west cameroon. PLoS Neglected Tropical Diseases. 2021;15(3):e0009174.

52. Ocan A, Oyet C, Webbo F, Mwambi B, Taremwa IM. Prevalence, morphological characterization, and associated factors of anemia among children below 5 years of age attending St. Mary’s Hospital Lacor, Gulu District, Northern Uganda. Journal of blood medicine. 2018;9:195–201.

53. Olatunya OS, Oke OJ, Kuti BP, Ajayi IA, Olajuyin O, Omotosho-Olagoke O, et al. Factors influencing the academic performance of children with sickle cell anaemia in Ekiti, South West Nigeria. Journal of Tropical Pediatrics. 2018;64(1):67–74.

54. Olatunya OS, Olu-Taiwo A, Ogundare EO, Oluwayemi IO, Olaleye AO, Fadare JO, et al. Evaluation of a Portable Haemoglobin Metre Performance in Children with Sickle Cell Disease and Implications for Healthcare in Resource-poor Settings. Journal of tropical pediatrics. 2016;62(4):316–23.

55. Olupot-Olupot P, Prevatt N, Engoru C, Nteziyaremye J, Amorut D, Chebet M, et al. Evaluation of the diagnostic accuracy and cost of different methods for the assessment of severe anaemia in hospitalised children in Eastern Uganda. Wellcome open research. 2018;3:130.

56. Onyangore FO, Were GM, Mwamburi LA. Prevalence of iron deficiency anaemia and dietary iron intake among infants aged six to nine months in Keiyo South Sub County, Kenya. African Journal of Food, Agriculture, Nutrition and Development. 2016;16(2):10884–97.

57. Parbey PA, Tarkang E, Manu E, Amu H, Ayanore MA, Aku FY, et al. Risk Factors of Anaemia among Children under Five Years in the Hohoe Municipality, Ghana: A Case Control Study. Anemia. 2019;2019:2139717.

58. Ramaswamy G, Vohra K, Yadav K, Kaur R, Rai T, Jaiswal A, et al. Point-of-Care Testing Using Invasive and Non-Invasive Hemoglobinometers: Reliable and Valid Method for Estimation of Hemoglobin among Children 6-59 Months. J Trop Pediatr. 2021;67(1).

59. Silva DLF, Hofelmann DA, Taconeli CA, Lang RMF, Dallazen C, Tietzmann DC, et al. Individual and contextual predictors of children’s hemoglobin levels from Southern Brazilian municipalities in social vulnerability. Cadernos de saude publica. 2021;36(12):e00166619.

60. Smart LR, Ambrose EE, Raphael KC, Hokororo A, Kamugisha E, Tyburski EA, et al. Simultaneous point-of-care detection of anemia and sickle cell disease in Tanzania: the RAPID study. Ann Hematol. 2018;97(2):239–46.

61. Teshome EM, Andang’o PEA, Osoti V, Terwel SR, Otieno W, Demir AY, et al. Daily home fortification with iron as ferrous fumarate versus NaFeEDTA: a randomised, placebo-controlled, non-inferiority trial in Kenyan children. BMC Med. 2017;15(1):89.

62. Ughasoro MD, Madu AJ, Kela-Eke IC, Akubuilo U. Parental Perception of Childhood Anaemia and Efficiency of Instrument Assisted Pallor Detection among Mothers in Southeast Nigeria: A Field Validation Study. International Journal of Pediatrics. 2019;2019.

63. Ughasoro MD, Madu AJ, Kela-Eke IC. Evaluation of the Performance of Haemoglobin Colour Scale and Comparison with HemoCue Haemoglobin Assay in Diagnosing Childhood Anaemia: A Field Validation Study. Int J Pediatr. 2019;2019:3863070.

64. ClinicalTrials.gov [Internet]. Therapeutic Efficacy Study of AL and DP in Western Kenya. 2022; Available from: https://clinicaltrialsgov/show/NCT05060198. nct Identifier: NCT05060198. Accessed 20 June 2022.

65. ClinicalTrials.gov [Internet]. A Trial of 2 ’Point of Care’ Diagnostic Methods to Improve Detection and Treatment of Anaemia in African Children (EARS). 2022; Available from: https://clinicaltrials.gov/ct2/show/NCT00439595. nct Identifier: NCT00439595. Accessed 20 June 2022.

66. Jaggernath M, Naicker R, Madurai S, Brockman MA, Ndung’u T, Gelderblom HC. Diagnostic Accuracy of the HemoCue Hb 301, STAT-Site MHgb and URIT-12 Point-of-Care Hemoglobin Meters in a Central Laboratory and a Community Based Clinic in Durban, South Africa. PloS one. 2016;11(4):e0152184.

67. Raithatha SJ, Ranapurwala MF, Lahori S, Bopche CN, Phatak AG. Validation of hemochroma PLUS: A point of care testing device for haemoglobin estimation. Journal of Clinical and Diagnostic Research. 2019;13(5):LC07-LC9.

68. Singh A, Dubey A, Sonker A, Chaudhary R. Evaluation of various methods of point-of-care testing of haemoglobin concentration in blood donors. Blood Transfus. 2015;13(2):233–9.

69. Ahmad N, Awaluddin SM, Samad R, Noraida R, Kasim M, Yusof M, et al. Validity of Point-of-Care Testing Mission Plus in Detecting Anemia. International Journal of BioMedicine. 2015;5:91–4.

70. Mannino RG, Myers DR, Tyburski EA, Caruso C, Boudreaux J, Leong T, et al. Smartphone app for non-invasive detection of anemia using only patient-sourced photos. Nat Commun. 2018;9(1):4924.

71. Chimbatata CS, Chisale MR, Kayira AB, Sinyiza FW, Mbakaya BC, Kaseka PU, et al. Paediatric sickle cell disease at a tertiary hospital in Malawi: a retrospective cross-sectional study. BMJ Paediatr Open. 2021;5(1):e001097.

72. World Health Organisation. Pocket book of hospital care for children: guidelines for the management of common childhood illnesses. Second ed. Geneva: World Health Organization; 2013. p308.

73. Shamah Levy T, Méndez-Gómez-Humarán I, Morales Ruán MD, Martinez Tapia B, Villalpando Hernández S, Hernández Ávila M. Validation of Masimo Pronto 7 and HemoCue 201 for hemoglobin determination in children from 1 to 5 years of age. PLoS One. 2017;12(2):e0170990.

74. Gwetu TP, Chhagan MK. Evaluation of the diagnostic accuracy of the HemoCue device for detecting anaemia in healthy school-aged children in KwaZulu-Natal, South Africa. S Afr Med J. 2015;105(7):596–9.

75. Gwetu TP, Chhagan M, Craib M, Kauchali S. Hemocue Validation for the Diagnosis of Anaemia in Children: A Semi-Systematic Review. Pediatrics & Therapeutics. 2014;4:1–4.

76. Hinnouho G-M, Barffour MA, Wessells KR, Brown KH, Kounnavong S, Chanhthavong B, et al. Comparison of haemoglobin assessments by HemoCue and two automated haematology analysers in young Laotian children. Journal of Clinical Pathology. 2018;71(6):532-.

77. Lindblade KA, Mwololo K, Van Eijk AM, Peterson E, Odhiambo F, Williamson J, et al. Evaluation of the WHO Haemoglobin Colour Scale for diagnosis of anaemia in children and pregnant women as used by primary health care nurses and community health workers in western Kenya. Tropical Medicine & International Health. 2006;11(11):1679–87.

78. Karakochuk CD, Janmohamed A, Whitfield KC, Barr SI, Vercauteren SM, Kroeun H, et al. Evaluation of two methods to measure hemoglobin concentration among women with genetic hemoglobin disorders in Cambodia: a method-comparison study. Clin Chim Acta. 2015;441:148–55.

79. Hiscock R, Kumar D, Simmons SW. Systematic review and meta-analysis of method comparison studies of Masimo pulse co-oximeters (Radical-7™ or Pronto-7™) and HemoCue® absorption spectrometers (B-Hemoglobin or 201+) with laboratory haemoglobin estimation. Anaesth Intensive Care. 2015;43(3):341–50.

80. Shabaninejad H, Ghadimi N, Sayehmiri K, Hosseinifard H, Azarfarin R, Gorji HA. Comparison of invasive and noninvasive blood hemoglobin measurement in the operating room: a systematic review and meta-analysis. J Anesth. 2019;33(3):441–53.

81. An R, Huang Y, Man Y, Valentine RW, Kucukal E, Goreke U, et al. Emerging point-of-care technologies for anemia detection. Lab Chip. 2021;21(10):1843–65.

82. Neogi SB, Jyoti S, Shivam P, Nausheen Z, Maitreyee B, Rakhee K, et al. Diagnostic accuracy of point-of-care devices for detection of anemia in community settings in India. BMC Health Services Research. 2020;20(468).

83. Whitehead RD, Zhang M, Sternberg MR, Schleicher RL, Drammeh B, Mapango C, et al. Effects of preanalytical factors on hemoglobin measurement: A comparison of two HemoCue® point-of-care analyzers. Clinical Biochemistry. 2017;50(9):513–20.

84. Jain A, Chowdhury N. Comparison of the accuracy of capillary hemoglobin estimation and venous hemoglobin estimation by two models of HemoCue against automated cell counter hemoglobin measurement. Asian journal of transfusion science. 2020;14(1):49–53.

85. Medina Lara A, Mundy C, Kandulu J, Chisuwo L, Bates I. Evaluation and costs of different haemoglobin methods for use in district hospitals in Malawi. J Clin Pathol. 2005;58(1):56–60.

86. Lingervelder D, Koffijberg H, Kusters R, MJ IJ. Health Economic Evidence of Point-of-Care Testing: A Systematic Review. Pharmacoecon Open. 2021;5(2):157–73.

87. Wemyss TA, Nixon-Hill M, Outlaw F, Karsa A, Meek J, Enweronu-Laryea C, et al. Feasibility of smartphone colorimetry of the face as an anaemia screening tool for infants and young children in Ghana. PLoS One. 2023;18(3):e0281736.

