## Supplementary Material for "Use of Point-of-care Haemoglobin Tests to Diagnose Childhood Anaemia in Low-and Middle-Income Countries: A Systematic Review"

**Abbreviations**

POC(Hb)T: Point-of-care haemoglobin test

**Supplementary Table S1: Search Strategy**

Concepts 1-4 (combined with AND operator):

1. Anaemia
2. POC Hb tests
3. LMICs
4. Clinical Setting

Truncations and subject headings were adapted to each database.

LMIC filters were adapted from Cochrane Effective Practice and Organisation of Care filters for LMICs and updated according to World Banks Classification 2022.[1, 2] 28 countries were removed, and two countries added (Kiribati and Tuvalu) due to change in class. Venezuela was unclassified in 2022 and so remained included.

Entire platforms were searched on 10th June 2022 and English language filters applied.

| Database | Result |
|---|---|
| **MEDLINE (Ovid)**<br><br>1 exp Anemia/ (171178)<br>2 an?emi*.ti,ab. (160879)<br>3 1 or 2 (244796)<br>4 point-of-care systems/ or point-of-care testing/ or rapid on-site evaluation/ (18545)<br>5 Hemoglobinometry/ (6564)<br>6 Colorimetry/ (23666)<br>7 point-of-care test*.ti,ab. (6235)<br>8 point-of-care system*.ti,ab. (267)<br>9 point-of-care diagnostic*.ti,ab. (1968)<br>10 point-of-care measurement*.ti,ab. (124)<br>11 (rapid on-site adj2 (evaluation* or test*)).ti,ab. (578)<br>12 h?emoglobin determination.ti,ab. (239)<br>13 point-of-care Hb.ti,ab. (7)<br>14 (point-of-care adj3 h?emoglobin*).ti,ab. (158)<br>15 POC.ti,ab. (6491)<br>16 POCT.ti,ab. (2342)<br>17 h?emoglobinometry.ti,ab. (188)<br>18 HemoCue.ti,ab. (554)<br>19 TrueHb.ti,ab. (7)<br>20 Sahli's method.ti,ab. (13)<br>21 DiaSpect.ti,ab. (2)<br>22 Radical-7.ti,ab. (140)<br>23 pronto-7.ti,ab. (21)<br>24 I-STAT.ti,ab. (301)<br>25 h?emoglobin Colo?r Scale.ti,ab. (52)<br>26 HCS.ti,ab. (10075) | **104** |

| |
|---|
| 27 colo?rimetry.ti,ab. (3122)<br>28 Gem 4000.ti,ab. (14)<br>29 Hemo Control.ti,ab. (2)<br>30 STAT-Site Mhgb.ti,ab. (3)<br>31 URIT-12.ti,ab. (6)<br>32 Aptus.ti,ab. (64)<br>33 AnemoCheck LRS.ti,ab. (1)<br>34 AnemoCheck Low Resource Setting*.ti,ab. (0)<br>35 Mission Hb.ti,ab. (2)<br>36 Mission R plus.ti,ab. (1)<br>37 or/4-36 (71248)<br>38 (afghanistan or albania or algeria or american samoa or angola or argentina or armenia or armenian or azerbaijan or bangladesh or republic of belarus or belarus or byelarus or belorussia or byelorussian or belize or british honduras or benin or dahomey or bhutan or bolivia or "bosnia and herzegovina" or bosnia or herzegovina or botswana or bechuanaland or brazil or brasil or bulgaria or burkina faso or burkina fasso or upper volta or burundi or urundi or cabo verde or cape verde or cambodia or kampuchea or khmer republic or cameroon or cameron or cameroun or central african republic or ubangi shari or chad or china or colombia or comoros or comoro islands or iles comores or mayotte or democratic republic of the congo or democratic republic congo or congo or zaire or costa rica or "cote d'ivoire" or "cote d' ivoire" or cote divoire or cote d ivoire or ivory coast or cuba or djibouti or french somaliland or dominica or dominican republic or ecuador or egypt or united arab republic or el salvador or equatorial guinea or spanish guinea or eritrea or eswatini or swaziland or ethiopia or fiji or gabon or gabonese republic or gambia or "georgia (republic)" or georgian or ghana or gold coast or grenada or guam or guatemala or guinea or guinea bissau or guyana or british guiana or haiti or hispaniola or honduras or india or indonesia or timor or iran or iraq or isle of man or jamaica or jordan or kazakhstan or kazakh or kenya or Kiribati or "democratic people's republic of korea" or republic of korea or north korea or south korea or korea or kosovo or kyrgyzstan or kirghizia or kirgizstan or kyrgyz republic or kirghiz or laos or lao pdr or "lao people's democratic republic" or lebanon or lebanese republic or lesotho or basutoland or liberia or libya or libyan arab jamahiriya or macau or macao or republic of north macedonia or macedonia or madagascar or malagasy republic or malawi or nyasaland or malaysia or malay federation or malaya federation or maldives or indian ocean islands or indian ocean or mali or micronesia or federated states of micronesia or kiribati or marshall islands or nauru or northern mariana islands or palau or tuvalu or mauritania or mauritius or mexico or moldova or moldovian or mongolia or montenegro or morocco or ifni or mozambique or portuguese east africa or myanmar or burma or namibia or Nepal or nicaragua or niger or nigeria or pakistan or panama or papua new guinea or new guinea or paraguay or peru or philippines or philipines or phillipines or phillippines or romania or russia or russian federation or ussr or soviet union or union of soviet socialist republics or rwanda or ruanda or samoa or pacific islands or polynesia or samoan islands or navigator island or navigator islands or "sao tome and principe" or senegal or serbia or sierra leone or melanesia or solomon island or solomon islands or norfolk island or norfolk islands or somalia or south africa or south sudan or sri lanka or Ceylon or saint lucia or "st. lucia" or "saint vincent and the grenadines" or saint vincent or "st. vincent" or grenadines or sudan or suriname or surinam or dutch guiana or netherlands guiana or syria or syrian arab republic or tajikistan or tadjikistan or tadzhikistan or tadzhik or tanzania or tanganyika or thailand or siam or timor leste or east timor or togo or togolese republic or tonga or tunisia or turkey or turkmenistan or turkmen or Tuvalu or uganda or ukraine or uzbekistan or uzbek or vanuatu or new hebrides or venezuela or vietnam or viet nam or middle east or west bank |

| |
|---|
| or gaza or palestine or yemen or zambia or zimbabwe or northern rhodesia or global south or africa south of the sahara or sub-saharan africa or subsaharan africa or africa, central or central africa or africa, northern or north africa or northern africa or magreb or maghrib or sahara or africa, southern or southern africa or africa, eastern or east africa or eastern africa or africa, western or west africa or western africa or west indies or indian ocean islands or caribbean or central america or latin america or "south and central america" or south america or asia, central or central asia or asia, northern or north asia or northern asia or asia, southeastern or southeastern asia or south eastern asia or southeast asia or south east asia or asia, western or western asia or europe, eastern or east europe or eastern europe or developing country or developing countries or developing nation? or developing population? or developing world or less developed countr* or less developed nation? or less developed population? or less developed world or lesser developed countr* or lesser developed nation? or lesser developed population? or lesser developed world or under developed countr* or under developed nation? or under developed population? or under developed world or underdeveloped countr* or underdeveloped nation? or underdeveloped population? or underdeveloped world or middle income countr* or middle income nation? or middle income population? or low income countr* or low income nation? or low income population? or lower income countr* or lower income nation? or lower income population? or underserved countr* or underserved nation? or underserved population? or underserved world or under served countr* or under served nation? or under served population? or under served world or deprived countr* or deprived nation? or deprived population? or deprived world or poor countr* or poor nation? or poor population? or poor world or poorer countr* or poorer nation? or poorer population? or poorer world or developing econom* or less developed econom* or lesser developed econom* or under developed econom* or underdeveloped econom* or middle income econom* or low income econom* or lower income econom* or low gdp or low gnp or low gross domestic or low gross national or lower gdp or lower gnp or lower gross domestic or lower gross national or lmic or lmics or third world or lami countr* or transitional countr* or emerging economies or emerging nation?).ti,ab. (1453878)<br>39 Developing Countries/ (79392)<br>40 asia, central/ or asia, northern/ or asia, southeastern/ or asia, western/ or bangladesh/ or bhutan/ or india/ or middle east/ or afghanistan/ or iran/ or iraq/ or jordan/ or lebanon/ or syria/ or turkey/ or yemen/ or nepal/ or pakistan/ or sri lanka/ (273134)<br>41 europe, eastern/ or albania/ or "bosnia and herzegovina"/ or bulgaria/ or kosovo/ or "republic of north macedonia"/ or moldova/ or montenegro/ or "republic of belarus"/ or romania/ or russia/ or serbia/ or ukraine/ (87632)<br>42 samoa/ or american samoa/ (585)<br>43 africa, northern/ or "africa south of the sahara"/ or africa, central/ or africa, eastern/ or africa, southern/ or angola/ or botswana/ or eswatini/ or lesotho/ or malawi/ or mozambique/ or namibia/ or south africa/ or zambia/ or zimbabwe/ or africa, western/ (97219)<br>44 algeria/ or egypt/ or libya/ or morocco/ or tunisia/ (36191)<br>45 central america/ or latin america/ or south america/ or argentina/ or bolivia/ or brazil/ or colombia/ or ecuador/ or guyana/ or paraguay/ or peru/ or suriname/ or venezuela/ (181894)<br>46 armenia/ or azerbaijan/ or "georgia (republic)"/ (4639)<br>47 belize/ or costa rica/ or el salvador/ or guatemala/ or honduras/ or nicaragua/ or panama/ (13367) |

| | |
|---|---|
| 48 benin/ or burkina faso/ or cabo verde/ or cote d'ivoire/ or gambia/ or ghana/ or guinea/ or guinea-bissau/ or liberia/ or mali/ or mauritania/ or niger/ or nigeria/ or senegal/ or sierra leone/ or togo/ (68991)<br>49 burundi/ or djibouti/ or eritrea/ or ethiopia/ or kenya/ or rwanda/ or somalia/ or south sudan/ or sudan/ or tanzania/ or uganda/ (71365)<br>50 cambodia/ or indonesia/ or laos/ or malaysia/ or myanmar/ or philippines/ or thailand/ or timor-leste/ or vietnam/ (87794)<br>51 cameroon/ or central african republic/ or chad/ or congo/ or "democratic republic of the congo"/ or equatorial guinea/ or gabon/ or "sao tome and principe"/ (16144)<br>52 comoros/ or madagascar/ or mauritius/ (4708)<br>53 cuba/ or dominica/ or dominican republic/ or grenada/ or haiti/ or jamaica/ or saint lucia/ or "saint vincent and the grenadines"/ (14104)<br>54 indian ocean islands/ or fiji/ or papua new guinea/ or vanuatu/ or micronesia/ or west indies/ (10768)<br>55 macau/ or guam/ or palau/ (1264)<br>56 kazakhstan/ or kyrgyzstan/ or tajikistan/ or turkmenistan/ or uzbekistan/ (7296)<br>57 china/ or korea/ or "democratic people's republic of korea"/ or "republic of korea"/ or mongolia/ (283532)<br>58 Mexico/ (42673)<br>59 Caribbean Region/ (5242)<br>60 or/38-59 (1933182)<br>61 exp Hospitals/ (304281)<br>62 Community Health Centers/ (7444)<br>63 hospital*.ti,ab. (1487453)<br>64 (Community adj3 Health).ti,ab. (53776)<br>65 (clinic or clinics).ti,ab. (363067)<br>66 public health facilit*. ti,ab. (1854)<br>67 or/61-66 (1922465)<br>68 3 and 37 and 60 and 67 (111)<br>69 limit 68 to English language (104) | |
| **EMBASE (Ovid)**<br><br>1 exp *anemia/ (149720)<br>2 an?emi*.ti,ab. (233655)<br>3 1 or 2 (297890)<br>4 "point of care system"/ (3268)<br>5 "point of care testing"/ (17898)<br>6 hemoglobin determination/ (23863)<br>7 colorimetry/ (38374)<br>8 h?emoglobinometry.ti,ab. (88)<br>9 h?emoglobin determination.ti,ab. (251)<br>10 point-of-care test*.ti,ab. (8472)<br>11 point-of-care system*.ti,ab. (374)<br>12 point-of-care diagnostic*.ti,ab. (2138)<br>13 point-of-care measurement*.ti,ab. (200)<br>14 (rapid on-site adj2 (evaluation* or test*)).ti,ab. (1203)<br>15 point-of-care Hb.ti,ab. (16)<br>16 (point-of-care adj3 h?emoglobin*).ti,ab. (262)<br>17 POC.ti,ab. (8580)<br>18 h?emoglobinometry.ti,ab. (88) | **417** |

| |
|---|
| 19 HemoCue.ti,ab. (953)<br>20 TrueHb.ti,ab. (6)<br>21 Sahli's method.ti,ab. (40)<br>22 DiaSpect.ti,ab. (11)<br>23 Radical-7.ti,ab. (299)<br>24 pronto-7.ti,ab. (58)<br>25 I-STAT.ti,ab. (554)<br>26 h?emoglobin Colo?r Scale.ti,ab. (54)<br>27 HCS.ti,ab. (14289)<br>28 colo?rimetry.ti,ab. (4085)<br>29 Gem 4000.ti,ab. (48)<br>30 Hemo Control.ti,ab. (10)<br>31 STAT-Site Mhgb.ti,ab. (4)<br>32 URIT-12.ti,ab. (7)<br>33 Aptus.ti,ab. (93)<br>34 AnemoCheck LRS.ti,ab. (3)<br>35 AnemoCheck Low Resource Setting*.ti,ab. (0)<br>36 Mission Hb.ti,ab. (3)<br>37 Mission R plus.ti,ab. (0)<br>38 4 or 5 or 6 or 7 or 8 or 9 or 10 or 11 or 12 or 13 or 14 or 15 or 16 or 17 or 18 or 19 or 20 or 21 or 22 or 23 or 24 or 25 or 26 or 27 or 28 or 29 or 30 or 31 or 32 or 33 or 34 or 35 or 36 or 37 (111205)<br>39 (afghanistan or albania or algeria or american samoa or angola or argentina or armenia or armenian or azerbaijan or bangladesh or republic of belarus or belarus or byelarus or belorussia or byelorussian or belize or british honduras or benin or dahomey or bhutan or bolivia or "bosnia and herzegovina" or bosnia or herzegovina or botswana or bechuanaland or brazil or brasil or bulgaria or burkina faso or burkina fasso or upper volta or burundi or urundi or cabo verde or cape verde or cambodia or kampuchea or khmer republic or cameroon or cameron or cameroun or central african republic or ubangi shari or chad or china or colombia or comoros or comoro islands or iles comores or mayotte or democratic republic of the congo or democratic republic congo or congo or zaire or costa rica or "cote d’ivoire" or "cote d’ ivoire" or cote divoire or cote d ivoire or ivory coast or cuba or djibouti or french somaliland or dominica or dominican republic or ecuador or egypt or united arab republic or el salvador or equatorial guinea or spanish guinea or eritrea or eswatini or swaziland or ethiopia or fiji or gabon or gabonese republic or gambia or "georgia (republic)" or georgian or ghana or gold coast or grenada or guam or guatemala or guinea or guinea bissau or guyana or british guiana or haiti or hispaniola or honduras or india or indonesia or timor or iran or iraq or isle of man or jamaica or jordan or kazakhstan or kazakh or kenya or Kiribati or "democratic people’s republic of korea" or republic of korea or north korea or south korea or korea or kosovo or kyrgyzstan or kirghizia or kirgizstan or kyrgyz republic or kirghiz or laos or lao pdr or "lao people's democratic republic" or lebanon or lebanese republic or lesotho or basutoland or liberia or libya or libyan arab jamahiriya or macau or macao or republic of north macedonia or macedonia or madagascar or malagasy republic or malawi or nyasaland or malaysia or malay federation or malaya federation or maldives or indian ocean islands or indian ocean or mali or micronesia or federated states of micronesia or kiribati or marshall islands or nauru or northern mariana islands or palau or tuvalu or mauritania or mauritius or mexico or moldova or moldovian or mongolia or montenegro or morocco or ifni or mozambique or portuguese east africa or myanmar or burma or namibia or Nepal or nicaragua or niger or nigeria or pakistan or panama or papua new guinea or new guinea or paraguay or peru or philippines or philipines or phillipines or phillippines or romania or russia or russian |

| |
|---|
| federation or ussr or soviet union or union of soviet socialist republics or rwanda or ruanda or samoa or pacific islands or polynesia or samoan islands or navigator island or navigator islands or "sao tome and principe" or senegal or serbia or sierra leone or melanesia or solomon island or solomon islands or norfolk island or norfolk islands or somalia or south africa or south sudan or sri lanka or Ceylon or saint lucia or "st. lucia" or "saint vincent and the grenadines" or saint vincent or "st. vincent" or grenadines or sudan or suriname or surinam or dutch guiana or netherlands guiana or syria or syrian arab republic or tajikistan or tadjikistan or tadzhikistan or tadzhik or tanzania or tanganyika or thailand or siam or timor leste or east timor or togo or togolese republic or tonga or tunisia or turkey or turkmenistan or turkmen or Tuvalu or uganda or ukraine or uzbekistan or uzbek or vanuatu or new hebrides or venezuela or vietnam or viet nam or middle east or west bank or gaza or palestine or yemen or zambia or zimbabwe or northern rhodesia or global south or africa south of the sahara or sub-saharan africa or subsaharan africa or africa, central or central africa or africa, northern or north africa or northern africa or magreb or maghrib or sahara or africa, southern or southern africa or africa, eastern or east africa or eastern africa or africa, western or west africa or western africa or west indies or indian ocean islands or caribbean or central america or latin america or "south and central america" or south america or asia, central or central asia or asia, northern or north asia or northern asia or asia, southeastern or southeastern asia or south eastern asia or southeast asia or south east asia or asia, western or western asia or europe, eastern or east europe or eastern europe or developing country or developing countries or developing nation? or developing population? or developing world or less developed countr* or less developed nation? or less developed population? or less developed world or lesser developed countr* or lesser developed nation? or lesser developed population? or lesser developed world or under developed countr* or under developed nation? or under developed population? or under developed world or underdeveloped countr* or underdeveloped nation? or underdeveloped population? or underdeveloped world or middle income countr* or middle income nation? or middle income population? or low income countr* or low income nation? or low income population? or lower income countr* or lower income nation? or lower income population? or underserved countr* or underserved nation? or underserved population? or underserved world or under served countr* or under served nation? or under served population? or under served world or deprived countr* or deprived nation? or deprived population? or deprived world or poor countr* or poor nation? or poor population? or poor world or poorer countr* or poorer nation? or poorer population? or poorer world or developing econom* or less developed econom* or lesser developed econom* or under developed econom* or underdeveloped econom* or middle income econom* or low income econom* or lower income econom* or low gdp or low gnp or low gross domestic or low gross national or lower gdp or lower gnp or lower gross domestic or lower gross national or lmic or lmics or third world or lami countr* or transitional countr* or emerging economies or emerging nation?).ti,ab. (1770299)<br>40 developing country/ (98314)<br>41 low income country/ (10706)<br>42 middle income country/ (15194)<br>43 Afghanistan/ (6566)<br>44 American Samoa/ (285)<br>45 Bangladesh/ (19599)<br>46 Belarus/ (2917)<br>47 Bhutan/ (1029)<br>48 exp *China/ (32457)<br>49 Dominica/ (215) |

| |
|---|
| 50 Fiji/ (1787)<br>51 exp *"Georgia (republic)"/ (98)<br>52 Guam/ (947)<br>53 exp *India/ (25953)<br>54 exp *Korea/ (8309)<br>55 Macao/ (635)<br>56 maldives/ (402)<br>57 exp *"Federated States of Micronesia"/ (39)<br>58 Palau/ (328)<br>59 Mauritius/ (984)<br>60 exp *Mexico/ (5229)<br>61 Mongolia/ (3592)<br>62 Nepal/ (14282)<br>63 exp *Pakistan/ (3888)<br>64 Philippines/ (12530)<br>65 Samoa/ (657)<br>66 Melanesia/ (802)<br>67 south sudan/ (423)<br>68 Sri Lanka/ (9505)<br>69 Tonga/ (410)<br>70 Vanuatu/ (523)<br>71 "africa south of the sahara"/ or angola/ or benin/ or botswana/ or burkina faso/ or burundi/ or cameroon/ or cape verde/ or central africa/ or central african republic/ or chad/ or comoros/ or congo/ or cote d'ivoire/ or democratic republic congo/ or djibouti/ or equatorial guinea/ or eritrea/ or eswatini/ or ethiopia/ or gabon/ or gambia/ or ghana/ or guinea/ or guinea-bissau/ or kenya/ or lesotho/ or liberia/ or madagascar/ or malawi/ or mali/ or mozambique/ or namibia/ or niger/ or nigeria/ or rwanda/ or senegal/ or sierra leone/ or somalia/ or south africa/ or south sudan/ or sudan/ or tanzania/ or togo/ or uganda/ or zambia/ or zimbabwe/ (282372)<br>72 north africa/ or algeria/ or egypt/ or libyan arab jamahiriya/ or mauritania/ or morocco/ or tunisia/ or western sahara/ (49496)<br>73 caribbean islands/ or cuba/ or dominica/ or dominican republic/ or grenada/ or haiti/ or jamaica/ or saint lucia/ or "saint vincent and the grenadines"/ (21123)<br>74 central america/ or belize/ or caribbean/ or costa rica/ or el salvador/ or guatemala/ or honduras/ or nicaragua/ or panama/ (29159)<br>75 south america/ or argentina/ or bolivia/ or brazil/ or colombia/ or ecuador/ or guyana/ or paraguay/ or peru/ or suriname/ or venezuela/ (210044)<br>76 central asia/ or kazakhstan/ or kyrgyzstan/ or tajikistan/ or turkmenistan/ or uzbekistan/ (9018)<br>77 northern asia/ (7)<br>78 southeast asia/ or cambodia/ or indonesia/ or laos/ or malaysia/ or myanmar/ or papua new guinea/ or thailand/ or timor-leste/ or viet nam/ (120232)<br>79 western asia/ or armenia/ or azerbaijan/ or iran/ or iraq/ or jordan/ or lebanon/ or syrian arab republic/ or "turkey (republic)"/ or yemen/ (130544)<br>80 eastern europe/ or albania/ or "bosnia and herzegovina"/ or bulgaria/ or kosovo/ or moldova/ or "montenegro (republic)"/ or "republic of north macedonia"/ or romania/ or russian federation/ or serbia/ or ukraine/ (115766)<br>81 eastern europe/ or albania/ or "bosnia and herzegovina"/ or bulgaria/ or kosovo/ or moldova/ or "montenegro (republic)"/ or "republic of north macedonia"/ or romania/ or russian federation/ or serbia/ or ukraine/ (115766) |

| | |
|---|---|
| 82 39 or 40 or 41 or 42 or 43 or 44 or 45 or 46 or 47 or 48 or 49 or 50 or 51 or 52 or 53 or 54 or 55 or 56 or 57 or 58 or 59 or 60 or 61 or 62 or 63 or 64 or 65 or 66 or 67 or 68 or 69 or 70 or 71 or 72 or 73 or 74 or 75 or 76 or 77 or 78 or 79 or 80 or 81 (2066345)<br>83 exp *hospital/ (307716)<br>84 health center/ (38096)<br>85 hospital*.ti,ab. (2280549)<br>86 (community adj3 Health).ti,ab. (63510)<br>87 (clinic or clinics).ti,ab. (598443)<br>88 public health facilit*.ti,ab. (2173)<br>89 83 or 84 or 85 or 86 or 87 or 88 (2924436)<br>90 3 and 38 and 82 and 89 (434)<br>100 Limit 90 to English language (417) | |
| **Global Health (Ovid)**<br><br>1 anaemia/ (17194)<br>2 an?emi*.ti,ab. (32047)<br>3 or/1-2 (32726)<br>4 rapid methods/ (14604)<br>5 haemoglobin value/ (243)<br>6 colorimetry/ (1878)<br>7 point-of-care test*.ti,ab. (1646)<br>8 point-of-care system*.ti,ab. (22)<br>9 point-of-care diagnostic*.ti,ab. (435)<br>10 point-of-care measurement*.ti,ab. (7)<br>11 (rapid on-site adj2 (evaluation* or test*)).ti,ab. (38)<br>12 h?emoglobin determination.ti,ab. (28)<br>13 point-of-care Hb.ti,ab. (0)<br>14 (point-of-care adj3 h?emoglobin*).ti,ab. (29)<br>15 POC.ti,ab. (1124)<br>16 POCT.ti,ab. (361)<br>17 h?emoglobinometry.ti,ab. (11)<br>18 HemoCue.ti,ab. (228)<br>19 TrueHb.ti,ab. (4)<br>20 Sahli's method.ti,ab. (14)<br>21 DiaSpect.ti,ab. (0)<br>22 Radical-7.ti,ab. (9)<br>23 pronto-7.ti,ab. (2)<br>24 I-STAT.ti,ab. (27)<br>25 h?emoglobin Colo?r Scale.ti,ab. (28)<br>26 HCS.ti,ab. (1006)<br>27 colo?rimetry.ti,ab. (1090)<br>28 Gem 4000.ti,ab. (0)<br>29 Hemo Control.ti,ab. (3)<br>30 STAT-Site Mhgb.ti,ab. (1)<br>31 URIT-12.ti,ab. (2)<br>32 Aptus.ti,ab. (3)<br>33 AnemoCheck LRS.ti,ab. (0)<br>34 AnemoCheck Low Resource Setting*.ti,ab. (0)<br>35 Mission Hb.ti,ab. (1)<br>36 Mission R plus.ti,ab. (0) | **84** |

| |
|---|
| 37 or/4-36 (21304)<br>38 (afghanistan or albania or algeria or american samoa or angola or argentina or armenia or armenian or azerbaijan or bangladesh or republic of belarus or belarus or byelarus or belorussia or byelorussian or belize or british honduras or benin or dahomey or bhutan or bolivia or "bosnia and herzegovina" or bosnia or herzegovina or botswana or bechuanaland or brazil or brasil or bulgaria or burkina faso or burkina fasso or upper volta or burundi or urundi or cabo verde or cape verde or cambodia or kampuchea or khmer republic or cameroon or cameron or cameroun or central african republic or ubangi shari or chad or china or colombia or comoros or comoro islands or iles comores or mayotte or democratic republic of the congo or democratic republic congo or congo or zaire or costa rica or "cote d'ivoire" or "cote d' ivoire" or cote divoire or cote d ivoire or ivory coast or cuba or djibouti or french somaliland or dominica or dominican republic or ecuador or egypt or united arab republic or el salvador or equatorial guinea or spanish guinea or eritrea or eswatini or swaziland or ethiopia or fiji or gabon or gabonese republic or gambia or "georgia (republic)" or georgian or ghana or gold coast or grenada or guam or guatemala or guinea or guinea bissau or guyana or british guiana or haiti or hispaniola or honduras or india or indonesia or timor or iran or iraq or isle of man or jamaica or jordan or kazakhstan or kazakh or kenya or Kiribati or "democratic people's republic of korea" or republic of korea or north korea or south korea or korea or kosovo or kyrgyzstan or kirghizia or kirgizstan or kyrgyz republic or kirghiz or laos or lao pdr or "lao people's democratic republic" or lebanon or lebanese republic or lesotho or basutoland or liberia or libya or libyan arab jamahiriya or macau or macao or republic of north macedonia or macedonia or madagascar or malagasy republic or malawi or nyasaland or malaysia or malay federation or malaya federation or maldives or indian ocean islands or indian ocean or mali or micronesia or federated states of micronesia or kiribati or marshall islands or nauru or northern mariana islands or palau or tuvalu or mauritania or mauritius or mexico or moldova or moldovian or mongolia or montenegro or morocco or ifni or mozambique or portuguese east africa or myanmar or burma or namibia or Nepal or nicaragua or niger or nigeria or pakistan or panama or papua new guinea or new guinea or paraguay or peru or philippines or philipines or phillipines or phillippines or romania or russia or russian federation or ussr or soviet union or union of soviet socialist republics or rwanda or ruanda or samoa or pacific islands or polynesia or samoan islands or navigator island or navigator islands or "sao tome and principe" or senegal or serbia or sierra leone or melanesia or solomon island or solomon islands or norfolk island or norfolk islands or somalia or south africa or south sudan or sri lanka or Ceylon or saint lucia or "st. lucia" or "saint vincent and the grenadines" or saint vincent or "st. vincent" or grenadines or sudan or suriname or surinam or dutch guiana or netherlands guiana or syria or syrian arab republic or tajikistan or tadjikistan or tadzhikistan or tadzhik or tanzania or tanganyika or thailand or siam or timor leste or east timor or togo or togolese republic or tonga or tunisia or turkey or turkmenistan or turkmen or Tuvalu or uganda or ukraine or uzbekistan or uzbek or vanuatu or new hebrides or venezuela or vietnam or viet nam or middle east or west bank or gaza or palestine or yemen or zambia or zimbabwe or northern rhodesia or global south or africa south of the sahara or sub-saharan africa or subsaharan africa or africa, central or central africa or africa, northern or north africa or northern africa or magreb or maghrib or sahara or africa, southern or southern africa or africa, eastern or east africa or eastern africa or africa, western or west africa or western africa or west indies or indian ocean islands or caribbean or central america or latin america or "south and central america" or south america or asia, central or central asia or asia, northern or north asia or northern asia or asia, southeastern or southeastern asia or south eastern asia or southeast asia or south east asia or asia, western or western asia or europe, eastern or east europe or eastern europe or developing country or developing countries or developing nation? or |

| |
|---|
| developing population? or developing world or less developed countr* or less developed nation? or less developed population? or less developed world or lesser developed countr* or lesser developed nation? or lesser developed population? or lesser developed world or under developed countr* or under developed nation? or under developed population? or under developed world or underdeveloped countr* or underdeveloped nation? or underdeveloped population? or underdeveloped world or middle income countr* or middle income nation? or middle income population? or low income countr* or low income nation? or low income population? or lower income countr* or lower income nation? or lower income population? or underserved countr* or underserved nation? or underserved population? or underserved world or under served countr* or under served nation? or under served population? or under served world or deprived countr* or deprived nation? or deprived population? or deprived world or poor countr* or poor nation? or poor population? or poor world or poorer countr* or poorer nation? or poorer population? or poorer world or developing econom* or less developed econom* or lesser developed econom* or under developed econom* or underdeveloped econom* or middle income econom* or low income econom* or lower income econom* or low gdp or low gnp or low gross domestic or low gross national or lower gdp or lower gnp or lower gross domestic or lower gross national or lmic or lmics or third world or lami countr* or transitional countr* or emerging economies or emerging nation?).ti,ab. (805116)<br>39 Developing Countries/ (987312)<br>40 central america/ or belize/ or costa rica/ or el salvador/ or guatemala/ or honduras/ or nicaragua/ or panama/ or latin america/ (183376)<br>41 south america/ or argentina/ or bolivia/ or brazil/ or colombia/ or ecuador/ or guyana/ or paraguay/ or peru/ or suriname/ or venezuela/ (148613)<br>42 central asia/ or afghanistan/ or kazakhstan/ or kyrgyzstan/ or mongolia/ or tajikistan/ or turkmenistan/ or uzbekistan/ (12491)<br>43 south east asia/ or east timor/ or indonesia/ or malaysia/ or myanmar/ or philippines/ or thailand/ (105267)<br>44 west asia/ or armenia/ or azerbaijan/ or iran/ or iraq/ or jordan/ or lebanon/ or syria/ or turkey/ or yemen/ (145942)<br>45 belarus/ or ukraine/ or russia/ (17893)<br>46 north africa/ or egypt/ or libya/ (33980)<br>47 west africa/ or "africa south of sahara"/ or benin/ or burkina faso/ or cape verde/ or cote d'ivoire/ or gambia/ or ghana/ or guinea/ or guinea-bissau/ or liberia/ or mali/ or mauritania/ or niger/ or nigeria/ or senegal/ or sierra leone/ or togo/ or western sahara/ (203631)<br>48 central africa/ or burundi/ or cameroon/ or central african republic/ or chad/ or congo/ or congo democratic republic/ or equatorial guinea/ or gabon/ or "sao tome and principe"/ (17395)<br>49 east africa/ or djibouti/ or eritrea/ or ethiopia/ or kenya/ or madagascar/ or malawi/ or rwanda/ or somalia/ or south sudan/ or sudan/ or tanzania/ or uganda/ (72706)<br>50 southern africa/ or angola/ or botswana/ or comoros/ or lesotho/ or mozambique/ or namibia/ or south africa/ or swaziland/ or zambia/ or zimbabwe/ (47768)<br>51 caribbean community/ or dominica/ or grenada/ or guyana/ or haiti/ or jamaica/ or saint lucia/ or "saint vincent and the grenadines"/ or caribbean/ (18606)<br>52 bulgaria/ (5083)<br>53 Albania/ (1092)<br>54 Algeria/ (3626)<br>55 American Samoa/ (180)<br>56 Bangladesh/ (13955) |

| | |
|---|---|
| 57 Bhutan/ (650)<br>58 bosnia-hercegovina/ (1702)<br>59 Cambodia/ (3337)<br>60 china/ (214767)<br>61 Cuba/ (4657)<br>62 Fiji/ (785)<br>63 "republic of georgia"/ (839)<br>64 Guam/ (264)<br>65 India/ (124164)<br>66 Korea Republic/ or Korea Democratic People's Republic/ (30673)<br>67 Kosovo/ (459)<br>68 Laos/ (1959)<br>69 Macao/ (252)<br>70 "Republic of North Macedonia"/ (131)<br>71 Micronesia/ or "Federated States of Micronesia"/ (937)<br>72 Palau/ (100)<br>73 indian ocean islands/ or maldives/ or mauritius/ (4700)<br>74 Mexico/ (23307)<br>75 Moldova/ (583)<br>76 Montenegro/ (328)<br>77 Morocco/ (6581)<br>78 Nepal/ (8406)<br>79 Pakistan/ (19143)<br>80 Papua New Guinea/ (3481)<br>81 Romania/ (8781)<br>82 Samoa/ (387)<br>83 Serbia/ (3951)<br>84 Sri Lanka/ (5590)<br>85 Melanesia/ (5477)<br>86 Tunisia/ (6076)<br>87 Vanuatu/ (450)<br>88 Vietnam/ (9269)<br>89 or/38-88 (1205134)<br>90 exp hospitals/ (67963)<br>91 community health services/ (4763)<br>92 health centres/ (15776)<br>93 health clinics/ (2307)<br>94 hospital*.ti,ab. (315396)<br>95 (Community adj3 Health).ti,ab. (20699)<br>96 (clinic or clinics).ti,ab. (73365)<br>97 public health facilit*.ti,ab. (1380)<br>98 or/90-97 (404025)<br>99 3 and 37 and 89 and 98 (89)<br>100 limit 99 to English language (84) | |
| **Web of Science**<br><br>TS=(an$emi*)<br><br>AND | **34** |

| |
|---|
| TS=(“point-of-care test*” OR “Point-of-care system*” OR “Point-of-care diagnostic*” OR “Point-of-care measurement*” OR “(rapid on-site NEAR/1 (evaluation* OR test*))” OR “h$emoglobin determination” OR “point-of-care Hb” OR “(point-of-care NEAR/2 h$emoglobin*)” OR POC OR POCT OR h$emoglobinometry OR HemoCue OR TrueHb OR “Sahli’s method” OR DiaSpect OR “Radical-7” OR “Pronto-7” OR I-STAT OR “Haemoglobin Colo$r Scale” OR “Hemoglobin Colo$r Scale” OR HCS OR colo$rimetry OR “GEM 4000” OR “Hemo Control” OR “STAT-Site MHgb” OR URIT-12 OR Aptus OR “AnemoCheck LRS” OR “AnemoCheck Low Resource Setting*” OR “Mission Hb” OR “Mission R plus”)<br><br>AND<br><br>TS=(afghanistan OR albania OR algeria OR “american samoa” OR angola OR argentina OR armenia OR armenian OR azerbaijan OR bangladesh OR “republic of belarus” OR belarus OR byelarus OR belorussia OR byelorussian OR belize OR “british Honduras” OR benin OR dahomey OR bhutan OR bolivia OR "bosnia and herzegovina" OR bosnia OR herzegovina OR botswana OR bechuanaland OR brazil OR brasil OR bulgaria OR burkina faso OR “burkina fasso” OR “upper volta” OR burundi OR urundi OR cabo verde OR cape verde OR cambodia OR kampuchea OR “khmer republic” OR cameroon OR cameron OR cameroun OR “central african republic” OR ubangi shari OR chad OR china OR colombia OR comoros OR “comoro islands” OR “iles comores” OR mayotte OR “democratic republic of the congo” OR “democratic republic congo” OR congo OR zaire OR “costa rica” OR "cote d’ivoire" OR "cote d’ ivoire" OR “cote divoire” OR “cote d ivoire” OR “ivory coast” OR cuba OR djibouti OR “french Somaliland” OR dominica OR “dominican republic” OR ecuador OR egypt OR “united arab republic” OR “el Salvador” OR “equatorial guinea” OR “spanish guinea” OR eritrea OR eswatini OR swaziland OR ethiopia OR fiji OR gabon OR gabonese republic OR gambia OR "georgia (republic)" OR georgian OR ghana OR “gold coast” OR grenada OR guam OR guatemala OR guinea OR guinea bissau OR guyana OR “british Guiana” OR haiti OR hispaniola OR honduras OR india OR indonesia OR timor OR iran OR iraq OR “isle of man” OR jamaica OR jordan OR kazakhstan OR kazakh OR kenya OR Kiribati OR "democratic people’s republic of korea" OR “republic of korea” OR “north korea” OR “south korea” OR korea OR kosovo OR kyrgyzstan OR kirghizia OR kirgizstan OR “kyrgyz republic OR kirghiz” OR laos OR “lao pdr” OR "lao people's democratic republic" OR lebanon OR lebanese republic OR lesotho OR basutoland OR liberia OR libya OR “libyan arab Jamahiriya” OR macau OR macao OR “republic of north Macedonia” OR macedonia OR madagascar OR “malagasy republic” OR malawi OR nyasaland OR malaysia OR “malay federation” OR “malaya federation” OR maldives OR “indian ocean islands” OR “indian ocean” OR mali OR micronesia OR “federated states of Micronesia” OR kiribati OR marshall islands OR nauru OR “northern mariana islands” OR palau OR tuvalu OR mauritania OR mauritius OR mexico OR moldova OR moldovian OR mongolia OR montenegro OR morocco OR ifni OR mozambique OR portuguese east africa OR myanmar OR burma OR namibia OR Nepal OR nicaragua OR niger OR nigeria OR pakistan OR panama OR “papua new guinea” OR “new guinea” OR paraguay OR peru OR philippines OR philipines OR phillipines OR phillippines OR romania OR russia OR “russian federation” OR ussr OR “soviet union” OR “union of soviet socialist republics” OR rwanda OR ruanda OR samoa OR “pacific islands” OR polynesia OR “samoan islands” OR “navigator island” OR “navigator islands” OR "sao tome and principe" OR senegal OR serbia OR “sierra leone” OR melanesia OR “solomon island” OR “solomon islands” OR “norfolk island” OR “norfolk islands” OR somalia OR “south Africa” OR “south sudan” OR “sri lanka” OR Ceylon OR “saint lucia” OR "st. lucia" OR "saint vincent and the grenadines" OR “saint Vincent” OR "st. vincent" OR grenadines OR sudan OR suriname OR surinam OR “dutch Guiana” OR “netherlands Guiana” OR syria OR “syrian arab republic” OR tajikistan OR tadjikistan OR |

| | |
|---|---|
| tadzhikistan OR tadzhik OR tanzania OR tanganyika OR thailand OR siam OR “timor leste” OR “east timor” OR togo OR “togolese republic” OR tonga OR tunisia OR turkey OR turkmenistan OR turkmen OR Tuvalu OR uganda OR ukraine OR uzbekistan OR uzbek OR vanuatu OR new hebrides OR venezuela OR vietnam OR viet nam OR “middle east” OR “west bank” OR gaza OR palestine OR yemen OR zambia OR zimbabwe OR “northern Rhodesia” OR “global south” OR “africa south of the sahara” OR “sub-saharan Africa” OR “subsaharan Africa” OR “africa, central” OR “central Africa” OR “africa, northern” OR “north Africa” OR “northern Africa” OR magreb OR maghrib OR sahara OR “africa, southern” OR “southern Africa” OR “africa, eastern” OR “east Africa” OR “eastern Africa” OR “africa, western” OR “west Africa” OR “western Africa” OR “west indies” OR “indian ocean islands” OR caribbean OR “central America” OR “latin America” OR "south and central america" OR “south America” OR “asia, central” OR “central asia” OR “asia, northern” OR “north asia” OR “northern asia” OR “asia, southeastern” OR “southeastern asia” OR “south eastern asia” OR “southeast asia” OR “south east asia” OR “asia, western” OR “western asia” OR “europe, eastern” OR “east Europe” OR “eastern Europe” OR “developing country” OR “developing countries” OR “developing nation?” OR “developing population?” OR “developing world” OR “less developed countr*” OR “less developed nation?” OR “less developed population?” OR “less developed world” OR “lesser developed countr*” OR “lesser developed nation?” OR “lesser developed population?” OR “lesser developed world” OR “under developed countr*” OR “under developed nation?” OR “under developed population?” OR “under developed world” OR “underdeveloped countr*” OR “underdeveloped nation?” OR “underdeveloped population?” OR “underdeveloped world” OR “middle income countr*” OR “middle income nation?” OR “middle income population?” OR “low income countr*” OR “low income nation?” OR “low income population?” OR “lower income countr*” OR “lower income nation?” OR “lower income population?” OR “underserved countr*” OR “underserved nation?” OR “underserved population?” OR “underserved world” OR “under served countr*” OR “under served nation?” OR “under served population?” OR “under served world” OR “deprived countr*” OR “deprived nation?” OR “deprived population?” OR “deprived world” OR “poor countr*” OR “poor nation?” OR “poor population?” OR “poor world” OR “poorer countr*” OR “poorer nation?” OR “poorer population?” OR “poorer world” OR “developing econom*” OR “less developed econom*” OR “lesser developed econom*” OR “under developed econom*” OR “underdeveloped econom*” OR “middle income econom*” OR “low income econom*” OR “lower income econom*” OR “low gdp” OR “low gnp” OR “low gross domestic” OR “low gross national” OR “lower gdp” OR “lower gnp” OR “lower gross domestic” OR “lower gross national” OR lmic OR lmics OR “third world” OR “lami countr*” OR “transitional countr*” OR “emerging economies” OR “emerging nation?”)<br><br>AND<br><br>TS=(hospital* OR “(community NEAR/2 Health)” OR “(clinic or clinics)” OR “public health facilit*”) | |
| **CENTRAL**<br><br>(Anaemi* OR anemi*):ti,ab,kw<br><br>AND | **101** |

| |
|---|
| (“point-of-care test” OR “point-of-care tests” OR “point-of-care testing” OR “Point-of-care system” OR “point-of-care systems” OR “Point-of-care diagnostic” OR “point-of-care diagnostics” OR “Point-of-care measurement” OR “point-of-care measurements” OR “rapid on-site evaluation” OR “rapid on-site evaluations” OR “rapid on-site test” OR “rapid on-site tests” OR “rapid on-site testing” OR “haemoglobin determination” OR “hemoglobin determination” OR “point-of-care Hb” OR “point-of-care haemoglobin” OR “point-of-care hemoglobin” OR “point-of-care haemoglobinometry” OR “point-of-care hemoglobinometry” OR POC OR POCT OR haemoglobinometry OR hemoglobinometry OR HemoCue OR TrueHb OR “Sahli’s method” OR DiaSpect OR “Radical-7” OR “Pronto-7” OR I-STAT OR “Haemoglobin Colour Scale” OR “hemoglobin Colour Scale” OR “haemoglobin Color Scale” OR “hemoglobin color Scale” OR HCS OR colourimetry OR colorimetry OR “GEM 4000” OR “Hemo Control” OR “STAT-Site MHgb” OR URIT-12 OR Aptus OR “AnemoCheck LRS” OR “AnemoCheck Low Resource Setting” OR “AnemoCheck Low Resource Settings” OR “Mission Hb” OR “Mission R plus”):ti,ab,kw<br><br>AND<br><br>(afghanistan OR albania OR algeria OR "american samoa" OR angola OR argentina OR armenia OR armenian OR azerbaijan OR bangladesh OR "republic of belarus" OR belarus OR byelarus OR belorussia OR byelorussian OR belize OR "british Honduras" OR benin OR dahomey OR bhutan OR bolivia OR "bosnia and herzegovina" OR bosnia OR herzegovina OR botswana OR bechuanaland OR brazil OR brasil OR bulgaria OR burkina faso OR "burkina fasso" OR "upper volta" OR burundi OR urundi OR cabo verde OR cape verde OR cambodia OR kampuchea OR "khmer republic" OR cameroon OR cameron OR cameroun OR "central african republic" OR ubangi shari OR chad OR china OR colombia OR comoros OR "comoro islands" OR "iles comores" OR mayotte OR "democratic republic of the congo" OR "democratic republic congo" OR congo OR zaire OR "costa rica" OR "cote d’ivoire" OR "cote d’ ivoire" OR "cote divoire" OR "cote d ivoire" OR "ivory coast" OR cuba OR djibouti OR "french Somaliland" OR dominica OR "dominican republic" OR ecuador OR egypt OR "united arab republic" OR "el Salvador" OR "equatorial guinea" OR "spanish guinea" OR eritrea OR eswatini OR swaziland OR ethiopia OR fiji OR gabon OR gabonese republic OR gambia OR "georgia (republic)" OR georgian OR ghana OR "gold coast" OR grenada OR guam OR guatemala OR guinea OR guinea bissau OR guyana OR "british Guiana" OR haiti OR hispaniola OR honduras OR india OR indonesia OR timor OR iran OR iraq OR "isle of man" OR jamaica OR jordan OR kazakhstan OR kazakh OR kenya OR Kiribati OR "democratic people’s republic of korea" OR "republic of korea" OR "north korea" OR "south korea" OR korea OR kosovo OR kyrgyzstan OR kirghizia OR kirgizstan OR "kyrgyz republic OR kirghiz" OR laos OR "lao pdr" OR "lao people's democratic republic" OR lebanon OR lebanese republic OR lesotho OR basutoland OR liberia OR libya OR "libyan arab Jamahiriya" OR macau OR macao OR "republic of north Macedonia" OR macedonia OR madagascar OR "malagasy republic" OR malawi OR nyasaland OR malaysia OR "malay federation" OR "malaya federation" OR maldives OR "indian ocean islands" OR "indian ocean" OR mali OR micronesia OR "federated states of Micronesia" OR kiribati OR marshall islands OR nauru OR "northern mariana islands" OR palau OR tuvalu OR mauritania OR mauritius OR mexico OR moldova OR moldovian OR mongolia OR montenegro OR morocco OR ifni OR mozambique OR portuguese east africa OR myanmar OR burma OR namibia OR Nepal OR nicaragua OR niger OR nigeria OR pakistan OR panama OR "papua new guinea" OR "new guinea" OR paraguay OR peru OR philippines OR philipines OR phillipines OR phillippines OR romania OR russia OR "russian federation" OR ussr OR "soviet union" OR "union of soviet socialist republics" OR rwanda OR ruanda OR samoa OR "pacific islands" OR polynesia OR "samoan islands" OR "navigator island" OR "navigator |

| |
|---|
| islands" OR "sao tome and principe" OR senegal OR serbia OR "sierra leone" OR melanesia OR "solomon island" OR "solomon islands" OR "norfolk island" OR "norfolk islands" OR somalia OR "south Africa" OR "south sudan" OR "sri lanka" OR Ceylon OR "saint lucia" OR "st. lucia" OR "saint vincent and the grenadines" OR "saint Vincent" OR "st. vincent" OR grenadines OR sudan OR suriname OR surinam OR "dutch Guiana" OR "netherlands Guiana" OR syria OR "syrian arab republic" OR tajikistan OR tadjikistan OR tadzhikistan OR tadzhik OR tanzania OR tanganyika OR thailand OR siam OR "timor leste" OR "east timor" OR togo OR "togolese republic" OR tonga OR tunisia OR turkey OR turkmenistan OR turkmen OR Tuvalu OR uganda OR ukraine OR uzbekistan OR uzbek OR vanuatu OR new hebrides OR venezuela OR vietnam OR viet nam OR “middle east” OR “west bank” OR gaza OR palestine OR yemen OR zambia OR zimbabwe OR “northern Rhodesia” OR "global south" OR "africa south of the sahara" OR "sub-saharan Africa" OR "subsaharan Africa" OR "africa, central" OR "central Africa" OR "africa, northern" OR "north Africa" OR "northern Africa" OR magreb OR maghrib OR sahara OR "africa, southern" OR "southern Africa" OR "africa, eastern" OR "east Africa" OR "eastern Africa" OR "africa, western" OR "west Africa" OR "western Africa" OR "west indies" OR "indian ocean islands" OR caribbean OR "central America" OR "latin America" OR "south and central america" OR "south America" OR "asia, central" OR "central asia" OR "asia, northern" OR "north asia" OR "northern asia" OR "asia, southeastern" OR "southeastern asia" OR "south eastern asia" OR "southeast asia" OR "south east asia" OR "asia, western" OR "western asia" OR "europe, eastern" OR "east Europe" OR "eastern Europe" OR "developing country" OR "developing countries" OR "developing nation" OR "developing nations" OR "developing population" OR "developing populations" OR "developing world" OR "less developed country" OR "less developed countries" OR "less developed nation" OR "less developed nations" OR "less developed population" OR "less developed populations" OR "less developed world" OR "lesser developed country" OR "lesser developed countries" OR "lesser developed nation" OR "lesser developed nations" OR "lesser developed population" OR "lesser developed populations" OR "lesser developed world" OR "under developed country" OR "under developed countries" OR "under developed nation" OR "under developed nations" OR "under developed population" OR "under developed populations" OR "under developed world" OR "underdeveloped country" OR "underdeveloped countries" OR "underdeveloped nation" OR "underdeveloped nations" OR "underdeveloped population" OR "underdeveloped populations" OR "underdeveloped world" OR "middle income country" OR "middle income countries" OR "middle income nation" OR "middle income nations" OR "middle income population" OR "middle income populations" OR "low income country" OR "low income countries" OR "low income nation" OR "low income nations" OR "low income population" OR "low income populations" OR "lower income country" OR "lower income countries" OR "lower income nation" OR "lower income nations" OR "lower income population" OR "lower income populations" OR "underserved country" OR "underserved countries" OR "underserved nation" OR "underserved nations" OR "underserved population" OR "underserved populations" OR "underserved world" OR "under served country" OR "under served countries" OR "under served nation" OR "under served nations" OR "under served population" OR "under served populations" OR "under served world" OR "deprived country" OR "deprived countries" OR "deprived nation" OR "deprived nations" OR "deprived population" OR "deprived populations" OR "deprived world" OR "poor country" OR "poor countries" OR "poor nation" OR "poor nations" OR "poor population" OR "poor populations" OR "poor world" OR "poorer country" OR "poorer countries" OR "poorer nation" OR "poorer nation" OR "poorer population" OR "poorer populations" OR "poorer world" OR "developing economy" OR "developing economies" OR "less developed economy" OR "less developed economies" OR "lesser developed economy" OR "lesser |

| | |
|---|---|
| developed economies" OR "under developed economy" OR "under developed economies" OR "underdeveloped economy" OR "underdeveloped economies" OR "middle income economy" OR "middle income economies" OR "low income economy" OR "low income economies" OR "lower income economy" OR "lower income economies" OR "low gdp" OR "low gnp" OR "low gross domestic" OR "low gross national" OR "lower gdp" OR "lower gnp" OR "lower gross domestic" OR "lower gross national" OR lmic OR lmics OR "third world" OR "lami country" OR "lami countries" OR "transitional country" OR "transitional countries" OR "emerging economies" OR "emerging nation" OR "emerging nations"):ti,ab,kw<br><br>AND<br><br>(hospital* OR “(community NEAR/2 Health)” OR “(clinic or clinics)” OR “public health facility” OR “public health facilities”):ti,ab,kw | |
| **ProQuest Dissertations and Thesis**<br>Anywhere except full text:<br><br>(An*emi*)<br><br>AND<br><br>(“point-of-care test” OR “point-of-care tests” OR “point-of-care testing” OR “Point-of-care system” OR “point-of-care systems” OR “Point-of-care diagnostic” OR “point-of-care diagnostics” OR “Point-of-care measurement” OR “point-of-care measurements” OR “rapid on-site evaluation” OR “rapid on-site evaluations” OR “rapid on-site test” OR “rapid on-site tests” OR “rapid on-site testing” OR “haemoglobin determination” OR “hemoglobin determination” OR “point-of-care Hb” OR “point-of-care haemoglobin” OR “point-of-care hemoglobin” OR “point-of-care haemoglobinometry” OR “point-of-care hemoglobinometry” OR POC OR POCT OR haemoglobinometry OR hemoglobinometry OR HemoCue OR TrueHb OR “Sahli’s method” OR DiaSpect OR “Radical-7” OR “Pronto-7” OR I-STAT OR “Haemoglobin Colour Scale” OR “hemoglobin Colour Scale” OR “haemoglobin Color Scale” OR “hemoglobin color Scale” OR HCS OR colourimetry OR colorimetry OR “GEM 4000” OR “Hemo Control” OR “STAT-Site MHgb” OR URIT-12 OR Aptus OR “AnemoCheck LRS” OR “AnemoCheck Low Resource Setting” OR “AnemoCheck Low Resource Settings” OR “Mission Hb” OR “Mission R plus”)<br><br>AND<br><br>(afghanistan OR albania OR algeria OR “american samoa” OR angola OR argentina OR armenia OR armenian OR azerbaijan OR bangladesh OR “republic of belarus” OR belarus OR byelarus OR belorussia OR byelorussian OR belize OR “british Honduras” OR benin OR dahomey OR bhutan OR bolivia OR "bosnia and herzegovina" OR bosnia OR herzegovina OR botswana OR bechuanaland OR brazil OR brasil OR bulgaria OR burkina faso OR “burkina fasso” OR “upper volta” OR burundi OR urundi OR cabo verde OR cape verde OR cambodia OR kampuchea OR “khmer republic” OR cameroon OR cameron OR cameroun OR “central african republic” OR ubangi shari OR chad OR china OR colombia OR comoros OR “comoro islands” OR “iles comores” OR mayotte OR “democratic republic of the congo” OR “democratic republic congo” OR congo OR zaire OR “costa rica” OR "cote | **2** |

| |
|---|
| d’ivoire" OR "cote d’ ivoire" OR “cote divoire” OR “cote d ivoire” OR “ivory coast” OR cuba OR djibouti OR “french Somaliland” OR dominica OR “dominican republic” OR ecuador OR egypt OR “united arab republic” OR “el Salvador” OR “equatorial guinea” OR “spanish guinea” OR eritrea OR eswatini OR swaziland OR ethiopia OR fiji OR gabon OR gabonese republic OR gambia OR "georgia (republic)" OR georgian OR ghana OR “gold coast” OR grenada OR guam OR guatemala OR guinea OR guinea bissau OR guyana OR “british Guiana” OR haiti OR hispaniola OR honduras OR india OR indonesia OR timor OR iran OR iraq OR “isle of man” OR jamaica OR jordan OR kazakhstan OR kazakh OR kenya OR Kiribati OR "democratic people’s republic of korea" OR “republic of korea” OR “north korea” OR “south korea” OR korea OR kosovo OR kyrgyzstan OR kirghizia OR kirgizstan OR “kyrgyz republic OR kirghiz” OR laos OR “lao pdr” OR "lao people's democratic republic" OR lebanon OR lebanese republic OR lesotho OR basutoland OR liberia OR libya OR “libyan arab Jamahiriya” OR macau OR macao OR “republic of north Macedonia” OR macedonia OR madagascar OR “malagasy republic” OR malawi OR nyasaland OR malaysia OR “malay federation” OR “malaya federation” OR maldives OR “indian ocean islands” OR “indian ocean” OR mali OR micronesia OR “federated states of Micronesia” OR kiribati OR marshall islands OR nauru OR “northern mariana islands” OR palau OR tuvalu OR mauritania OR mauritius OR mexico OR moldova OR moldovian OR mongolia OR montenegro OR morocco OR ifni OR mozambique OR portuguese east africa OR myanmar OR burma OR namibia OR Nepal OR nicaragua OR niger OR nigeria OR pakistan OR panama OR “papua new guinea” OR “new guinea” OR paraguay OR peru OR philippines OR philipines OR phillipines OR phillippines OR romania OR russia OR “russian federation” OR ussr OR “soviet union” OR “union of soviet socialist republics” OR rwanda OR ruanda OR samoa OR “pacific islands” OR polynesia OR “samoan islands OR navigator island” OR “navigator islands” OR "sao tome and principe" OR senegal OR serbia OR “sierra leone” OR melanesia OR “solomon island” OR “solomon islands” OR “norfolk island” OR “norfolk islands” OR somalia OR “south Africa” OR “south sudan” OR “sri lanka” OR Ceylon OR “saint lucia” OR "st. lucia" OR "saint vincent and the grenadines" OR “saint Vincent” OR "st. vincent" OR grenadines OR sudan OR suriname OR surinam OR “dutch Guiana” OR “netherlands Guiana” OR syria OR “syrian arab republic” OR tajikistan OR tadjikistan OR tadzhikistan OR tadzhik OR tanzania OR tanganyika OR thailand OR siam OR “timor leste” OR “east timor” OR togo OR “togolese republic” OR tonga OR tunisia OR turkey OR turkmenistan OR turkmen OR Tuvalu OR uganda OR ukraine OR uzbekistan OR uzbek OR vanuatu OR new hebrides OR venezuela OR vietnam OR viet nam OR “middle east” OR “west bank” OR gaza OR palestine OR yemen OR zambia OR zimbabwe OR “northern Rhodesia” OR “global south” OR “africa south of the sahara” OR “sub-saharan Africa” OR “subsaharan Africa” OR “africa, central” OR “central Africa” OR “africa, northern” OR “north Africa” OR “northern Africa” OR magreb OR maghrib OR sahara OR “africa, southern” OR “southern Africa” OR “africa, eastern” OR “east Africa” OR “eastern Africa” OR “africa, western” OR “west Africa” OR “western Africa” OR “west indies” OR “indian ocean islands” OR caribbean OR “central America” OR “latin America” OR "south and central america" OR “south America” OR “asia, central” OR “central asia” OR “asia, northern” OR “north asia” OR “northern asia” OR “asia, southeastern” OR “southeastern asia” OR “south eastern asia” OR “southeast asia” OR “south east asia” OR “asia, western” OR “western asia” OR “europe, eastern” OR “east Europe” OR “eastern Europe” OR “developing country” OR “developing countries” OR “developing nation?” OR “developing population?” OR “developing world” OR “less developed countr*” OR “less developed nation?” OR “less developed population?” OR “less developed world” OR “lesser developed countr*” OR “lesser developed nation?” OR “lesser developed population?” OR “lesser developed world” OR “under developed countr*” OR “under developed nation?” OR “under developed population?” OR “under developed world” OR |

| | |
|---|---|
| “underdeveloped countr*” OR “underdeveloped nation?” OR “underdeveloped population?” OR “underdeveloped world” OR “middle income countr*” OR “middle income nation?” OR “middle income population?” OR “low income countr*” OR “low income nation?” OR “low income population?” OR “lower income countr*” OR “lower income nation?” OR “lower income population?” OR “underserved countr*” OR “underserved nation?” OR “underserved population?” OR “underserved world” OR “under served countr*” OR “under served nation?” OR “under served population?” OR “under served world” OR “deprived countr*” OR “deprived nation?” OR “deprived population?” OR “deprived world OR poor countr*” OR “poor nation?” OR “poor population?” OR “poor world” OR “poorer countr*” OR “poorer nation?” OR “poorer population?” OR “poorer world” OR “developing econom*” OR “less developed econom*” OR “lesser developed econom*” OR “under developed econom*” OR “underdeveloped econom*” OR “middle income econom*” OR “low income econom*” OR “lower income econom*” OR “low gdp” OR “low gnp” OR “low gross domestic” OR “low gross national” OR “lower gdp” OR “lower gnp” OR “lower gross domestic” OR “lower gross national” OR lmic OR lmics OR “third world” OR “lami countr*” OR “transitional countr*” OR “emerging economies” OR “emerging nation?”)<br><br>AND<br><br>hospital* OR “(community NEAR/2 Health)” OR “(clinic or clinics)” OR “public health facilit*” | |
| **LILACS (VHL)**<br>Bold: DeCS (meSH) terms<br><br>TW:(**anemia** OR anaemi* OR anemi*)<br><br>AND<br><br>TW:("point-of-care test" OR "point-of-care tests" OR "**point-of-care testing**" OR "Point-of-care system" OR "**point-of-care systems**" OR "Point-of-care diagnostic" OR "point-of-care diagnostics" OR "Point-of-care measurement" OR "point-of-care measurements" OR "rapid on-site evaluation" OR "rapid on-site evaluations" OR "rapid on-site test" OR "rapid on-site tests" OR "rapid on-site testing" OR "haemoglobin determination" OR "hemoglobin determination" OR "point-of-care Hb" OR "point-of-care haemoglobin" OR "point-of-care hemoglobin" OR "point-of-care haemoglobinometry" OR "point-of-care hemoglobinometry" OR POC OR POCT OR **haemoglobinometry** OR hemoglobinometry OR HemoCue OR TrueHb OR "Sahli’s method" OR DiaSpect OR "Radical-7" OR "Pronto-7" OR I-STAT OR "Haemoglobin Colour Scale" OR "hemoglobin Colour Scale" OR "haemoglobin Color Scale" OR "hemoglobin color Scale" OR HCS OR colourimetry OR **colorimetry** OR "GEM 4000" OR "Hemo Control" OR "STAT-Site MHgb" OR URIT-12 OR Aptus OR "AnemoCheck LRS" OR "AnemoCheck Low Resource Setting" OR "AnemoCheck Low Resource Settings" OR "Mission Hb" OR "Mission R plus")<br><br>AND<br><br>TW:(afghanistan OR albania OR algeria OR "american samoa" OR angola OR argentina OR armenia OR armenian OR azerbaijan OR bangladesh OR "republic of belarus" OR belarus OR byelarus OR belorussia OR byelorussian OR belize OR "british Honduras" OR benin OR | **0** |

| |
|---|
| dahomey OR bhutan OR bolivia OR "bosnia and herzegovina" OR bosnia OR herzegovina OR botswana OR bechuanaland OR brazil OR brasil OR bulgaria OR burkina faso OR "burkina fasso" OR "upper volta" OR burundi OR urundi OR cabo verde OR cape verde OR cambodia OR kampuchea OR "khmer republic" OR cameroon OR cameron OR cameroun OR "central african republic" OR ubangi shari OR chad OR china OR colombia OR comoros OR "comoro islands" OR "iles comores" OR mayotte OR "democratic republic of the congo" OR "democratic republic congo" OR congo OR zaire OR "costa rica" OR "cote d'ivoire" OR "cote d' ivoire" OR "cote divoire" OR "cote d ivoire" OR "ivory coast" OR cuba OR djibouti OR "french Somaliland" OR dominica OR "dominican republic" OR ecuador OR egypt OR "united arab republic" OR "el Salvador" OR "equatorial guinea" OR "spanish guinea" OR eritrea OR eswatini OR swaziland OR ethiopia OR fiji OR gabon OR gabonese republic OR gambia OR "georgia (republic)" OR georgian OR ghana OR "gold coast" OR grenada OR guam OR guatemala OR guinea OR guinea bissau OR guyana OR "british Guiana" OR haiti OR hispaniola OR honduras OR india OR indonesia OR timor OR iran OR iraq OR "isle of man" OR jamaica OR jordan OR kazakhstan OR kazakh OR kenya OR Kiribati OR "democratic people's republic of korea" OR "republic of korea" OR "north korea" OR "south korea" OR korea OR kosovo OR kyrgyzstan OR kirghizia OR kirgizstan OR "kyrgyz republic OR kirghiz" OR laos OR "lao pdr" OR "lao people's democratic republic" OR lebanon OR lebanese republic OR lesotho OR basutoland OR liberia OR libya OR "libyan arab Jamahiriya" OR macau OR macao OR "republic of north Macedonia" OR macedonia OR madagascar OR "malagasy republic" OR malawi OR nyasaland OR malaysia OR "malay federation" OR "malaya federation" OR maldives OR "indian ocean islands" OR "indian ocean" OR mali OR micronesia OR "federated states of Micronesia" OR kiribati OR marshall islands OR nauru OR "northern mariana islands" OR palau OR tuvalu OR mauritania OR mauritius OR mexico OR moldova OR moldovian OR mongolia OR montenegro OR morocco OR ifni OR mozambique OR portuguese east africa OR myanmar OR burma OR namibia OR Nepal OR nicaragua OR niger OR nigeria OR pakistan OR panama OR "papua new guinea" OR "new guinea" OR paraguay OR peru OR philippines OR philipines OR phillipines OR phillippines OR romania OR russia OR "russian federation" OR ussr OR "soviet union" OR "union of soviet socialist republics" OR rwanda OR ruanda OR samoa OR "pacific islands" OR polynesia OR "samoan islands" OR "navigator island" OR "navigator islands" OR "sao tome and principe" OR senegal OR serbia OR "sierra leone" OR melanesia OR "solomon island" OR "solomon islands" OR "norfolk island" OR "norfolk islands" OR somalia OR "south Africa" OR "south sudan" OR "sri lanka" OR Ceylon OR "saint lucia" OR "st. lucia" OR "saint vincent and the grenadines" OR "saint Vincent" OR "st. vincent" OR grenadines OR sudan OR suriname OR surinam OR "dutch Guiana" OR "netherlands Guiana" OR syria OR "syrian arab republic" OR tajikistan OR tadjikistan OR tadzhikistan OR tadzhik OR tanzania OR tanganyika OR thailand OR siam OR "timor leste" OR "east timor" OR togo OR "togolese republic" OR tonga OR tunisia OR turkey OR turkmenistan OR turkmen OR Tuvalu OR uganda OR ukraine OR uzbekistan OR uzbek OR vanuatu OR new hebrides OR venezuela OR vietnam OR viet nam OR "middle east" OR "west bank" OR gaza OR palestine OR yemen OR zambia OR zimbabwe OR "northern Rhodesia" OR "global south" OR "**africa south of the sahara**" OR "sub-saharan Africa" OR "subsaharan Africa" OR "africa, central" OR "central Africa" OR "africa, northern" OR "north Africa" OR "northern Africa" OR magreb OR maghrib OR sahara OR "africa, southern" OR "southern Africa" OR "africa, eastern" OR "east Africa" OR "eastern Africa" OR "africa, western" OR "west Africa" OR "western Africa" OR "west indies" OR "indian ocean islands" OR caribbean OR "central America" OR "latin America" OR "south and central america" OR "south America" OR "asia, central" OR "central asia" OR "asia, northern" OR "north asia" OR "northern asia" OR "asia, southeastern" OR "southeastern asia" OR "south eastern asia" OR "southeast asia" OR "south east asia" OR "asia, western" OR "western asia" OR |

| | |
|---|---|
| "europe, eastern" OR "east Europe" OR "eastern Europe" OR "developing country" OR "**developing countries**" OR "developing nation" OR "developing nations" OR "developing population" OR "developing populations" OR "developing world" OR "less developed country" OR "less developed countries" OR "less developed nation" OR "less developed nations" OR "less developed population" OR "less developed populations" OR "less developed world" OR "lesser developed country" OR "lesser developed countries" OR "lesser developed nation" OR "lesser developed nations" OR "lesser developed population" OR "lesser developed populations" OR "lesser developed world" OR "under developed country" OR "under developed countries" OR "under developed nation" OR "under developed nations" OR "under developed population" OR "under developed populations" OR "under developed world" OR "underdeveloped country" OR "underdeveloped countries" OR "underdeveloped nation" OR "underdeveloped nations" OR "underdeveloped population" OR "underdeveloped populations" OR "underdeveloped world" OR "middle income country" OR "middle income countries" OR "middle income nation" OR "middle income nations" OR "middle income population" OR "middle income populations" OR "low income country" OR "low income countries" OR "low income nation" OR "low income nations" OR "low income population" OR "low income populations" OR "lower income country" OR "lower income countries" OR "lower income nation" OR "lower income nations" OR "lower income population" OR "lower income populations" OR "underserved country" OR "underserved countries" OR "underserved nation" OR "underserved nations" OR "underserved population" OR "underserved populations" OR "underserved world" OR "under served country" OR "under served countries" OR "under served nation" OR "under served nations" OR "under served population" OR "under served populations" OR "under served world" OR "deprived country" OR "deprived countries" OR "deprived nation" OR "deprived nations" OR "deprived population" OR "deprived populations" OR "deprived world" OR "poor country" OR "poor countries" OR "poor nation" OR "poor nations" OR "poor population" OR "poor populations" OR "poor world" OR "poorer country" OR "poorer countries" OR "poorer nation" OR "poorer nation" OR "poorer population" OR "poorer populations" OR "poorer world" OR "developing economy" OR "developing economies" OR "less developed economy" OR "less developed economies" OR "lesser developed economy" OR "lesser developed economies" OR "under developed economy" OR "under developed economies" OR "underdeveloped economy" OR "underdeveloped economies" OR "middle income economy" OR "middle income economies" OR "low income economy" OR "low income economies" OR "lower income economy" OR "lower income economies" OR "low gdp" OR "low gnp" OR "low gross domestic" OR "low gross national" OR "lower gdp" OR "lower gnp" OR "lower gross domestic" OR "lower gross national" OR lmic OR lmics OR "third world" OR "lami country" OR "lami countries" OR "transitional country" OR "transitional countries" OR "emerging economies" OR "emerging nation" OR "emerging nations")<br><br>AND<br><br>TW:(**hospitals** OR hospital OR "community health" OR clinic* OR **health facilities** OR **community health centers** OR "public health facilities" OR "public health facility") | |
| **WHO ICTRP**<br><br>Title: (“point-of-care test*” OR “Point-of-care system*” OR “Point-of-care diagnostic*” OR “Point-of-care measurement*” OR “rapid on-site evaluation*” OR “rapid on-site test*” | **8** |

| | |
|---|---|
| OR “haemoglobin determination” OR “hemoglobin determination” OR “point-of-care Hb” OR “point-of-care haemoglobin” OR “point-of-care hemoglobin” OR “point-of-care haemoglobinometry” OR “point-of-care hemoglobinometry” OR POC OR POCT OR haemoglobinometry OR hemoglobinometry OR HemoCue OR TrueHb OR “Sahli’s method” OR DiaSpect OR “Radical-7” OR “Pronto-7” OR I-STAT OR “Haemoglobin Colour Scale” OR “hemoglobin Colour Scale” OR “haemoglobin Color Scale” OR “hemoglobin color Scale” OR HCS OR colourimetry OR colorimetry OR “GEM 4000” OR “Hemo Control” OR “STAT-Site MHgb” OR URIT-12 OR Aptus OR “AnemoCheck LRS” OR “AnemoCheck Low Resource Setting” OR “AnemoCheck Low Resource Settings” OR “Mission Hb” OR “Mission R plus”)<br><br>Condition: Anaemi* OR anemi* | |
| **ClinicalTrials.gov**<br><br>Condition: Anaemia OR anemia OR anaemic OR anemic OR anaemias OR anemias<br><br>Targeted intervention: “point-of-care test” OR “point-of-care tests” OR “point-of-care testing” OR “Point-of-care system” OR “point-of-care systems” OR “Point-of-care diagnostic” OR “point-of-care diagnostics” OR “Point-of-care measurement” OR hemocue | **16** |

**Supplementary Table S2: Data Extraction Table of Included Study Characteristics**

| Study and publication/ registration year | Study design | Aim of study | Population and sample size (for those undergoing POC Hb testing if provided) | Location and setting | POC Hb test(s) used (blood sample and timing if provided) | Percentage of patients diagnosed with mild/moderate /severe/overall anaemia and/or mean Hb concentration by POC(Hb)T |
|---|---|---|---|---|---|---|
| 1. Alamneh *et al.*, 2021[3] | Cross-sectional | To assess the magnitude and associated factors of anaemia among children aged 6-59 months attending at Debre Markos Referral Hospital, Northwest Ethiopia | 310 children 6-59 months (<5 years) (All children included, except those with severe medical conditions) | Debre Markos Referral Hospital, Northwest Ethiopia | HemoCue 301 (capillary and venous) | Mild (10-10.9 g/dl): 7.1%<br><br>Moderate (7-9.9g/dl): 3.5%<br><br>Severe (<7g/dl): 1.3%<br><br>Overall: 11.9% (95% CI: 8.5-16.2) |
| 2. Aldridge *et al.*, 2012[4] | Diagnostic study | To estimate the diagnostic accuracy of HCS in preschool-age children when used in real-life conditions, relative to a reference method (HemoCue) and to the Integrated Management of Childhood Illness (IMCI) recommended method (clinical examination for palmar pallor) | 788 children aged 2-59 months (All children included) | 6 Mother and Child Health Clinics on Pemba Island in the Zanzibar archipelago, Tanzania (Gombani, Kiuyu, Shidi, Vumba, Bogoa and Kiwani) | HCS (capillary) and HemoCue (capillary) | Severe (<5g/dl): 0.8% (95% CI: 0.3-1.6)<br><br>Overall (<11g/dl): 71% (95% CI: 68-74)<br><br>Mean Hb (HemoCue): 10.1g/dl (95% CI: 10.0-10.2)<br>Mean Hb (HCS): 11.6 g/dl (95% CI: 11.5-11.8) |
| 3. Bojang *et al.*, 2011[5] | RCT | To compare the effectiveness of delivery of intermittent preventative | 12,326 children 6 months - 6 years | 26 rural reproductive and child health | HemoCue | Mean±SD Hb:<br><br>RCH (n=525) |

| Study and publication/ registration year | Study design | Aim of study | Population and sample size (for those undergoing POC Hb testing if provided) | Location and setting | POC Hb test(s) used (blood sample and timing if provided) | Percentage of patients diagnosed with mild/moderate /severe/overall anaemia and/or mean Hb concentration by POC(Hb)T |
|---|---|---|---|---|---|---|
| | | treatment for malaria to Gambian children <6 years by virtual health workers (VHWs) and reproductive and child health (RCH) trekking teams | 68 children admitted to one of the health facilities in the study area during follow-up period and therefore had Hb assessed (30 in RCH and 36 in VHW) | clinics on the south bank of Upper River Region, The Gambia | (Secondary endpoint of study – morbidity surveillance – Hb measured when referred to health facility) | 10.2±2.0g/dl<br><br>VHW (n=571)<br>10.4±2.0 g/dl<br><br>Moderate anaemia (Hb<7g/dl):<br>RCH: 6.1%<br>VHW: 4.8% |
| 4. Bond *et al.*, 2014[6] | Diagnostic study | To evaluate the accuracy of HemoSpec in the field | 70 children aged 0-15 years | Paediatric ward of Queen Elizabeth Central Hospital, Blantyre, Malawi | HemoSpec (capillary) | Not reported |
| 5. Calis *et al.*, 2016[7] | Case-control | To assess the causative factors in Malawian children with SA | 1138 children aged 6-60 months (case patients with SA (Hb<5g/dl) = 381, community control (healthy children | Outpatient department at Chikwawa District Hospital (rural) and Queen Elizabeth Referral Central Hospital (urban | HemoCue | Mean±SD Hb:<br><br>Case patients:<br>3.6±0.8g/dl<br><br>Community control:<br>9.9±1.9g/dl |

| Study and publication/ registration year | Study design | Aim of study | Population and sample size (for those undergoing POC Hb testing if provided) | Location and setting | POC Hb test(s) used (blood sample and timing if provided) | Percentage of patients diagnosed with mild/moderate /severe/overall anaemia and/or mean Hb concentration by POC(Hb)T |
|---|---|---|---|---|---|---|
| | | | Hb>5g/dl) = 380, hospital control (Hb>5g/dl)= 377) | Blantyre), Malawi | | Hospital control: 9.6±2.2g/dl |
| 6. Choukem *et al.,* 2020[8] | Diagnostic study | To evaluate the technical and clinical accuracy of two POC haemoglobin meters and their agreement with a reference analyser in diagnosing anaemia in the African setting | All children and adults aged 4 -85 years<br><br>(12 children aged 4-14 years)<br>(216 aged >15 years) | Inpatient and Outpatient departments at Douala General Hospital, in Douala capital city of Cameroon | URIT-12 (capillary) and Mission Hb (capillary) | N/A - only reported for pooled children and adults sample |
| 7. Cusick *et al.*, 2016[9] | RCT | 1) Determine values of iron and inflammatory markers in children with severe malarial anaemia (SMA) or cerebral malaria (CM) and also in healthy community children (CC) before antimalarial treatment<br>2) Establish whether the 28-day values of iron and inflammatory markers differed between children in each group who received iron with their antimalarial treatment | 239 children aged 18 months- 5 years<br>(SMA = 77, CM = 79,<br>CC = 83) | Mulago hospital, Kampala, Uganda | HemoCue (venous)<br><br>Baseline and hospital follow-up at 28 days | Mean±SD Hb:<br><br>SMA: 3.74±0.95 g/dl<br><br>CM: 6.99±1.89 g/dl<br><br>CC: 11.72±1.23 g/dl |

| Study and publication/ registration year | Study design | Aim of study | Population and sample size (for those undergoing POC Hb testing if provided) | Location and setting | POC Hb test(s) used (blood sample and timing if provided) | Percentage of patients diagnosed with mild/moderate /severe/overall anaemia and/or mean Hb concentration by POC(Hb)T |
|---|---|---|---|---|---|---|
| | | compared with those who received antimalarial treatment alone | | | | |
| 8. Cusick *et al.*, 2016[10]<br><br>NCT01754701* | RCT | To determine whether iron supplementation given concurrently with antimalarial treatment compared with 4 weeks after antimalarial treatment is better incorporated into RBCs and improves haematologic recovery at 8 weeks | 100 children aged 6-59 months with fever and malaria | Paediatric Acute Care Unit at Mulago Hospital, Kampala, Uganda | HemoCue<br><br>Baseline – to assess trial eligibility | Mean Hb: 8.0 g/dl |
| 9. Degarege *et al.*, 2014[11] | Cross-sectional | To assess the associations between helminth infections and ABO blood group, anaemia, and undernutrition in febrile outpatients of all age groups | Febrile children and adults with suspected malaria (those with confirmed malaria excluded)<br><br>(142 children aged 1-15 years)<br><br>(333 aged 16-80 years) | Outpatient department at Dore Bafeno Health Centre, Dore Bafeno District, Southern Ethiopia | HemoCue 201 (capillary) | N/A - only reported for pooled children and adults sample |

| Study and publication/ registration year | Study design | Aim of study | Population and sample size (for those undergoing POC Hb testing if provided) | Location and setting | POC Hb test(s) used (blood sample and timing if provided) | Percentage of patients diagnosed with mild/moderate /severe/overall anaemia and/or mean Hb concentration by POC(Hb)T |
|---|---|---|---|---|---|---|
| 10. De Wit *et al.*, 2016[12]<br><br>NCT02741024 | RCT | To investigate whether increased treatment failure rates contributed to the apparent increase in malaria diagnoses in Baraka, South Kivu, DRC, by comparing the efficacy of Artesunate-amodiaquine (ASAQ) with Artemether-lumefantrine (AL) in children aged 6-59 months with confirmed uncomplicated falciparum malaria | 288 children with fever and uncomplicated malaria aged 6-59 months<br><br>ASAQ (n=144)<br>AL (n=144) | Outpatient clinic of Baraka General Hospital and Health Centre of Baraka in South Kivu, Democratic Republic of Congo (DRC) | HemoCue 301<br><br>Baseline and follow-up clinic visits on day 1,2,3,7,14,21,28,35, and 42 | Baseline Hb:<br><br>ASAQ: 9.5 g/dl (IQR 8.5-10.8)<br><br>AL: 9.7 g/dl (IQR 8.7-10.8) |
| 11. Dhabangi *et al.*, 2015[13]<br><br>Protocol – supplement 1*<br><br>Tissue Oxygenation by Transfusion in severe Anaemia with Lactic acidosis (TOTAL) trial | RCT | To determine whether transfusion of RBCs stored for ≥25 days is not inferior to RBCs stored for ≤10 days for global tissue oxygenation as judged by the reduction of elevated blood lactate levels | 290 children aged 6-60 months with SA and lactic acidosis<br>(827 assessed)<br><br>Longer RBC storage (n=145)<br><br>Shorter RBC storage (n=145) | Paediatric Acute Care Unit of Mulago Hospital, Kampala, Uganda | HemoCue (capillary or venous) and I-STAT Hb (Chem8+ and EC8+ cartridges) (venous)<br><br>Hours 0,2,4,6,8 and 24 after transfusion | Baseline Mean±SD Hb:<br><br>Longer RBC storage: 3.7±1.3 g/dl<br><br>Shorter RBC storage: 3.6±1.3 g/dl |

| Study and publication/ registration year | Study design | Aim of study | Population and sample size (for those undergoing POC Hb testing if provided) | Location and setting | POC Hb test(s) used (blood sample and timing if provided) | Percentage of patients diagnosed with mild/moderate /severe/overall anaemia and/or mean Hb concentration by POC(Hb)T |
|---|---|---|---|---|---|---|
| 12. Gujo and Kare, 2021[14] | Cross-sectional | To assess the prevalence of intestinal parasite infection and its association with anaemia among children aged 6-59 months in Yirgalem General Hospital, Ethiopia | 367 children aged 6-59 months | Paediatric outpatient department of Yirgalem General Hospital, Southern Ethiopia | HemoCue 301 (capillary) | Mild (Hb 10-10.9g/dl): 24.5%<br><br>Moderate (Hb 7-9.9g/dl): 21.5%<br><br>Severe (Hb <7g/dl): 2.8%<br><br>Overall (Hb <11g/dl): 48.8% (95% CI: 43.7-53.9)<br><br>Mean±SD Hb: 11.46±1.95 g/dl |
| 13. Hawkes *et al.*, 2014[15] | Diagnostic study | To assess the agreement and diagnostic accuracy of POC devices for glucose, lactate, and haemoglobin in the management of severe malaria in a resource-constrained hospital in Uganda | 108 critically ill children aged 1-10 years with severe malaria | Regional Referral Hospital in Jinja, Uganda | I-STAT Hb (venous) (CHEM8+ cartridge) | Only provided for reference CBC:<br><br>Severe (<5g/dl): 21% |
| 14. Heckman *et al.*, 2020[16] | Cross-sectional | To investigate the prevalence of anaemia in children aged <5 years presenting for well-child examinations at a | 67 children <5 years<br><br>(52 clinically well) | Thohoyandou Health Centre (THC) in Thohoyandou, Limpopo | HemoCue 201+ (capillary) | Anaemia (Hb>2 SD below mean of age-specific, altitude adjusted values): 75% (out of 52) |

| Study and publication/ registration year | Study design | Aim of study | Population and sample size (for those undergoing POC Hb testing if provided) | Location and setting | POC Hb test(s) used (blood sample and timing if provided) | Percentage of patients diagnosed with mild/moderate /severe/overall anaemia and/or mean Hb concentration by POC(Hb)T |
|---|---|---|---|---|---|---|
| | | community health centre in South Africa | (71 children followed up) | Province, South Africa<br><br>Follow up: THC and Pfanani Clinic, Thohoyandou | | Median Hb: 9.65±2.6 g/dl<br><br>Follow-up:<br>Anaemia: 76% (out of 71) |
| 15. Kamugisha *et al.*, 2008[17] | Cross-sectional | To determine the level of malaria burden using Paracheck Pf® dipstick and microscopy among children clinically diagnosed with malaria and attending a healthcare facility in Tanga City, Tanzania | 301 children aged ≤10 years with suspected malaria | Outpatient department at Makorora Health Centre in Tanga City in north-eastern Tanzania | HemoCue (capillary) | Anaemia (Hb <11g/dl): 53.16%<br><br>Mean Hb: 10.92 g/dl (95% CI: 10.70-11.14)<br><br>Range: 4.70-16.0 g/dl |
| 16. Kebede *et al.*, 2021[18] | Cross-sectional | To determine the prevalence of anaemia and its associated factors among under-five children in Shanan Gibe Hospital, Southwest Ethiopia | 368 children <5years | Shanan Gibe Hospital, Jimma town, Southwest Ethiopia | HemoCue (capillary) | Anaemia (Hb <11g/dl): 48.9% (95% CI: 39.24-53.19)<br><br>Mild (Hb 10-10.9g/dl): 24.9%<br><br>Moderate (Hb 7.0-9.9g/dl): 15.8% |

| Study and publication/ registration year | Study design | Aim of study | Population and sample size (for those undergoing POC Hb testing if provided) | Location and setting | POC Hb test(s) used (blood sample and timing if provided) | Percentage of patients diagnosed with mild/moderate /severe/overall anaemia and/or mean Hb concentration by POC(Hb)T |
|---|---|---|---|---|---|---|
| | | | | | | Severe (Hb <7g/dl): 8.2% |
| 17. Keitel *et al.*, 2017[19]<br><br>NCT02225769 | RCT | 1)To determine whether a novel electronic algorithm using POC testing (e-POCT) was non-inferior in terms of clinical outcome to a validated electronic algorithm when managing febrile illness in children <5 years<br>2) To compare the proportion of antibiotic prescriptions and SAEs | 1586 children aged 2-59 months with acute febrile illness | Outpatient departments of 3 district hospitals and 6 health centres in Dar es Salaam, Tanzania | HemoCue 201+<br><br>When prompted by electronic algorithm (Admission, day 3, 7 or if concern) | Severe (Hb<6g/dl): 1.3%<br><br>Mean±SD: 9.7±1.5 g/dl |
| 18. Maiga *et al.*, 2020[20] | RCT | To understand the effects of longitudinal intermittent preventative treatment for malaria (sulphadoxine-pyrimethamine+ artesunate (SP+AS), amodiaquine +artesunate (AQ+AS)) among Malian schoolchildren | 296 school children aged 6-13 years | Study clinic in the rural village of Kolle, Mali (57km southwest of Bamako, capital city of Mali) | HemoCue (capillary)<br><br>Baseline and follow-up clinic visits (monthly) | Baseline Mean±SD Hb:<br>SP+AS: 11.4±1.4 g/dl<br>AQ+AS:11.3±1.5 g/dl<br>Control: 11.2±1.4 g/dl<br><br>Follow-up (2007):<br>SP+AS: 11.96g/dl (95% CI: 11.89-12.03)<br>AQ+AS: 11.91g/dl (95% CI: 11.98-12.05) |

| Study and publication/ registration year | Study design | Aim of study | Population and sample size (for those undergoing POC Hb testing if provided) | Location and setting | POC Hb test(s) used (blood sample and timing if provided) | Percentage of patients diagnosed with mild/moderate /severe/overall anaemia and/or mean Hb concentration by POC(Hb)T |
|---|---|---|---|---|---|---|
| | | | | | | Control: 11.60g/dl (95% CI: 11.53-11.67)<br><br>Follow up (2008):<br>SP+AS: 12.06 (95% CI: 11.98-12.15)<br>AQ+AS: 11.92g/dl (95% CI: 11.84-11.99)<br>Control: 11.64g/dl (95% CI: 11.56-11.72)<br><br>Follow-up (2009):<br>SP+AS: 12.63g/dl (95% CI: 12.27-12.52)<br>AQ+AS: 12.26g/dl (95% CI: 12.15-12.36)<br>Control: 12.16g/dl (95% CI: 12.05-12.27) |
| 19. Maitland *et al.*, 2019[21] (1)<br><br>Maitland *et al.*, 2019 (2)[22] | RCT | 1) To investigate immediate transfusion versus triggered transfusion in 1565 children with uncomplicated SA 2) To investigate whether a | 3983 children aged 2 months-12 years (1565 with Hb 4-6g/dl uncomplicated SA | Paediatric wards of 3 hospitals in Uganda: Mulago National Referral Hospital, Kampala, Mbale | HemoCue 301<br><br>(Screening, every 8 hours during first 24 hours, 48 hours or if triggered | Baseline (total):<br>Median Hb (IQR):<br>5.1 (4.6-5.7) g/dl (n=1565)[21]<br>4.2 (3.4-5.2) g/dl (n=3196)[22] (n=3188)[23] |

| **Study and publication/ registration year** | **Study design** | **Aim of study** | **Population and sample size (for those undergoing POC Hb testing if provided)** | **Location and setting** | **POC Hb test(s) used (blood sample and timing if provided)** | **Percentage of patients diagnosed with mild/moderate /severe/overall anaemia and/or mean Hb concentration by POC(Hb)T** |
|---|---|---|---|---|---|---|
| George *et al.,* 2022 (secondary analysis)[23]<br><br>ISRCTN84086586<br><br>The Transfusion and Treatment of Severe Anaemia in African Children Trial (TRACT) | | higher volume of whole blood with haemoglobin <6g/dl would improve outcomes compared with the standard volume in 3196 children (778 uncomplicated SA, 2418 complicated SA)<br><br>Secondary analysis:<br>To examine the safety and efficacy of different pack types on haematological correction, re-transfusion, mortality, and readmission to hospital | (2418 with Hb<4g/dl/ complicated SA)<br><br>(788 children included in both analyses) | Regional Referral Hospital, Soroti Regional Referral Hospital and one hospital in Malawi: Queen Elizabeth Central Hospital, Blantyre | by clinical deterioration and 28, 90, and 180 days post discharge | 48 hours (Mean±SD Hb)[21]:<br>Immediate transfusion: 8.4±1.7 g/dl<br>Triggered transfusion: 5.5±1.1 g/dl<br>Control: 7.0±1.7 g/dl |
| 20. Maitland *et al.*, 2011[24] (1)<br><br>Maitland *et al.,* 2013[25]<br><br>Kiguli *et al*., 2015 (2)[26]<br><br>ISRCTN69856593 | RCT | (1)To investigate the practice of early resuscitation with a saline bolus as compared with no bolus (control) and with an albumin bolus as compared with a saline bolus<br>(2) To present quality-controlled data on the | 3082 children aged 60 days -12 years with severe febrile illness<br><br>Exclusion: severe malnutrition, trauma, surgery burns etc | Paediatric wards at Kilifi District Hospital (Kenya), Teule District Hospital, Muheza, (Tanzania), Mulago National | HemoCue 301 (venous – admission)<br><br>(8, 24 and 48 hours post randomisation or if clinical deterioration) | Mild (Hb 7.0-10.0g/dl):<br>Admission: 27%<br>8 hours: 40%<br>24 hours: 45%<br><br>Moderate (Hb 5.0-7.0g/dl):<br>Admission: 16%<br>8 hours: 30% |

| Study and publication/ registration year | Study design | Aim of study | Population and sample size (for those undergoing POC Hb testing if provided) | Location and setting | POC Hb test(s) used (blood sample and timing if provided) | Percentage of patients diagnosed with mild/moderate /severe/overall anaemia and/or mean Hb concentration by POC(Hb)T |
|---|---|---|---|---|---|---|
| Fluid Expansion as Supportive Therapy (FEAST) Trial | | prevalence, clinical features, and transfusion management of anaemia in children presenting to hospitals in three East African countries with serious febrile illness and clinical signs of impaired peripheral perfusion | | Referral Hospital, Kampala, (Uganda), St Mary's Hospital, Lacor (Uganda), Soroti and Mbale Regional Referral Hospitals (Eastern Uganda) | | 24 hours: 29%<br><br>Severe (Hb<5.0g/dl): admission:33%<br>8 hours: 15%<br>24 hours: 9%<br><br>Total anaemic at admission: 76%<br><br>Mean±SD Hb: 7.1±3.2 g/dl[24] |
| 21. McGann *et al.*, 2015[27] | Diagnostic study | To evaluate the accuracy of a novel POC assay to detect moderate and severe anaemia in a limited-resource setting | 86 children with SCA | Outpatient Sickle Cell Clinic at Hospital Pediátrico David Bernardino in Luanda, Angola | Novel POC assay (capillary and venous) | Only provided for reference haematology analyser:<br><br>6.6±1.3 g/dl<br><br>Range: 2.8-9.3g/dl |
| 22. Mtove *et al.*, 2010[28] | Not reported | 1) To estimate the case fraction of invasive salmonellosis among paediatric admissions<br>2) To examine associations with common co- | 1478 children with fever aged 2 months-14 years | Teule Hospital, rural district hospital of Muheza in north-eastern Tanzania | HemoCue | Anaemia (<8g/dl): 45.6%<br><br>Severe (<5g/dl): 18.5%<br><br>Mean Hb: 8.0g/dl |

| **Study and publication/ registration year** | **Study design** | **Aim of study** | **Population and sample size (for those undergoing POC Hb testing if provided)** | **Location and setting** | **POC Hb test(s) used (blood sample and timing if provided)** | **Percentage of patients diagnosed with mild/moderate /severe/overall anaemia and/or mean Hb concentration by POC(Hb)T** |
|---|---|---|---|---|---|---|
| | | morbidities and describe its clinical features<br>3) To compare findings with those from previous studies among children in Sub-Saharan Africa | | | | |
| 23. Nass *et al.*, 2020[29]<br><br>NCT02941081 | Diagnostic study | To evaluate the accuracy of Aptus and HemoCue 301 against an automated haematology analyser in Gambian children aged 6-35 months and the Aptus' usage in the field | 223 children aged 6-35 months with iron deficiency anaemia | Rural clinics in Wuli and Sandu districts in the Bassé government area, Upper River Region, The Gambia | HemoCue 301 (venous and capillary) and Aptus (venous and capillary)<br><br>Capillary - weekly<br><br>Venous – day 85 | Mean±SD Hb:<br><br>Aptus (Capillary): 10.33±1.11 g/dl<br><br>HemoCue (capillary): 10.01±1.07 g/dl<br><br>Aptus (venous): 10.4±1.0 g/dl<br><br>HemoCue (venous): 10.6±0.9 g/dl |
| 24. Nkrumah *et al.*, 2011[30] | Diagnostic study | To generate data to support or refute the use of HemoCue POC device for Hb estimation in mobile blood donations and critical care areas in health facilities | 398 children and adults aged 1-64 years<br><br>(87 aged 1-4 years) | Agogo Presbyterian Hospital in the Asante Akim North District, Ashanti Region, Ghana | HemoCue B-haemoglobin (venous) | Mean±SD Hb:<br><br>Children aged 1-4 years: 9.3±2.9 g/dl<br>Range: 2.7-16.8 g/dl |

| Study and publication/ registration year | Study design | Aim of study | Population and sample size (for those undergoing POC Hb testing if provided) | Location and setting | POC Hb test(s) used (blood sample and timing if provided) | Percentage of patients diagnosed with mild/moderate /severe/overall anaemia and/or mean Hb concentration by POC(Hb)T |
|---|---|---|---|---|---|---|
| | | | (89 aged 5-14 years)<br>(222 aged >15 years) | | | Children aged 5-14 years: 9.8±3.1 g/dl<br>Range: 3.3-20.3 g/dl |
| 25. Ntonifor *et al.*, 2021[31] | Cross-sectional | To assess the implications of intestinal helminths on malaria clinical outcomes focusing on trends in malaria parasitaemia, anaemia and pyrexia among outpatients co-infected with malaria and intestinal helminths | 358 children and adults aged 3-62 years with fever and suspected malaria | Regional Hospital Bamenda and the Medicalised Health Centre Nkwen-Bamenda, Cameroon | URIT-12 (venous)<br><br>At enrolment | N/A - only reported for pooled children and adults sample |
| 26. Ocan *et al.*, 2018[32] | Cross-sectional | To determine the prevalence, severity, morphological characterisation, and the associated factors of anaemia among children <5 years at St. Mary's Hospital Lacor | 343 children <5 years | Paediatric outpatient department at St Mary's Hospital Lacor, Gulu District, Northern Uganda | HemoCue 201+ (capillary) | Anaemia (Hb <11g/dl): 46.6% (95% CI: 42.1-51.46)<br><br>Of those 46.6% anaemic were:<br><br>Mild (Hb 10-10.9g/dl): 29.4% |

| Study and publication/ registration year | Study design | Aim of study | Population and sample size (for those undergoing POC Hb testing if provided) | Location and setting | POC Hb test(s) used (blood sample and timing if provided) | Percentage of patients diagnosed with mild/moderate /severe/overall anaemia and/or mean Hb concentration by POC(Hb)T |
|---|---|---|---|---|---|---|
| | | | | | | Moderate (Hb 7-9.9g/dl): 58.8%<br><br>Severe (Hb <7g/dl): 11.9% |
| 27. Olatunya *et al.*, 2018[33] | Case-control | To determine the academic performance of children with SCA in comparison with their peers using school reports as surrogate | 101 children with steady state SCA aged 4-12 years<br><br>101 matched controls (non SCA) | Paediatric sickle cell clinic of the Ekiti State University Teaching Hospital, Ado Ekiti, Ekiti State, Nigeria | Mission Hb | Mean ±SD Hb:<br><br>SCA: 7.6±1.1 g/dl<br><br>Control: 10.57± 1.0 g/dl |
| 28. Olatunya *et al.*, 2016[34] | Diagnostic study | To examine the agreement between a POC device (Mission Hb) and haematology analyser for determining Hb concentration in children with SCD and its usefulness in resource-poor settings | 92 children aged 6-204 months (17 years) with SCD | Paediatric haematology unit of the Ekiti State University Teaching Hospital, Ado Ekiti, south-west Nigeria | Mission Hb (venous) | Severe (Hb<7g/dl): 20.7%<br><br>Mean±SD Hb: 7.69±1.6 g/dl<br><br>Range: 4.90-14.20g/dl |
| 29. Olupot-Olupot *et al.*, 2019[35] | Diagnostic study | To compare the accuracy of four methods of haemoglobin estimation that are commonly | 322 children aged 2 months-12 years with pallor | Paediatric wards of Mbale and Soroti Regional Referral | HemoCue 301 (capillary) and HCS | HemoCue 301:<br>Mild (Hb 10-11.9g/dl): 2.7% |

| Study and publication/ registration year | Study design | Aim of study | Population and sample size (for those undergoing POC Hb testing if provided) | Location and setting | POC Hb test(s) used (blood sample and timing if provided) | Percentage of patients diagnosed with mild/moderate /severe/overall anaemia and/or mean Hb concentration by POC(Hb)T |
|---|---|---|---|---|---|---|
| | | available in Uganda in terms of their ability to correctly identify children who are clinically severely anaemic and require blood transfusion | | Hospitals in Eastern Uganda | | Moderate (Hb 5-9.9g/dl): 45.2%<br><br>Severe (<5g/dl): 52.1% (Overall:100%)<br><br>Mean±SD Hb: 5.0±2.0 g/dl<br><br>Range: 0.7-11.8 g/dl<br><br>HCS:<br>Mild (Hb 10-11g/dl): 17.5%<br><br>Moderate (Hb 5-9.9g/dl): 49.4%<br><br>Severe (Hb<5g/dl): 22.8% (Overall: 89.7%) |
| 30. Onyangore *et al.*, 2016[36] | Cross-sectional | To assess the prevalence of iron deficiency anaemia and dietary iron intake among infants aged 6-9 | 244 infants 6-9 months | 3 health facilities in Metkei and Chepkorio | HemoControl (capillary) | Mild (10-10.9g/dl): 19.3%<br><br>Moderate (7-9.9 g/dl): 2.5% |

| Study and publication/ registration year | Study design | Aim of study | Population and sample size (for those undergoing POC Hb testing if provided) | Location and setting | POC Hb test(s) used (blood sample and timing if provided) | Percentage of patients diagnosed with mild/moderate /severe/overall anaemia and/or mean Hb concentration by POC(Hb)T |
|---|---|---|---|---|---|---|
| | | months in Keiyo South Sub County, Kenya | | divisions situated in the highlands of Keiyo South Sub County, Kenya: Kamwosor Sub County Hospital, Nyaru dispensary and Chepkorio Health Centre | | Severe (<7g/dl): 2.5%<br><br>Anaemia (Hb 10-10.9 and 7-9.9) = 21.7%<br>(Overall: 24.3%)<br><br>Mean±SD Hb: 11.3±0.84 g/dl<br><br>Range: 8.0-14.1 g/dl |
| 31. Parbey *et al.*, 2019[37] | Case-control | To determine the associated risk factors of anaemia among children <5 years in Hohoe Municipality | 210 children aged 6-59 months (70 anaemia cases, 140 controls) | Child Welfare Clinics in Hohoe Municipality, district of the Volta Region of Ghana | URIT-12 (capillary) | Anaemia (<11g/dl): 53.8% |
| 32. Parker *et al.*, 2018[38]<br><br>NCT0260350 | Diagnostic study | 1) To evaluate the accuracy of both the HemoCue 201+ wicking and gravity methodologies for measuring Hb levels 2) To evaluate the accuracy of the Pronto device for | 132 children aged 6-59 months<br><br>HemoCue wicking (n=66)<br>HemoCue gravity (n=66)<br>Pronto (n=112) | Centre Hospitlier Universitaire de Kigali in Rwanda | HemoCue 201+ gravity and wicking method (venous) and Pronto with DCI-mini sensors | HemoCue 201+ wicking:<br>Anaemia (Hb<11.5g/dl): 44%<br>Mean±SD Hb: 11.6±1.5 g/dl<br>Range: 7.9-16.1 g/dl<br><br>HemoCue 201+ gravity: |

| Study and publication/ registration year | Study design | Aim of study | Population and sample size (for those undergoing POC Hb testing if provided) | Location and setting | POC Hb test(s) used (blood sample and timing if provided) | Percentage of patients diagnosed with mild/moderate /severe/overall anaemia and/or mean Hb concentration by POC(Hb)T |
|---|---|---|---|---|---|---|
| | | measuring Hb levels among children in Rwanda | | | (thumb, ring finger, big toe) | Anaemia (Hb <11.5g/dl): 39%<br>Mean Hb: 11.8±1.7 g/dl<br>Range: 6.5-19.4 g/dl<br><br>Pronto:<br>Anaemia (<11.5g/dl): 42%<br>Mean Hb: 11.6±0.9 g/dl<br>Range: 9.7-14.3 g/dl |
| 33. Ramaswamy *et al.*, 2021[39] | Diagnostic study | To assess the validity of POC tests for estimation of haemoglobin among children aged 6-59 months in a facility setting compared with the gold standard haematology analyser | 120 children with minor ailments aged 6-59 months | Paediatric outpatient department clinics at Sub District Hospital, Ballabgarh in Haryana, India | HemoCue 301 (capillary), Rad-67 Pulse Co-Oximeter and rainbow DCI-mini sensor (thumb, finger, or toe) | HemoCue 301:<br>Anaemia: 73.3%<br>Mild: 28.4%<br>Moderate: 60.2%<br>Severe: 11.4%<br>Mean±SD Hb: 9.7±1.9 g/dl<br>Range: 4.4-13.5 g/dl<br><br>Rad-67:<br>Anaemia: 19.2%<br>Mild: 52.2%<br>Moderate:43.5%<br>Severe: 4.3% |

| Study and publication/ registration year | Study design | Aim of study | Population and sample size (for those undergoing POC Hb testing if provided) | Location and setting | POC Hb test(s) used (blood sample and timing if provided) | Percentage of patients diagnosed with mild/moderate /severe/overall anaemia and/or mean Hb concentration by POC(Hb)T |
|---|---|---|---|---|---|---|
| | | | | | | Mean±SD Hb: 11.9±1.5 g/dl<br>Range: 5.1-15.2g/dl<br><br>(Does not report threshold values) |
| 34. Silva *et al.*, 2021[40] | Cross-sectional | To evaluate the variability of children's haemoglobin levels from the Southern municipalities included in the Brazil Without Poverty Plan and their association with factors at the municipal and individual levels | 1,501 children aged 12-59 months | Basic Health Units in Paraná, Santa Catarina and Rio Grande do Sul states in Southern region of Brazil | HemoCue (venous) | Mean Hb:<br><br>12.8g/dl (95% CI: 12.7-12.8) |
| 35. Smart *et al.*, 2018[41]<br><br>Robust Assays for Point-of-care Identification of Disease (RAPID) study | Diagnostic study | To evaluate the diagnostic accuracy of a POC immunoassay (Sickle SCAN) to diagnose SCD and a first-generation POC color-based assay to detect anaemia compared to their respective gold standards in a real-world setting and to determine the | 429 children aged 1-20 years with suspected anaemia | Outpatient sickle cell clinic, emergency department, paediatric ward and paediatric outpatient clinic at Bugando Medical Centre in Mwanza, | Novel colour-based POC assay (capillary) | Only provided for haematology analyser:<br><br>Severe (Hb ≤7g/dl): 45%<br>Very severe (Hb≤5 g/dl): 20%<br><br>Mean±SD Hb: 7.3±2.7g/dl |

| Study and publication/ registration year | Study design | Aim of study | Population and sample size (for those undergoing POC Hb testing if provided) | Location and setting | POC Hb test(s) used (blood sample and timing if provided) | Percentage of patients diagnosed with mild/moderate /severe/overall anaemia and/or mean Hb concentration by POC(Hb)T |
|---|---|---|---|---|---|---|
| | | interobserver reliability between experienced and inexperienced end-users | | north-western Tanzania (Consultant and Teaching Hospital) | | |
| 36. Tack *et al.*, 2022[42]<br><br>NCT04473768<br><br>Clinical Decision Support in Non-typhoidal Salmonella (NTS) Bloodstream Infections in Children (DeNTs) study | Retrospective study (part of the DeNTs prospective cohort study) | To investigate obstacles experienced throughout the lifecycle of handheld diagnostic devices used for hospital triage of children <5 years presenting with severe febrile illness in a low-resource setting<br><br>DeNTS: To assess the contribution of handheld diagnostic instruments and POC tests to the detection of danger signs associated with bloodstream infections | 1880 children aged 28 days-5 years with severe febrile illness | Triage room of the Paediatric ward of St. Luc General Referral Hospital in Kisantu, Kongo Central Province, Democratic Republic of Congo (DRC) | HemoCue 801 (capillary) | Not reported |
| 37. Teshome *et al.*, 2017[43] | RCT | To assess non-inferiority of daily home fortification with 3mg iron as | 338 children aged 12-36 months (433 screened) | Research clinic in Kisumu West District, Kenya | HemoCue 301 (capillary) | 3mg NaFeEDTA:<br>Mild (Hb 10-19.9g/dl): 27.7% |

| Study and publication/ registration year | Study design | Aim of study | Population and sample size (for those undergoing POC Hb testing if provided) | Location and setting | POC Hb test(s) used (blood sample and timing if provided) | Percentage of patients diagnosed with mild/moderate /severe/overall anaemia and/or mean Hb concentration by POC(Hb)T |
|---|---|---|---|---|---|---|
| NCT02073149 | | NaFeEDTA compared with 12.5mg iron as encapsulated ferrous fumarate, with haemoglobin concentration after 30 days | 3mg NaFeEDTA (n=112)<br><br>12.5g ferrous fumarate (n=114)<br><br>Placebo (n=112) | | Baseline and follow-up clinic visits | Moderate (Hb 7-9.9g/dl): 30.4%<br>Mean±SD Hb: 10.59±1.33 g/dl<br><br>12.5mg ferrous fumarate:<br>Mild: 32.5%<br>Moderate: 31.6%<br>Mean±SD Hb: 10.47±1.33 g/dl<br><br>Placebo:<br>Mild:34.8%<br>Moderate:29.5%<br>Mean±SD Hb: 10.44±1.32 g/dl |
| 38. Ughasoro *et al.*, 2019[44] | Diagnostic study | To determine the discriminatory ability of Hb-Anae screening tool in detecting childhood anaemia when applied by the parents on their children in the community as well as determine the | 552 children <10 years | Health centres in Abakpa (urban) and Ibagwa (rural) in Enugu East Local Government Area of Enugu | HemoCue 301 (venous and capillary) | Anaemia (Hb<11g/dl): 55.1% |

| Study and publication/ registration year | Study design | Aim of study | Population and sample size (for those undergoing POC Hb testing if provided) | Location and setting | POC Hb test(s) used (blood sample and timing if provided) | Percentage of patients diagnosed with mild/moderate /severe/overall anaemia and/or mean Hb concentration by POC(Hb)T |
|---|---|---|---|---|---|---|
| | | sensitivity and specificity of the anaemia screening tool, compared with POC HemoCue haemoglobinometer | | State and Umuahia (urban) and Nkwegwu (rural) in Umuahia North Local Government Area in Abia State, southeast Nigeria | | |
| 39. Ughasoro *et al.*, 2019[45] | Diagnostic study | To evaluate the diagnostic accuracy of the HCS, among, children, implemented by different healthcare workers located in the urban and rural health facilities compared with the validated digital HemoCue analyser using standard protocol | 524 children <10 years | Health centres in Abakpa (urban) and Ibagwa (rural) in Enugu East Local Government Area of Enugu State and Umuahia (urban) and Nkwegwu (rural) in Umuahia North Local | HemoCue 301 (capillary) and HCS (capillary) | Anaemia (Hb<11g/dl):<br><br>Hemocue 301: 44.5%<br><br>HCS: 50% |

| **Study and publication/ registration year** | **Study design** | **Aim of study** | **Population and sample size (for those undergoing POC Hb testing if provided)** | **Location and setting** | **POC Hb test(s) used (blood sample and timing if provided)** | **Percentage of patients diagnosed with mild/moderate /severe/overall anaemia and/or mean Hb concentration by POC(Hb)T** |
|---|---|---|---|---|---|---|
| | | | | Government Area in Abia State, southeast Nigeria | | |
| 40. NCT05060198[46] (2021, ongoing-recruitment completed)<br><br>(CENTRAL* ID: CN-02331862) | RCT | To assess the therapeutic efficacy of Artemether-lumefantrine (AL) and Dihydroartemisinin-piperaquine (DP) for symptomatic, uncomplicated Plasmodium Falciparum infections among children based on parasitological, clinical and haematologic parameters | 350 children aged 6-59 months with fever and uncomplicated malaria | Outpatient clinics of hospitals and/or clinics in Western Kenya | HemoCue (capillary)<br><br>Baseline and follow-up clinic visits at days 7,14,28 and 42 | No results |
| 41. NCT00439595[47] (2007, ongoing – recruitment completed)<br><br>Diagnostic Methods to Improve Detection and Treatment of | Cluster RCT | To compare the effectiveness in basic health facilities of two simple diagnostic tools compared to control dispensaries in increasing rates of detection and treatment of anaemia | Children <5 years and pregnant women with suspected anaemia | Basic Health Facilities in Handeni District, Tanga, north-eastern Tanzania | HemoCue 201 and HCS | No results |

| Study and publication/ registration year | Study design | Aim of study | Population and sample size (for those undergoing POC Hb testing if provided) | Location and setting | POC Hb test(s) used (blood sample and timing if provided) | Percentage of patients diagnosed with mild/moderate /severe/overall anaemia and/or mean Hb concentration by POC(Hb)T |
|---|---|---|---|---|---|---|
| Anaemia in African Children (EARS) Trial | | | | | | |

POC(Hb)T = Point-of-care Haemoglobin Test. POC = Point-of care. Hb = Haemoglobin. HCS = Haemoglobin Colour Scale. SA = Severe anaemia. RCT= Randomised Controlled Trial. SD = Standard deviation. mg = milligram. CI = confidence Interval. CBC = Complete blood count. RBC = Red blood cell. SAE's = Severe adverse events. SCA = Sickle cell anaemia. SCD = Sickle cell disease. * Some information extracted from related/supporting documents.

**Supplementary Table S3: Data Extraction Table of Challenges Reported.**

| Study | POC Hb test(s) used | Challenges to POC Hb test use reported |
|---|---|---|
| 1. Aldridge *et a*l., 2012[4] | HCS | "Considerable inter-observer variability between HCS readers – lack of training (only 1 hour provided)"<br>"HCS overestimated Hb compared with HemoCue by 1.3/dl"<br>"LOA too wide to be clinically acceptable"<br>"Sensitivity and specificity clinically unacceptable" |
| 2. Bond *et al.*, 2014[6] | HemoSpec<br><br>HemoCue 201+ | Clinically unacceptable accuracy for HemoSpec:<br>"2g/dl accuracy is not sufficient for clinical use"<br>"While 2g/dl accuracy in the field represents better accuracy than other low-cost methods such as HCS, it is not sufficient for clinical use" |
| 3. Choukem *et al.*, 2020[8] | URIT-12<br><br>Mission Hb | "Both overestimated mean Hb values"<br><br>"Mission Hb failed to meet clinical accuracy requirements" |

| Study | POC Hb test(s) used | Challenges to POC Hb test use reported |
|---|---|---|
| 4. Hawkes *et al*., 2014[15] | I-STAT | "Sometimes a lack of cartridges or test strips at time of patient admission, repeated test failure due to elevated ambient temperature, problems with sample loading in cartridge, coagulation of whole-blood sample"<br><br>"Sources of variability include cartridge transportation and storage conditions, practitioner technique and testing environment"<br><br>"I-STAT Hb provided an unbiased estimate but LOAs indicate substantial variability in the measurement" |
| 5. McGann *et al*., 2015[27] | Novel POC colour-based assay | "Color scale used for result interpretation included intervals of approx. 1.0g/dl Hb, which made it difficult to estimate the value to 0.1 or 0.2g/dl precision"<br><br>Variability and subjectivity of result interpretation<br>"An objective reading device such as a smartphone app would improve the variability and reduce the subjectivity of result interpretation"<br>"For more reliable and broad implementation of this POC assay, a more precise means by which to determine the Hb concentration, such as an application software program with automatic scoring, will be necessary"<br><br>"POC assay is also dependent upon specific volumes of both assay reagents and added blood" – uses collection tubes in this study<br>"As the reaction is sensitive to the volume of blood collected, we utilised collection tubes of specific volumes and via capillary action" |
| 6. Nass *et al*., 2020[29] | Aptus and HemoCue 301 | Aptus:<br>"Centrifugation process is sensitive to both significant vibrations and spatial orientation, which led to instances in which a measurement was not initiated or aborted"<br>"Errors in sampling are only detected with photometry, which in majority of cases made it impossible to collect a second sample from the same participant – as already left clinic" |

| Study | POC Hb test(s) used | Challenges to POC Hb test use reported |
|---|---|---|
| | | "Errors in capillary blood sampling most commonly occurred due to underfilling of the cuvette – many children distressed by finger-prick, making it difficult to produce a droplet and appropriately fill the Aptus cuvette with the resulting blood film formed on the finger"<br><br>"Common error was overfilling cuvettes when venous blood was used on a plastic surface"<br>"Longer duration of measurement and difficult in handling the cuvettes"<br><br>"Both devices were shown to overestimate Hb concentration"<br>"Wide limits of agreement in Bland-Altman plots – exceeds clinically acceptable inaccuracy of ±1.0g/dl"<br>"Showed a relatively limited bias but large LOA"<br><br>Cost:<br>Hemocue 301: analyser £400, consumables £80 per 200 tests<br>Aptus: analyser £250, consumables £60 per 200 tests |
| 7. Nkrumah *et al.*, 2011[30] | HemoCue B-haemoglobin | "Presence of air bubbles, excess blood on the back of the microcuvettes, over filling of microcuvettes and insufficient mixing of the samples may lead to erroneous results" |
| 8. Olatunya *et al.*, 2016[34] | Mission Hb | "Requires proper maintenance for it to function optimally"<br>"Pass the device through quality control checks routinely" |
| 9. Olupot-Olupot et al., 2019[35] | HCS, HemoCue 301 | HemoCue 301:<br>"Initial cost between 250-350 USD for the analyser (approx. 4 USD per test) – more expensive than Sahli's method (initial 40-50 USD and 0.25 USD per test)"<br><br>"Data suggests that in situations when HemoCue is unaffordable…." |
| 10. Parker *et al.*, 2018[38] | HemoCue 201+ wicking and gravity method, Pronto DCI-mini | "HemoCue wicking method and Pronto device were found to overestimate Hb concentration among anaemic children and underestimate Hb concentration among healthy children"<br>"Possible user variability and blood sampling techniques of HemoCue wicking method"<br>"HemoCue 201+ relatively high cost and limited availability of the disposable cuvettes" |

| Study | POC Hb test(s) used | Challenges to POC Hb test use reported |
|---|---|---|
| | | "Effects of user variability and environmental factors such as heat and humidity"<br>"Accuracy of Pronto is not clinically acceptable"<br>"Results show a detection failure rate of 15%" (Pronto) |
| 11. Ramaswamy *et al.*, 2021[39] | HemoCue 301 and Rad-67 Spot check pulse co oximeter | "Several factors affect accuracy including geographic variations (altitude and temperature), procedural differences in capillary blood sample collection (intra and inter observer variations), study participant related factors (e.g. age, sex, biological determinants)"<br><br>Rad-67:<br>"Performance dependent on signal quality indicators such as motion, perfusion index quality and ambient light interference"<br>"Was difficult to obtain stable indicators amongst children with continuous body movement"<br>"Showed poor concordance with haematology analyser"<br>"HemoCue and Rad-67 overestimated Hb value"<br><br>Cost:<br>HemoCue 301: "286 USD – approx. 0.41 USD per test"<br>Rad-67: "1770 USD – approx. 0.68 USD per test – relatively high"<br>Sahli's: "13 USD – approx. 0.20 USD per test" |
| 12. Smart *et al.*, 2017[41] | Novel POC colour-based assay | "Challenge of reagent instability resulting in color drift away from the pre-printed color scale"<br>"Only measures a maximum Hb concentration of 9.1g/dl" |
| 13. Tack *et al.*, 2022[42] | HemoCue 801 | "Pronto/Rad-67 not ordered due to high cost (USD 1-2 per test)"<br><br>"Difficulties in capillary blood sampling and transfer to microcuvettes caused unreliable Hb measurements at the start of the study"<br>"Poor repeatability of Hb measurements (up to 2g/dl differences) was linked with subtle underfilling of the microcuvette and 'milking' of the finger during capillary blood sampling"<br>"Device implementation required high investments (standard operating procedures, hands-on training workshops, supervised rotation, 'device experts'" |

| Study | POC Hb test(s) used | Challenges to POC Hb test use reported |
|---|---|---|
| | | "Order-of-draw of successive tests performed on capillary blood (order switched to take HemoCue sample before malaria rapid test to ensure 10µl aspiration – 3rd drop not always sufficient)"<br><br>"A consistent point impacting affordability, delivery and access was the lack of an in-country distributor, which required logistics (stock management, international shipments), impacted work (calibration) and entailed additional costs"<br><br>"Single-use consumables (microcuvettes) requiring disposal"<br><br>"Difficult to fill microcuvette correctly/completely – if child moves, if blood drop is small, if blood drop is poorly delineated (particularly in severely anaemic children) – agitation complicated measurements by capillary blood sampling"<br>"Massing finger or strong pressure on finger can disturb result"<br><br>Maintenance:<br>"Cleaning is a bit delicate and complicated: concentration needed to clean the interior part without leaving cotton particles – wait until all parts are dry before reassembling – sometimes difficult to reinsert the microcuvette support"<br>"QC fluid is expensive and difficult to get and store due to cold chain requirements" |

POC = Point-of care. Hb = Haemoglobin. HCS = Haemoglobin Colour Scale. LOA = Limits of Agreement. USD = United States Dollar. QC = Quality Control. g/dl = grams per decilitre.

**Supplementary Table S4: Adapted QUADAS-2 Tool[48]**

**Phase 1: State the review question:**

| |
|---|
| *Patients (setting, intended use of index test, presentation, prior testing):*<br><br>Any children aged <21 years attending health facilities in LMICs (clinics, hospitals, basic health units, medical practices etc.) for a POC Hb test to diagnose anaemia |
| *Index test(s):*<br><br>Any POC(Hb)T (providing immediate results at site of care) |
| *Reference standard and target condition:*<br><br>Any reference test used (other POC(Hb)T or non POC(Hb)T e.g., automated haematology analyser)<br><br>Target condition: anaemia (all cause and severity) |

**Phase 2: Draw a flow diagram for the primary study**

**Phase 3: Risk of bias and applicability judgements**

Signalling questions were added to aid judgement of RoB for domain two: "Were methods for test conduct clearly described?" and domain three: "Was HemoCue used as reference test?". The signalling question "Were the index test results interpreted without knowledge of results of reference standard?" was omitted for objective index tests.

**Domain 1: PATIENT SELECTION**

**A: Risk of Bias**

- Was a consecutive or random sample of patients enrolled? Yes/No/Unclear
- Was a case-control design avoided? Yes/No/Unclear
- Did the study avoid inappropriate exclusions? Yes/No/Unclear

Could the selection of patients have introduced bias? **RISK: LOW/HIGH/UNCLEAR**

**B: Concerns regarding applicability**

Is there concern that the included patients do not match the review question? **CONCERN: LOW/HIGH/UNCLEAR**

**Domain 2: INDEX TEST(S)**

If more than one index test was used, please complete for each test.

**A: Risk of Bias**

- Were the index test results interpreted without knowledge of the results of the reference standard? (omitted for objective index tests) Yes/No/Unclear
- If a threshold was used, was it reported or pre-specified? Yes/No/Unclear
- Were methods for test conduct clearly described? Yes/No/Unclear

Could the conduct or interpretation of the index test have introduced bias? **RISK: LOW/HIGH/UNCLEAR**

**B: Concerns regarding applicability**

Is there concern that the index test, its conduct, or interpretation differ from the review question? **CONCERN: LOW/HIGH/UNCLEAR**

**Domain 3: RERERENCE STANDARD**

**A: Risk of Bias**

- Is the reference standard likely to correctly classify the target condition? Yes/No/Unclear
- Was HemoCue used as reference test? Yes/No/Unclear
- Were the reference standard results interpreted without knowledge of the results of the index test? Yes/No/Unclear

Could the reference standard, its conduct, or its interpretation have introduced bias? **RISK: LOW/HIGH/UNCLEAR**

**B: Concerns regarding applicability**

Is there concern that the target condition as defined by the reference standard does not match the review question? **CONCERN: LOW/HIGH/UNCLEAR**

**Domain 4: FLOW AND TIMING**

**A: Risk of Bias**

- Was there an appropriate interval between index test(s) and reference standard? Yes/No/Unclear
- Did all patients receive a reference standard? Yes/No/Unclear
- Did patients receive the same reference standard? Yes/No/Unclear
- Were all patients included in the analysis? Yes/No/Unclear

Could the patient flow have introduced bias? **RISK: LOW/HIGH/UNCLEAR**

**Supplementary Table S5: Blood Sample, Volume and Result Time Required for included POC(Hb)Ts**

| POC Hb test | Blood sample and volume | Result time |
|---|---|---|
| **Invasive** | | |
| **HemoCue 301** (Hemocue AB, Ängelholm, Sweden) | 10µl<br>capillary, venous or arterial | ≤3 secs |
| **HemoCue 201+**<br>(Hemocue AB, Ängelholm, Sweden) | 10µl<br>capillary, venous or arterial | 15-60 secs |
| **HCS** (COPACK, Oststeinbek, Germany) | Drop of capillary or venous blood | 30 secs |
| **Mission Hb** (ACON Laboratories, Inc., San Diego, USA) | 10µl capillary or venous | ≤15 secs |
| **URIT-12** (URIT Medical Electronics, Guangxi, China) | 13-15µl capillary or venous | ≤10 secs |
| **I-STAT** (Abbott, Abbott Park, IL) | 65µl capillary, venous or arterial (EC8+)<br><br>95µl venous or arterial (CHEM8+) | 120 secs |
| **HemoCue 201** (Hemocue AB, Ängelholm, Sweden) | 10µl<br>capillary, venous or arterial | 15-60 secs |
| **Novel colour-based assay** | 10µl capillary or venous | 60 secs |
| **HemoCue 801** (Hemocue AB, Ängelholm, Sweden) | 10µl<br>capillary, venous or arterial | ≤1 sec |
| **HemoCue B-haemoglobin**<br>(Hemocue AB, Ängelholm, Sweden) | 10µl<br>capillary, venous or arterial | 30-50 secs |
| **Aptus** (Entia, London, UK) | 4-8µl<br>Capillary or venous | ≤60 secs |
| **HemoSpec** | 10µl whole blood | Approx. 20 secs |
| **HemoControl**<br>(EKF Diagnostics, Cardiff, UK) | 8µl capillary, venous or arterial | 25-60 secs |

| POC Hb test | Blood sample and volume | Result time |
|---|---|---|
| **Non-invasive** | | |
| **Rad-67™ Pulse CO-Oximeter® and rainbow® DCI®-mini-Sensor** (Masimo Corporation, Irvine, USA) | Not required | 30 secs |
| **Pronto® device with DCI-mini™ sensors** (Masimo Corporation, Irvine, USA) | Not required | 40 secs |

Hb = Haemoglobin. IL = Illinois. UK = United Kingdom. USA = United States of America. nm = Nanometre. μl = Microlitre. Secs = Seconds.

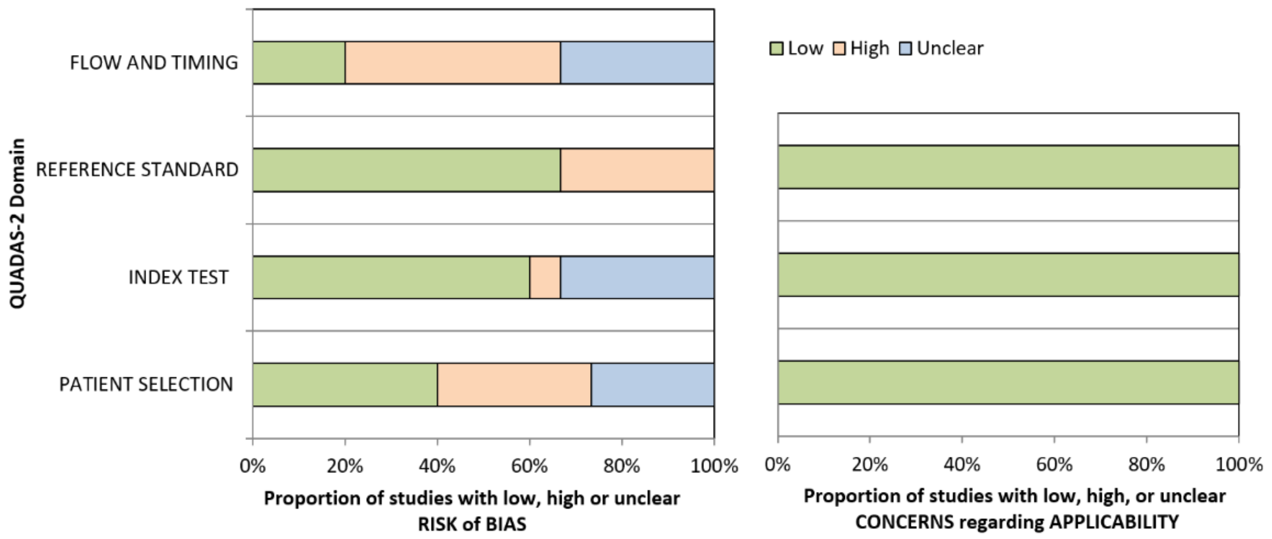

**Supplementary Figure S1. QUADAS-2 Risk of Bias and Applicability Concerns for Included Diagnostic Studies**

Presented as percentage of studies with low, high, or unclear risk of bias and applicability concerns for each domain. Where studies tested two or more point-of-care haemoglobin tests (index tests), separate judgements were made for each domain.

**Supplementary Table S6: Justifications for Quality Assessment Judgements.**

| Diagnostic study | Domain 1: Patient Selection | | Domain 2: Index Test(s) | | Domain 3: Reference Standard | | Domain 4: Flow and Timing |
|---|---|---|---|---|---|---|---|
| | A: Risk of Bias | B: Concern | A: Risk of Bias | B: Concern | A: Risk of Bias | B: Concern | A: Risk of Bias |
| Aldridge *et al.*, 2012[4] | Low ☺ | Low ☺ | Low ☺ | Low ☺ | High ☹ | Low ☺ | High ☹ |
| | Consecutive sample, avoided case-control design and inappropriate exclusions (no refusals) | Patients and setting match review question | HCS performed before/without knowledge of reference standard. Training provided and threshold pre-specified (Hb<11g/dl and SA Hb<5g/dl). Methods for test conduct well described | POC Hb test, conduct, and interpretation match review question | HemoCue used as reference method. Literature is limited and variable on test accuracy in this population and setting | Target condition (anaemia) defined by reference test matches review question | 11 patients excluded from final analysis due to incomplete set of data |
| Bond *et al.*, 2014[6] | Unclear ? | Low ☺ | Low ☺ | Low ☺ | High ☹ | Low ☺ | High ☹ |
| | Unclear if a consecutive or random sample was enrolled | Patients and setting match review question | Methods for test conduct described. Threshold for anaemia not pre-specified but sensitivity/specificity data not reported (and therefore not applicable) | POC Hb test, conduct, and interpretation match review question | HemoCue 201+ used as reference method. Literature is limited and variable on test accuracy in this population and setting | Target condition (anaemia) defined by reference test matches review question | 19 patients excluded from final analysis |

| **Diagnostic study** | **Domain 1: Patient Selection** | | **Domain 2: Index Test(s)** | | **Domain 3: Reference Standard** | | **Domain 4: Flow and Timing** |
|---|---|---|---|---|---|---|---|
| | **A: Risk of Bias** | **B: Concern** | **A: Risk of Bias** | **B: Concern** | **A: Risk of Bias** | **B: Concern** | **A: Risk of Bias** |
| Hawkes *et al.*, 2014[15] | Unclear ? | Low ☺ | Low ☺ | Low ☺ | Low ☺ | Low ☺ | High ☹ |
| | Unclear if a consecutive or random sample was enrolled | Patients and setting match review question | Routine QC, methods described and acceptable threshold for SA used (Hb <5g/dl) | POC Hb test, conduct, and interpretation match review question | Reference test likely to correctly classify anaemia (laboratory haematology analyser) | Target condition (anaemia) defined by reference test matches review question | 72 patients missing from final analysis. Reference sample sent to laboratory within 24 hours – may alter sample if not stored properly etc. |
| McGann *et al.*, 2015[27] | Unclear ? | Low ☺ | Low ☺ | Low ☺ | Low ☺ | Low ☺ | Unclear ? |
| | Unclear if a consecutive or random sample was enrolled or if inappropriate exclusions were avoided | Patients and setting match review question | A single blinded reader assessed all samples – minimises bias in colour-interpretation. Acceptable threshold for SA used (Hb <7g/dl and <5g/dl for very SA). Methods for test conduct well described | POC assay, conduct, and interpretation match review question | Reference test likely to correctly classify anaemia (laboratory haematology analyser) | Target condition (anaemia) defined by reference test matches review question | Does not report total number of children enrolled (pilot study) or time interval between samples for index test and reference standard |

| **Diagnostic study** | **Domain 1: Patient Selection** | | **Domain 2: Index Test(s)** | | **Domain 3: Reference Standard** | | **Domain 4: Flow and Timing** |
|---|---|---|---|---|---|---|---|
| | **A: Risk of Bias** | **B: Concern** | **A: Risk of Bias** | **B: Concern** | **A: Risk of Bias** | **B: Concern** | **A: Risk of Bias** |
| Nass *et al.*, 2020[29] (HemoCue 301 venous vs. haematology analyser) | High ☹ | Low ☺ | Low ☺ | Low ☺ | Low ☺ | Low ☺ | Unclear ? |
| | Random sample but exclusions included SA, malaria and SCD that could lead to underestimation/ overestimation of diagnostic accuracy | Patients and setting match review question | Venous blood collected following standardised protocols, internal QC. Threshold for anaemia not pre-specified but sensitivity/specificity data not reported (and therefore not applicable). Methods for test conduct well described. | POC Hb test, conduct, and interpretation match review question | Reference test likely to correctly classify anaemia (laboratory haematology analyser) | Target condition (anaemia) defined by reference test matches review question | Unclear if all patients recruited were included in final analysis of 223 children |
| Nass *et al.*, 2020[29] (Aptus venous vs. Haematology analyser) | High ☹ | Low ☺ | Low ☺ | Low ☺ | Low ☺ | Low ☺ | Unclear ? |
| | Random sample but exclusions included SA, | Patients and setting match review question | Venous blood collected following standardised | POC Hb test, conduct, and interpretation | Reference test likely to correctly | Target condition (anaemia) | Unclear if all patients recruited were included in final |

| **Diagnostic study** | **Domain 1: Patient Selection** | | **Domain 2: Index Test(s)** | | **Domain 3: Reference Standard** | | **Domain 4: Flow and Timing** |
|---|---|---|---|---|---|---|---|
| | **A: Risk of Bias** | **B: Concern** | **A: Risk of Bias** | **B: Concern** | **A: Risk of Bias** | **B: Concern** | **A: Risk of Bias** |
| | malaria and SCD that could lead to underestimation/ overestimation of diagnostic accuracy | | protocols, internal QC, error messages reduce bias in sampling technique. Threshold for anaemia not pre-specified but sensitivity/specificity data not reported (and therefore not applicable). Methods for test conduct well described | match review question | classify anaemia (laboratory haematology analyser) | defined by reference test matches review question | analysis of 223 children |
| Nass *et al.*, 2020[29] (Aptus venous vs. HemoCue 301 venous) | High ☹ | Low ☺ | Low ☺ | Low ☺ | High ☹ | Low ☺ | Unclear ? |
| | Random sample but exclusions included SA, malaria and SCD that could lead to underestimation/ | Patients and setting match review question | Venous blood collected following standardised protocols, internal QC, error messages reduce bias in sampling technique. Threshold for anaemia not pre- | POC Hb test, conduct, and interpretation match review question | HemoCue 301 used as comparator method. Literature is limited and variable on test accuracy in this | Target condition (anaemia) defined by reference test matches review question | Unclear if all patients recruited were included in final analysis of 223 children |

| **Diagnostic study** | **Domain 1: Patient Selection** | | **Domain 2: Index Test(s)** | | **Domain 3: Reference Standard** | | **Domain 4: Flow and Timing** |
|---|---|---|---|---|---|---|---|
| | **A: Risk of Bias** | **B: Concern** | **A: Risk of Bias** | **B: Concern** | **A: Risk of Bias** | **B: Concern** | **A: Risk of Bias** |
| | overestimation of diagnostic accuracy | | specified but sensitivity/specificity data not reported (and therefore not applicable). Methods for test conduct well described | | population and setting | | |
| Nass *et al.*, 2020[29] (Aptus capillary vs. HemoCue 301 capillary) | High ☹ | Low ☺ | High ☹ | Low ☺ | High ☹ | Low ☺ | Unclear ? |
| | Random sample but exclusions included SA, malaria and SCD that could lead to underestimation/ overestimation of diagnostic accuracy | Patients and setting match review question | Limitations reported of techniques used and could introduce bias. For example, use of thumb instead of second or third finger for capillary sample and squeezing of finger – did not follow manufacturer's instructions | POC Hb test, conduct, and interpretation match review question | HemoCue 301 used as comparator method. Literature is limited and variable on test accuracy in this population and setting | Target condition (anaemia) defined by reference test matches review question | Unclear if all patients recruited were included in final analysis of 223 children |

| **Diagnostic study** | **Domain 1: Patient Selection** | | **Domain 2: Index Test(s)** | | **Domain 3: Reference Standard** | | **Domain 4: Flow and Timing** |
|---|---|---|---|---|---|---|---|
| | **A: Risk of Bias** | **B: Concern** | **A: Risk of Bias** | **B: Concern** | **A: Risk of Bias** | **B: Concern** | **A: Risk of Bias** |
| Olatunya *et al.*, 2016[34] | Low ☺ | Low ☺ | Low ☺ | Low ☺ | Low ☺ | Low ☺ | Unclear ? |
| | Consecutive sample used and avoided case-control design | Patients and setting match review question | Training provided on device according to manufacturer's instructions, good QC, and acceptable threshold for SA pre-specified (Hb<7g/dl). Methods for test conduct described. | POC Hb test, conduct, and interpretation match review question | Reference test likely to correctly classify anaemia (laboratory haematology analyser) | Target condition (anaemia) defined by reference test matches review question | Does not report if all patients tested were included in final analysis |
| Olupot-Olupot *et al.*, 2019[35] | Low ☺ | Low ☺ | Unclear ? | Low ☺ | High ☹ | Low ☺ | High ☹ |
| | Consecutive sample, avoided case-control design and inappropriate exclusions | Patients and setting match review question | Unclear if HCS conducted before/without knowledge of HemoCue result | POC Hb test, conduct, and interpretation match review question | HemoCue 301 used as reference method. Literature is limited and variable on test accuracy in this population and setting | Target condition (anaemia) defined by reference test matches review question | Not all patients included in final analysis and no explanation for this. 322 participants enrolled, 263 and 292 included in HCS and reference analyses, respectively |

| **Diagnostic study** | **Domain 1: Patient Selection** | | **Domain 2: Index Test(s)** | | **Domain 3: Reference Standard** | | **Domain 4: Flow and Timing** |
|---|---|---|---|---|---|---|---|
| | **A: Risk of Bias** | **B: Concern** | **A: Risk of Bias** | **B: Concern** | **A: Risk of Bias** | **B: Concern** | **A: Risk of Bias** |
| Parker *et al.*, 2018[38] (HemoCue 201+) | Low ☺ | Low ☺ | Unclear ? | Low ☺ | Low ☺ | Low ☺ | Low ☺ |
| | Random sample, avoided case-control design and inappropriate exclusions | Patients and setting match review question | Threshold for anaemia (Hb<11.5g/dl) pre-specified but little information provided on test conduct | POC Hb test, conduct, and interpretation match review question | Reference test likely to correctly classify anaemia (laboratory haematology analyser) | Target condition (anaemia) defined by reference test matches review question | All patients included in final analysis for HemoCue, samples taken at same time and received same reference standard |
| Parker *et al.*, 2018[38] (Pronto) | Low ☺ | Low ☺ | Unclear ? | Low ☺ | Low ☺ | Low ☺ | High ☹ |
| | Random sample, avoided case-control design and inappropriate exclusions | Patients and setting match review question | Threshold for anaemia (Hb<11.5g/dl) pre-specified but little information provided on test conduct | POC Hb test, conduct, and interpretation match review question | Reference test likely to correctly classify anaemia (laboratory haematology analyser) | Target condition (anaemia) defined by reference test matches review question | Only 112 of 132 children completed Pronto measurements and included in final analysis – could be due to patient factors such as $SpO_2$ resulting in detection failure and introducing bias |

| Diagnostic study | Domain 1: Patient Selection | | Domain 2: Index Test(s) | | Domain 3: Reference Standard | | Domain 4: Flow and Timing |
|---|---|---|---|---|---|---|---|
| | A: Risk of Bias | B: Concern | A: Risk of Bias | B: Concern | A: Risk of Bias | B: Concern | A: Risk of Bias |
| Ramaswamy *et al.*, 2021[39] (HemoCue 301) | High ☹ | Low ☺ | Unclear ? | Low ☺ | Low ☺ | Low ☺ | Low ☺ |
| | Consecutive sample but severely ill children were excluded – could introduce bias and lead to overestimation/ underestimation of diagnostic accuracy | Patients and setting match review question | Threshold used for anaemia diagnosis not reported – could introduce bias if non-acceptable threshold used<br><br>(Methods for test conduct described) | POC Hb tests, conduct, and interpretation match review question | Reference test likely to correctly classify anaemia (laboratory haematology analyser) | Target condition (anaemia) defined by reference test matches review question | All patients enrolled were included in final analysis and received same reference standard. Blood samples were collected at same time with no interventions between tests |
| Ramaswamy *et al.*, 2021[39] (Rad-67) | High ☹ | Low ☺ | Unclear ? | Low ☺ | Low ☺ | Low ☺ | Low ☺ |
| | Consecutive sample but severely ill children were excluded – could introduce bias and lead to overestimation/ underestimation of diagnostic accuracy | Patients and setting match review question | Threshold used for anaemia diagnosis not reported – could introduce bias if non-acceptable threshold used.<br><br>(Methods for test conduct described) | POC Hb test, conduct, and interpretation match review question | Reference test likely to correctly classify anaemia (laboratory haematology analyser) | Target condition (anaemia) defined by reference test matches review question | All patients enrolled were included in final analysis and received same reference standard |

| **Diagnostic study** | **Domain 1: Patient Selection** | | **Domain 2: Index Test(s)** | | **Domain 3: Reference Standard** | | **Domain 4: Flow and Timing** |
|---|---|---|---|---|---|---|---|
| | **A: Risk of Bias** | **B: Concern** | **A: Risk of Bias** | **B: Concern** | **A: Risk of Bias** | **B: Concern** | **A: Risk of Bias** |
| Smart *et al.*, 2017[41] | Unclear ? | Low ☺ | Low ☺ | Low ☺ | Low ☺ | Low ☺ | High ☹ |
| | Unclear if a consecutive or random sample was enrolled or if inappropriate exclusions were avoided | Patients and setting match review question | Observers were blinded and training provided (although minimal). Acceptable threshold for SA used (Hb≤7g/dl and very SA Hb≤5g/dl). Methods for test conduct well described. | POC assay, conduct, and interpretation match review question | Reference test likely to correctly classify anaemia (laboratory haematology analyser) | Target condition (anaemia) defined by reference test matches review question | Not all patients included in final analysis and not all patients received a reference standard |
| Ughasoro *et al.*, 2019[45] | Low ☺ | Low ☺ | Low ☺ | Low ☺ | High ☹ | Low ☺ | High ☹ |
| | Random sample, case-control design avoided and no exclusion criteria – randomly selected households with children <10 years | Patients and setting match review question – selected households invited to health facility for Hb measurement | Training was provided on use of HCS, results were interpreted independently/before reference method and threshold was pre-specified (Hb<11g/dl). Methods for test conduct well described. | POC Hb test, conduct, and interpretation match review question | HemoCue 301 used as reference method. Literature is limited and variable on test accuracy in this population and setting | Target condition (anaemia) defined by reference test matches review question | 573 children enrolled but only 524 included in final analysis and no explanation for this. Exact time interval between index and reference test not reported |

POC = Point-of care. HCS = Haemoglobin Colour Scale. Hb = Haemoglobin. SA = Severe anaemia. QC = Quality control. SCD = Sickle cell disease. $SpO_2$ = Oxygen saturation.

### References

1. Cochrane Effective Practice and Organisation of Care (EPOC) LMIC Filters. 2020 [Available from: https://epoc.cochrane.org/lmic-filters. [Accessed 20 April 2022].
2. World Bank Country and Lending Groups 2022 [Available from: https://datahelpdesk.worldbank.org/knowledgebase/articles/906519-world-bank-country-and-lending-groups. [Accessed 20 April 2022].
3. Alamneh YM, Akalu TY, Shiferaw AA, Atnaf A. Magnitude of anemia and associated factors among children aged 6-59 months at Debre Markos referral hospital, Northwest Ethiopia: a hospital-based cross-sectional study. *Italian journal of pediatrics*. 2021;**47**(1):172.
4. Aldridge C, Foster HME, Albonico M, Ame SM, Montresor A. Evaluation of the diagnostic accuracy of the Haemoglobin Colour Scale to detect anaemia in young children attending primary healthcare clinics in Zanzibar. *Tropical Medicine and International Health.* 2012;**17**(4):423-9.
5. Bojang KA, Akor F, Conteh L, Webb E, Bittaye O, Conway DJ, *et al*. Two strategies for the delivery of IPTc in an area of seasonal malaria transmission in the Gambia: A randomised controlled trial. *PLoS Medicine.* 2011;**8**(2):e1000409.
6. Bond M, Mvula J, Molyneux E, Richards-Kortum R. Design and Performance of a Low-Cost, Handheld Reader for Diagnosing Anemia in Blantyre, Malawi. *Health Innov Point Care Conf.* 2014;**2014**:267-70.
7. Calis JC, Phiri KS, Faragher EB, Brabin BJ, Bates I, Cuevas LE, *et al*. Severe anemia in Malawian children. *Malawi Med J.* 2016;**28**(3):99-107.
8. Choukem S-P, Sih C, Ntumsi AT, Dimala CA, Mboue-Djieka Y, Ngouadjeu EDT, *et al*. Evaluation of the accuracy of two point-of-care haemoglobin meters used in sub-Saharan African population: a cross-sectional study. *BMC cardiovascular disorders.* 2020;**20**(1):111.
9. Cusick SE, Opoka RO, Ssemata AS, Georgieff MK, John CC. Comparison of iron status 28 d after provision of antimalarial treatment with iron therapy compared with antimalarial treatment alone in Ugandan children with severe malaria. *American Journal of Clinical Nutrition.* 2016;**103**(3):919-25.
10. Cusick SE, Opoka RO, Abrams SA, John CC, Georgieff MK, Mupere E. Delaying Iron therapy until 28 days after antimalarial treatment is associated with greater Iron incorporation and equivalent hematologic recovery after 56 days in children: A randomized controlled trial. *Journal of Nutrition.* 2016;**146**(9):1769-74.
11. Degarege A, Animut A, Medhin G, Legesse M, Erko B. The association between multiple intestinal helminth infections and blood group, anaemia and nutritional status in human populations from Dore Bafeno, southern Ethiopia. *Journal of Helminthology.* 2014;**88**(2):152-9.
12. de Wit M, Funk AL, Moussally K, Nkuba DA, Siddiqui R, Bil K, *et al*. In vivo efficacy of artesunate-amodiaquine and artemether-lumefantrine for the treatment of uncomplicated falciparum malaria: an open-randomized, non-inferiority clinical trial in South Kivu, Democratic Republic of Congo. *Malar J.* 2016;**15**(1):455.
13. Dhabangi A, Ainomugisha B, Cserti-Gazdewich CM, Ddungu H, Kyeyune D, Musisi E, *et al.* Tissue oxygenation by transfusion in severe anemia with lactic acidosis (total): A prospective, randomized, non-inferiority trial of blood storage duration. *Blood*. 2015;**126**(23):769.
14. Gujo AB, Kare AP. Prevalence of Intestinal Parasite Infection and its Association with Anemia among Children Aged 6 to 59 Months in Sidama National Regional State, Southern Ethiopia. *Clinical medicine insights Pediatrics*. 2021;**15**:11795565211029259.
15. Hawkes M, Conroy AL, Opoka RO, Namasopo S, Liles WC, John CC*, et al*. Performance of point-of-care diagnostics for glucose, lactate, and hemoglobin in the management of severe malaria in a resource-constrained hospital in Uganda. *The American journal of tropical medicine and hygiene. 2014;**90**(4):605-8.*

16. Heckman J, Samie A, Bessong P, Ntsieni M, Hamandi H, Kohler M, *et al.* Anaemia among clinically well under-fives attending a community health centre in Venda, Limpopo Province. *South African medical journal = Suid-Afrikaanse tydskrif vir geneeskunde.* 2010;**100**(7):445-8.
17. Kamugisha ML, Msangeni H, Beale E, Malecele EK, Akida J, Ishengoma DR, *et al.* Paracheck PfR compared with microscopy for diagnosis of Plasmodium falciparum malaria among children in Tanga City, north-eastern Tanzania. *Tanzania Journal of Health Research.* 2008;**10**(1):14-9.
18. Kebede D, Getaneh F, Endalamaw K, Belay T, Fenta A. Prevalence of anemia and its associated factors among under-five age children in Shanan gibe hospital, Southwest Ethiopia. *BMC pediatrics.* 2021;**21**(1):542.
19. Keitel K, Kagoro F, Samaka J, Masimba J, Said Z, Temba H, *et al.* A novel electronic algorithm using host biomarker point-of-care tests for the management of febrile illnesses in Tanzanian children (e-POCT): A randomized, controlled non-inferiority trial. *PLoS Med.* 2017;**14**(10):e1002411.
20. Maiga H, Barger B, Sagara I, Guindo A, Traore OB, Tekete M, *et al*. Impact of three-year intermittent preventive treatment using artemisinin-based combination therapies on malaria morbidity in malian schoolchildren. *Tropical Medicine and Infectious Disease.* 2020;**5**(3):148.
21. Maitland K, Kiguli S, Olupot-Olupot P, Engoru C, Mallewa M, Saramago Goncalves P, *et al.* Immediate Transfusion in African Children with Uncomplicated Severe Anemia. *N Engl J Med.* 2019;**381(**5):407-19.
22. Maitland K, Olupot-Olupot P, Kiguli S, Chagaluka G, Alaroker F, Opoka R, *et al.* Transfusion Volume for Children with Severe Anemia in Africa. *The New England journal of medicine.* 2019;**381:**420-31.
23. George E, Uyoga S, M'Baya B, Byabazair D, Kiguli S, Olupot-Olupot P, *et al.* Whole blood versus red cell concentrates for children with severe anaemia: a secondary analysis of the Transfusion and Treatment of African Children (TRACT) trial. *The Lancet Global Health.* 2022;**10**:e360-e8.
24. Maitland K, Kiguli S, Opoka RO, Engoru C, Olupot-Olupot P, Akech SO, *et al*. Mortality after fluid bolus in African children with severe infection. *N Engl J Med.* 2011;**364**(26):2483-95.
25. Maitland K, George EC, Evans JA, Kiguli S, Olupot-Olupot P, Akech SO, *et al.* Exploring mechanisms of excess mortality with early fluid resuscitation: insights from the FEAST trial. *BMC Med.* 2013;**11**:68.
26. Kiguli S, Maitland K, George EC, Olupot-Olupot P, Opoka RO, Engoru C, *et al.* Anaemia and blood transfusion in African children presenting to hospital with severe febrile illness. *BMC Med.* 2015;**13**:21-.
27. McGann PT, Tyburski EA, de Oliveira V, Santos B, Ware RE, Lam WA. An accurate and inexpensive color-based assay for detecting severe anemia in a limited-resource setting. *Am J Hematol.* 2015;**90**(12):1122-7.
28. Mtove G, Amos B, von Seidlein L, Hendriksen I, Mwambuli A, Kimera J, *et al*. Invasive salmonellosis among children admitted to a rural Tanzanian hospital and a comparison with previous studies. *PloS one.* 2010;**5**(2):e9244.
29. Nass SA, Hossain I, Sanyang C, Baldeh B, Pereira DIA. Hemoglobin point-of-care testing in rural Gambia: Comparing accuracy of HemoCue and Aptus with an automated hematology analyzer. *PLoS One.* 2020;**15**(10):e0239931.
30. Nkrumah B, Nguah SB, Sarpong N, Dekker D, Idriss A, May J, *et al.* Hemoglobin estimation by the HemoCue® portable hemoglobin photometer in a resource poor setting. *BMC Clin Pathol.* 2011;**11**:5.
31. Ntonifor HN, Chewa JS, Oumar M, Mbouobda HD. Intestinal helminths as predictors of some malaria clinical outcomes and il-1beta levels in outpatients attending two public hospitals in bamenda, north west cameroon. *PLoS Neglected Tropical Diseases.* 2021;**15**(3):e0009174.
32. Ocan A, Oyet C, Webbo F, Mwambi B, Taremwa IM. Prevalence, morphological characterization, and associated factors of anemia among children below 5 years of age attending St. Mary's Hospital Lacor, Gulu District, Northern Uganda. *Journal of blood medicine.* 2018;**9**:195-201.

33. Olatunya OS, Oke OJ, Kuti BP, Ajayi IA, Olajuyin O, Omotosho-Olagoke O, *et al.* Factors influencing the academic performance of children with sickle cell anaemia in Ekiti, South West Nigeria. *Journal of Tropical Pediatrics.* 2018;**64**(1):67-74.
34. Olatunya OS, Olu-Taiwo A, Ogundare EO, Oluwayemi IO, Olaleye AO, Fadare JO, *et al.* Evaluation of a Portable Haemoglobin Metre Performance in Children with Sickle Cell Disease and Implications for Healthcare in Resource-poor Settings. *Journal of tropical pediatrics*. 2016;**62**(4):316-23.
35. Olupot-Olupot P, Prevatt N, Engoru C, Nteziyaremye J, Amorut D, Chebet M, *et al.* Evaluation of the diagnostic accuracy and cost of different methods for the assessment of severe anaemia in hospitalised children in Eastern Uganda. *Wellcome open research.* 2018;**3**:130.
36. Onyangore FO, Were GM, Mwamburi LA. Prevalence of iron deficiency anaemia and dietary iron intake among infants aged six to nine months in Keiyo South Sub County, Kenya. *African Journal of Food, Agriculture, Nutrition and Development.* 2016;**16**(2):10884-97.
37. Parbey PA, Tarkang E, Manu E, Amu H, Ayanore MA, Aku FY, *et al*. Risk Factors of Anaemia among Children under Five Years in the Hohoe Municipality, Ghana: A Case Control Study. *Anemia.* 2019;2019:2139717.
38. Parker M, Han Z, Abu-Haydar E, Matsiko E, Iyakaremye D, Tuyisenge L, *et al.* An evaluation of hemoglobin measurement tools and their accuracy and reliability when screening for child anemia in Rwanda: A randomized study. *PloS one.* 2018;**13**(1):e0187663.
39. Ramaswamy G, Vohra K, Yadav K, Kaur R, Rai T, Jaiswal A, *et al*. Point-of-Care Testing Using Invasive and Non-Invasive Hemoglobinometers: Reliable and Valid Method for Estimation of Hemoglobin among Children 6-59 Months*. J Trop Pediatr.* 2021;**67**(1).
40. Silva DLF, Hofelmann DA, Taconeli CA, Lang RMF, Dallazen C, Tietzmann DC, *et al*. Individual and contextual predictors of children's hemoglobin levels from Southern Brazilian municipalities in social vulnerability. *Cadernos de saude publica.* 2021;**36**(12):e00166619.
41. Smart LR, Ambrose EE, Raphael KC, Hokororo A, Kamugisha E, Tyburski EA, *et al*. Simultaneous point-of-care detection of anemia and sickle cell disease in Tanzania: the RAPID study. *Ann Hematol.* 2018;**97**(2):239-46.
42. Tack B, Vita D, Mansosa I, Mbaki TN, Wasolua N, Luyindula A, *et al.* Field Experiences with Handheld Diagnostic Devices to Triage Children under Five Presenting with Severe Febrile Illness in a District Hospital in DR Congo. *Diagnostics (Basel)*. 2022;**12**(3).
43. Teshome EM, Andang'o PEA, Osoti V, Terwel SR, Otieno W, Demir AY, *et al*. Daily home fortification with iron as ferrous fumarate versus NaFeEDTA: a randomised, placebo-controlled, non-inferiority trial in Kenyan children. *BMC Med.* 2017;**15**(1):89.
44. Ughasoro MD, Madu AJ, Kela-Eke IC, Akubuilo U. Parental Perception of Childhood Anaemia and Efficiency of Instrument Assisted Pallor Detection among Mothers in Southeast Nigeria: A Field Validation Study. *International Journal of Pediatrics*. 2019;2019.
45. Ughasoro MD, Madu AJ, Kela-Eke IC. Evaluation of the Performance of Haemoglobin Colour Scale and Comparison with HemoCue Haemoglobin Assay in Diagnosing Childhood Anaemia: A Field Validation Study. *Int J Pediatr.* 2019;2019:3863070.
46. ClinicalTrials.gov [Internet]. Therapeutic Efficacy Study of AL and DP in Western Kenya. 2022; Available from: https://clinicaltrialsgov/show/NCT05060198. nct Identifier: NCT05060198. Accessed 20 June 2022.
47. ClinicalTrials.gov [Internet]. A Trial of 2 'Point of Care' Diagnostic Methods to Improve Detection and Treatment of Anaemia in African Children (EARS). 2022; Available from: https://clinicaltrials.gov/ct2/show/NCT00439595. nct Identifier: NCT00439595. Accessed 20 June 2022.
48. Whiting PF, Rutjes AW, Westwood ME, Mallett S, Deeks JJ, Reitsma JB, *et al.* QUADAS-2: a revised tool for the quality assessment of diagnostic accuracy studies. *Ann Intern Med.* 2011;**155**(8):529-36.
